# Effects of Soluble Corn Fiber Consumption on Executive Functions and Gut Microbiota in Middle to Older Age Adults: A Randomized Controlled Crossover Trial

**DOI:** 10.1101/2025.06.12.25329532

**Authors:** David A. Alvarado, Tori A. Holthaus, Shelby Martell, Nicole L. Southey, Marco Atallah, Rhea Sarma, David Revilla, Marina Brown, Twinkle Mehta, Naiman A. Khan, Hannah D. Holscher

## Abstract

**Background:** Dietary fiber may support cognition through gastrointestinal-microbiota mechanisms, but clinical evidence is limited.

**Objectives:** We aimed to determine whether soluble corn fiber (SCF) improved cognition and altered fecal microbiota and fermentation end products in adults.

**Methods:** In a randomized, double-blind, crossover trial, 42 healthy adults (45-75y) consumed SCF (18g/d) or a maltodextrin placebo control (CON: 22g/d) for 4 weeks, separated by a washout. Cognitive outcomes included executive function with event-related potentials, relational memory, neuropsychological performance, and mood. Secondary outcomes included fecal microbiota, metabolomics, and gastrointestinal tolerance. Tertiary analyses related microbial and metabolite changes to cognitive improvements using correlation, mediation, and moderation models, and explored SCF fermentation pathways with 16S-predicted functional profiling, shotgun metagenomics and *in vitro* culturing.

**Results:** SCF improved reaction times (RT) during congruent (β = -9.8 ms, 95% CI: [-18.4, -1.2], FDR *P =* 0.01) and incongruent (β = -14.2 ms, 95% CI: [-22.8, -5.6], FDR *P =* 0.003) flanker trials and increased *Parabacteroides* (∼4-fold, β = 1.44 log, 95% CI [1.01, 1.88], FDR *P <* 0.001). At the SCF endpoint, congruent RT tended to be inversely associated with fecal acetate (ρ = -0.33) and propionate (ρ = -0.36), while *Parabacteroides* was marginally positively associated with acetate (ρ = 0.34) (all FDR *P <* 0.1). Moderation analyses indicated that SCF-RT relation varied by *Parabacteroides* magnitude change. At endpoint, SCF increased predicted functional potential of carbohydrate-related KOs and pathways (FDR *P <* 0.05). *In vitro* culturing confirmed *P. distasonis* ferments SCF.

**Conclusion:** SCF consumption improved attentional inhibition, altered the gut microbiota, and selectively enriched *Parabacteroides*. Although mediation analyses did not support a direct microbiota-to-cognition pathway, moderation analyses suggested that SCF-related cognitive effects may depend in part on *Parabacteroides* abundance. Collectively, these findings suggest that certain cognitive benefits of SCF consumption may be partly underpinned by the gut microbiota.

## Introduction

A healthy diet is a key determinant of metabolic health, and emerging evidence extends these benefits to brain function and cognition ^1–3^. Dietary fiber is associated with reduced risk of obesity, cardiovascular disease, type 2 diabetes, and colon cancer ^4,5^, yet nearly 95% of Americans fail to meet recommended intake levels ^6^. Further, dietary fiber provides a primary substrate for microbes residing in the gastrointestinal (GI) tract, shaping ecological community diversity and metabolic capacity ^7^. Preclinical studies show that dietary fibers improve cognition ^8,9^, but human trials remain limited ^10–12^, particularly those linking the microbiota, GI-derived metabolites, and cognitive outcomes ^13^. Therefore, further evidence is necessary to comprehensively characterize the effect of dietary fiber on general and specific domains of cognitive function.

Soluble corn fiber (SCF) is a resistant maltodextrin with an average degree of polymerization (DP) between 10-12 units ^14^, that supports glycemic control, calcium absorption, and gastrointestinal health ^15–17^. Its non-viscous, water-soluble structure lacks β-linkages, yet branching of the α-linkages confers resistance to complete digestion in the small intestine (<30%) ^18,19^. The undigested fraction undergoes colonic fermentation, enriching saccharolytic taxa such as *Bifidobacterium*, *Lactobacillus*, and *Parabacteroides* ^20,21^, and increasing short-chain fatty acids (SCFA) concentrations across age groups ^22^, while reducing deleterious fermentation end products (phenols, indoles, ammonia) ^19,23^. SCFAs are increasingly recognized as key mediators of GI-brain communication by binding free fatty acid receptors (FFAR/FFAR3) to regulate immune ^24^ and enteroendocrine signaling ^25^, modulate microglial activity and systemic inflammation ^26^, and stimulate vagal and serotonergic pathways ^27–30^.

Executive functions, including inhibitory control, are central to attention, working memory, and flexible behavior ^31^. These top-down processes can be assessed through computer-based tasks that measure a subject’s ability to maintain attention while faced with distracting stimuli, hold and manipulate information in mind, and require multi-tasking and set-shifting, while electrophysiological markers can provide complementary insight into neural activity during such tasks ^32,33^. Given the growing evidence that microbially derived metabolites influence brain activity ^27,34–39^, assessing both behavioral and neural indices provides a comprehensive overview to evaluate dietary fiber’s cognitive impact.

To address these gaps, we conducted a randomized, double-blind, placebo-controlled, crossover trial to test the impact of daily SCF consumption in middle- to older-aged adults. The primary aim was to determine whether four weeks of SCF consumption improved executive function, changes in the amplitude (size) and latency (timing) of the P3 event-related potential (ERP), as well as additional behavioral tasks assessing relational memory and neuropsychological function. We hypothesized that SCF would enhance cognitive control and reduce the number of neural resources required to accomplish these cognitive processes. The secondary aim examined the effects of SCF on fecal microbiota composition and fermentation metabolites, hypothesizing that SCF consumption would enrich health-associated taxa and effect microbial-derived metabolites. Finally, the tertiary aim tested whether the SCF-induced changes in microbial or metabolite profiles explained changes in cognitive outcomes, hypothesizing that these changes would contribute to the cognitive benefits induced by SCF.

## Methods

### Study design and intervention

This clinical trial, registered as the SCOPE study (Soluble Corn fiber for Promoting Executive function; NCT05066425), included two 4-week intervention periods separated by an ∼4-week washout. Participants consumed soluble corn fiber (SCF; PROMITOR® Soluble Fibre™, 22g/d containing 18g fiber) or the maltodextrin placebo control (CON; 22g/d) in a counterbalanced order. The two interventions were provided by Tate & Lyle in identical, pre-measured sachets that were indistinguishable in taste and appearance. Sachets were labeled as 789 or 030, and participants and research staff remained blinded to the code assignment until data analysis was complete to ensure blinding. Participants were given 35 sachets per condition and were instructed to consume one daily, dissolved in a fluid of their choice. Participants recorded intake of the sachet on daily compliance logs. Unused sachets and logs were collected at the end of each period to assess study adherence.

All study visits and testing sessions were conducted at the University of Illinois Urbana-Champaign (Supplemental **Figure 1**). Anthropometric measures, physical activity level, demographic information, and medical history were collected at baseline and visits 4, 5, and 6 as potential covariates.

### Participants criteria

Adults aged 45-75 years were recruited from the East Central Illinois area using email newsletters, social media, community postings using the postal service, and flyers posted on public transport. All participants were informed of all procedures and provided their written informed consent before any data was collected. The SCOPE study was approved by the University of Illinois Institutional Review Board (May 2021) and conducted following the principles of the Declaration of Helsinki.

Exclusion criteria included physician-diagnosed gastrointestinal or metabolic disease (inflammatory bowel disease and diabetes), neurological disorder (dementia, Parkinson’s disease, multiple sclerosis, brain tumors, autism), colorblindness or vision not corrected to 20/20, antibiotic use within the past month, tobacco use, food allergies, pregnancy, or lactating, BMI <18.5 or >34.9 kg/m^2^, Mini-Mental State Examination <24, or if habitual dietary fiber intake was >70% of the daily recommended intake (≥18 g/d for women, ≥27 g/d for men) using the Dietary History Questionnaire (*DHQ3*) food frequency questionnaire.

Eligible participants agreed to maintain habitual dietary patterns and physical activity, to avoid probiotic or prebiotics supplements, and to report any changes in medications during the SCOPE study.

Enrolled participants (**Table 1**) were randomized via coin toss to start one of the two intervention conditions. Height and weight were measured in triplicate, without shoes and in light clothing, to determine BMI using a *SECA* stadiometer (model 240; *SECA*, Hamburg, Germany) and a Tanita WB-300 Plus digital scale (Tanita, Tokyo, Japan). Physical activity was assessed using the Godin-Shephard Leisure-Time Exercise Questionnaire (*Godin*), which records the frequency of strenuous, moderate, and light physical activities performed during a typical week, which are totaled into a final score using a weighted formula ^40^.

**Table 1.** Baseline demographic and clinical characteristics of participants randomized to treatment counterbalanced.

|  | Overall<br>N = 42 | CON<br>N = 21 | SCF<br>N = 21 |
| --- | --- | --- | --- |
| **<u>Education</u>** |  |  |  |
| *High School Graduate* | 2 (4.8%) | 0 (0%) | 2 (9.5%) |
| *Some college* | 6 (14%) | 3 (14%) | 3 (14%) |
| *Bachelor's Degree* | 13 (31%) | 8 (38%) | 5 (24%) |
| *Advanced Degree* | 21 (50%) | 10 (48%) | 11 (52%) |
| **<u>Ethnicity</u>** |  |  |  |
| *Asian* | 6 (14%) | 3 (14%) | 3 (14%) |
| *Black or African American* | 3 (7.1%) | 1 (4.8%) | 2 (9.5%) |
| *Mixed or Other* | 1 (2.4%) | 0 (0%) | 1 (4.8%) |
| *White (Caucasian)* | 32 (76%) | 17 (81%) | 15 (71%) |
| *Latino* | 1 (2.4%) | 1 (4.8%) | 0 (0%) |
| **<u>Sex</u>** |  |  |  |
| *Female* | 28 (67%) | 15 (71%) | 13 (62%) |
| *Male* | 14 (33%) | 6 (29%) | 8 (38%) |
| **<u>Age (years)</u>** | 56.2 ± 7.3 | 54.8 ± 6.2 | 57.5 ± 8.2 |
| **<u>Fiber per 1000kcal</u>** | 11.4 ± 3.4 | 12.0 ± 3.3 | 10.8 ± 3.5 |
| **<u>BMI (kg/m<sup>2</sup>)</u>** | 25.9 ± 4.0 | 25.4 ± 3.7 | 26.3 ± 4.2 |
| **<u>Income</u>** |  |  |  |
| *<\$31,000-40,000* | 3 (9.5%) | 1 (4.8%) | 2 (14.3%) |
| *\$41,000-50,000* | 3 (7.1%) | 1 (4.8%) | 2 (9.5%) |
| *\$51,000-60,000* | 3 (7.1%) | 2 (9.5%) | 1 (4.8%) |
| *\$61,000-70,000* | 4 (9.5%) | 1 (4.8%) | 3 (14%) |
| *\$71,000-80,000* | 5 (12%) | 4 (19%) | 1 (4.8%) |
| *\$81,000-90,000* | 1 (2.4%) | 1 (4.8%) | 0 (0%) |
| *\$91,000-100,000* | 7 (16.7%) | 4 (19%) | 3 (14%) |
| *Greater than \$100,000* | 15 (35.7%) | 7 (33%) | 8 (38%) |

Baseline characteristics of participants enrolled and randomized into the clinical trial (N = 42), grouped by initial treatment assignment. Dietary fiber (g/1000 kcal) values reflect baseline intake assessed using DHQ3 prior to the first intervention period. Height and weight were measured at the baseline visit and used to calculate body mass index (BMI; kg/m<sup>2</sup>). Continuous variables are reported as mean ± standard deviation; categorical variables are reported as number of participants and percentage within each group [n (%)].

**Table 2.** Behavioral and ERP measures during the flanker task across treatment and time.

| | Maltodextrin | | Soluble Corn Fiber | | Treatment Effect (ΔSCF - ΔCON) | | | |
| --- | --- | --- | --- | --- | --- | --- | --- | --- |
|  | PRE | POST | PRE | POST |  |  |  |  |
| Behavioral Flanker | Mean ± SEM (*n*) | Mean ± SEM (*n*) | Mean ± SEM (*n*) | Mean ± SEM (*n*) | $\beta$ | SE | 95% CI | *p* |
| **Accuracy (%)** |  |  |  |  |  |  |  |  |
| Congruent | 96.9 ± 0.52 (39) | 95.5 ± 1.37 (39) | 97.0 ± 0.48 (40) | 97.6 ± 0.45 (40) | 0.65 | 0.8 | [-0.93, 2.23] | 0.42 |
| Incongruent | 92.4 ± 1.00 (39) | 92.0 ± 1.53 (39) | 93.6 ± 0.79 (40) | 94.4 ± 0.70 (40) | -0.68 | 1.1 | [-2.82, 1.46] | 0.53 |
| **Reaction Time (ms)** |  |  |  |  |  |  |  |  |
| Congruent | 395. ± 5.79 (39) | **398. ± 6.69<sup>a</sup>** (39) | 400.71 ± 6.22 (40) | **389. ± 5.52<sup>b</sup>** (40) | -18.4 | 4.2 | [-18.4, -1.2] | **0.01\*** |
| Incongruent | 431. ± 6.62 (39) | **437. ± 7.77<sup>a</sup>** (39) | 435.43 ± 6.88 (40) | **424. ± 6.35<sup>b</sup>** (40) | -14.2 | 4.4 | [-22.8, -5.6] | **0.003\*\*** |
| **ERP Flanker** |  |  |  |  |  |  |  |  |
| **Amplitude (μV)** |  |  |  |  |  |  |  |  |
| Congruent | 5.96 ± 1.73 (39) | 6.78 ± 0.68 (36) | 6.64 ± 0.60 (39) | 7.07 ± 0.77 (37) | 0.88 | 1.18 | [-2.22, 1.01] | 0.46 |
| Incongruent | 6.36 ± 1.64 (39) | 6.77 ± 0.69 (36) | 7.14 ± 0.65 (39) | 7.50 ± 0.79 (37) | 1.08 | 1.10 | [-1.06, 3.24] | 0.33 |
| **Peak Latency (ms)** |  |  |  |  |  |  |  |  |
| Congruent | 386. ± 6.71 (39) | 399. ± 10.18 (36) | 394. ± 7.29 (39) | 398. ± 7.99 (37) | -6.16 | 0.32 | [-31.1, 18.7] | 0.67 |
| Incongruent | 424. ± 8.34 (39) | 443. ± 11.70 (36) | 438. ± 9.90 (39) | 429. ± 8.27 (37) | -23.0 | 15.4 | [-53.2, 6.81] | 0.14 |

Values are means ± standard error of the mean (SEM) and sample size (*n*) across congruencies at each timepoint. Measures include accuracy (%), reaction time (ms), P3 amplitude ( μV), and P3 peak latency (ms). Linear mixed-effect models included participant ID as a random effect, and the fixed effects of treatment, time, and their interaction. Reported are the treatment-by-time interaction estimates ( $\beta$ , Standard Error (SE), 95% confidence interval (CI), *p*-values). Pairwise comparisons were conducted within each congruency when a treatment-by-time interaction met the significance threshold ( $P < 0.05$ ), with *p*-values adjusted using the Benjamin-Hochberg procedure. Superscripts (a, b) denote significant *pairwise* differences within congruency trials between treatments and time. Significance threshold *p*: ‘\*\*\*’ $< 0.01$ , ‘\*\*’ $< 0.05$ , ‘†’ $< 0.1$ .

**Table 3.** 16S Alpha diversity and differential abundance of genera across intervention periods.

| | Maltodextrin | | Soluble Corn Fiber | | Treatment Effect (ΔSCF - ΔCON) | | | |
| --- | --- | --- | --- | --- | --- | --- | --- | --- |
| | PRE | POST | PRE | POST | $\beta$ | SE | 95% CI | FDR *P* |
| **16S ASVs** | Mean ± SEM (*n*) | Mean ± SEM (*n*) | Mean ± SEM (*n*) | Mean ± SEM (*n*) | | | | |
| **Genera** (% of sequence) |  |  |  |  |  |  |  |  |
| *Bacteroides* | 25.2 ± 1.77 (38) | 23.7 ± 1.90 (38) | 27.3 ± 2.06 (37) | 22.6 ± 2.07 (37) | -0.15 | 0.10 | [-0.34, 0.05] | 0.11 |
| *Faecalibacterium* | 10.2 ± 1.02 (38) | 10.1 ± 1.11 (38) | 8.66 ± 1.15 (37) | 11.0 ± 1.30 (37) | 0.23 | 0.21 | [-0.18, 0.64] | 0.47 |
| *Parabacteroides* | 2.52 ± 0.35<sup>a</sup> (38) | **2.06 ± 0.24<sup>a</sup>** (38) | 2.09 ± 0.26<sup>a</sup> (37) | **8.37 ± 0.98<sup>b</sup>** (37) | 1.44 | 0.22 | [1.01, 1.88] | **<0.001\*\*\*** |
| *Alistipes* | 3.46 ± 0.51 (38) | 2.82 ± 0.35 (38) | 3.88 ± 0.59 (37) | 3.96 ± 0.69 (37) | 0.08 | 0.20 | [-0.31, 0.46] | 0.50 |
| *Blautia* | 3.76 ± 0.50 (38) | 4.52 ± 0.61 (38) | 3.54 ± 0.42 (37) | 3.24 ± 0.50 (37) | -0.27 | 0.13 | [-0.52, -0.02] | 0.10† |
| *Agathobacter* | 4.16 ± 0.88 (38) | 5.97 ± 1.17 (38) | 3.99 ± 0.85 (37) | 2.80 ± 0.58 (37) | -0.46 | 0.23 | [-0.91, 0.01] | 0.11 |
| *Subdoligranulum* | 2.31 ± 0.29 (38) | 2.54 ± 0.38 (38) | 3.24 ± 0.58 (37) | 2.69 ± 0.49 (37) | -0.24 | 0.22 | [-0.68, 0.19] | 0.83 |
| *Prevotella* | 0.87 ± 0.41 (38) | 1.64 ± 0.74 (38) | 1.60 ± 0.89 (37) | 2.13 ± 1.04 (37) | -0.22 | 0.30 | [-0.81, 0.37] | 0.53 |
| *Dialister* | 1.34 ± 0.44<sup>a</sup> (38) | **1.46 ± 0.51<sup>a</sup>** (38) | 0.93 ± 0.34<sup>a</sup> (37) | **2.10 ± 0.63<sup>b</sup>** (37) | 1.06 | 0.39 | [0.29, 1.83] | **0.02\*** |
| *Bifidobacterium* | 2.50 ± 0.53 (38) | 2.69 ± 0.53 (38) | 2.12 ± 0.52 (37) | 2.09 ± 0.41 (37) | 0.09 | 0.31 | [-0.51, 0.70] | 0.92 |
| *Fusicatenibacter* | 0.60 ± 0.10<sup>a</sup> (38) | **0.67 ± 0.09<sup>a</sup>** (38) | 0.53 ± 0.11<sup>a</sup> (37) | **1.79 ± 0.28<sup>b</sup>** (37) | 1.10 | 1.10 | [0.59, 1.61] | **<0.001\*\*\*** |
| **Alpha Diversity Metrics** |  |  |  |  |  |  |  |  |
| Richness | 439.9 ± 19.8<sup>a</sup> (38) | **438.7 ± 18.2<sup>a</sup>** (38) | 443.6 ± 17.6<sup>a</sup> (37) | **413.7 ± 18.6<sup>b</sup>** (37) | -0.76 | 0.29 | [-1.32, -0.19] | **0.03\*** |
| Shannon | 3.89 ± 0.06 (38) | 3.88 ± 0.06 (38) | 3.81 ± 0.06 (37) | 3.71 ± 0.07 (37) | -0.08 | 0.07 | [-0.22, 0.06] | 0.26 |
| Faith's PD | 104.7 ± 1.62 (38) | 103.9 ± 1.54 (38) | 105.23 ± 1.54 (37) | 101.9 ± 1.80 (37) | -2.43 | 1.28 | [-4.93, 0.06] | 0.09† |

Values are means ± standard error of the mean (SEM) and sample size (*n*) across each timepoint. For differential abundance of 16S genera, a generalized linear mixed-effect negative binomial model included participant ID as a random effect, and the fixed effects of treatment, time, and their interaction. Reported are the treatment-by-time interaction log-scaled estimates ( $\beta$ , Standard Error (SE), 95% confidence interval (CI), FDR-adjusted *p*-values). For alpha diversity metrics, linear mixed-effect model. Reported are the treatment-by-time interaction estimates. Pairwise comparisons was conducted if an interaction (*treatment effect*) met the significance threshold ( $P < 0.05$ ), *p*-values adjusted using the Benjamin-Hochberg procedure. Superscripts (a, b) denote significant *pairwise* treatment effect. Significance threshold *p*: '\*\*\*' <0.001, '\*\*' <0.01, '\*' <0.05, '†' <0.1.

#### Aim 1: Cognitive Outcomes

To minimize learning effects, participants completed practice sessions at the start of each visit until at least 70% accuracy was met before experimental cognitive sessions were administered. The order of cognitive task administration at each testing visit (visits 3-6) was counterbalanced using a 2x2 Latin square design for each participant,

### Executive function

Attentional inhibition was assessed using a modified Eriksen flanker task as previously described ^41^. Participants were asked to respond (*Current Designs*, Philadelphia, PA, USA) quickly and accurately to the directionality of a central stimulus (3-cm tall white arrow) presented with two flanking stimuli (arrows) on each side (83s stimulus presentation; 100ms response window; jittered intertrial intervals of 1100, 1200, 1500ms). In congruent trial types, the flanking stimuli matched the direction of the central arrow (<<<<<, >>>>>); during incongruent trials, flanking stimuli opposed the central arrow (>><>>, <<><<), requiring greater inhibitory control. The task began with 40 practice trials followed by two blocks of 200 randomized experimental trials. Behavioral measures were accuracy (ACC) and reaction time (RT) for correct responses. Interference effects (flanker effect) for ACC and RT were calculated, respectively, by subtracting incongruent from congruent measures (ACC) and congruent from incongruent measures (RT). These measures index the ability to maintain task performance when resolving conflicting information.

### Neuroelectric indices

EEG activity was recorded simultaneously during the flanker task via a Neuro-scan Quik-cap with 64 passive, wet, sintered Ag/AgCl scalp electrodes arranged in the international 10–10 system, with electrodes placed near the outside of each eye and above and below the left eye ^42^. A midline sensor placed between CZ and CPZ served as a reference, and AFZ served as the ground. Continuous EEG data were digitized using a Neuroscan SynampsRT amplifier at a sampling rate of 500 Hz, amplified 500 times to an online low-pass 70-Hz filter with a direct current and a 60-Hz notch filter. Electrode impedance values were maintained at ≤20 kohms.

Signal processing was performed in MATLAB (MathWorks, Natick, MA, version 2021b) using EEGLab and ERPlab ^43,44^. Continuous data were re-referenced to the average of two mastoid electrodes. A trained research team member visually inspected all channels for excessive noise during pre-processing. Channels with excessive noise were interpolated with an average of the adjacent electrodes. Data were submitted to a 0.1 Hz high-pass Butterworth filter before eye-blink artifacts were rejected using independent component analysis (ICA). ICA and vertical eye sensor channel correlations greater than 0.35 were considered eye-blinks. A time window of -200 to 1200 ms around stimulus presentation was used for creating stimulus-locked epochs with a baseline correction of -200 ms to stimulus onset. A 30-Hz low-pass filter was used. Artifacts were detected and rejected if a moving window peak-to-peak amplitude exceeded 100 μV, using a 100-ms window width and step. Only correct trials with >80% of artifact-free trials in all flanker task trials were retained, and participants with excessive artifacts (>20%) were excluded (*n* = 3 CON, *n* = 4 SCF), consistent with previous literature ^45,46^.

Electrodes of interest were chosen based on a collapsed localizer technique ^47^. Evidence for selection was observed from maximal voltage localization of combined baseline and post-testing trials collapsed across condition and trial type. *Post-hoc* topographic average plots were constructed using a stylized topographic map plugin for EEGLAB/ERPLAB ^48^. The P3 component was defined as the region of interest comprised of C1, CZ, C2, CPZ, CP1, and CP2 with a localized peak and corresponding latency between 300 and 600ms post-stimulus onset. Amplitude and latency were extracted to complement the analysis of the behavioral Flanker task.

### Relational memory

Hippocampal-dependent relational memory was assessed using the computerized spatial reconstruction task presentation software (*iPosition*; Neurobehavioral Systems, Berkeley, CA) ^49^. Participants studied arrays of six abstract shapes on the computer screen in a randomized array for 6 seconds. The shapes were then masked by black squares for 18 seconds and could be revealed individually by clicking on them. After a 2-second delay, the stimuli reappeared at the top of the screen, and participants were asked to reconstruct the original array. Each participant completed 20 trials with randomized arrays. Two error metrics were used to assess performance: misplacement error (original misplacement), calculated as the average measure of distance between the actual and reconstructed stimuli locations, and object-location binding (accurate single item placement), the number of times the participant correctly placed a stimulus within a pre-defined radius.

### Neuropsychological battery

Additional cognitive performance across multiple domains was assessed using the Cambridge Neuropsychological Test Automated Battery (*CANTAB*, Cambridge Cognition Ltd) ^50,51^, administered on a touchscreen tablet. The battery included the Intra-Extra Dimensional Set Shift (IED), Stop Signal Task (SST), Rapid Visual Information Processing (RVP), and Spatial Working Memory (SWM). The IED is a test which assesses rule acquisition and reversal, attentional set formation, maintenance, and shifting. The RVP task assesses sustained attention, and the SST measures response inhibition. The SWM task assesses spatial working memory.

### Mood and Stress

Mood and stress were assessed using validated self-report instruments, including the Profile of Mood States (*POMS*), the Positive and Negative Affect Schedule (*PANAS*), and the Perceived Stress Scale (*PSS*). The *POMS* questionnaire includes 65-items that evaluates transient mood states across six subscales (tension/anxiety, depression, anger, vigor, fatigue, and confusion), with ratings from 0 (“not at all”) to 5 (“extremely”) ^52,53^. *PANAS* includes 20 items divided evenly into positive and negative affect subscales, scored from 1 (“very slightly or not at all”) to 5 (“extremely likely”) ^53,54^. *PSS-10* is a 10-item measure of stress over the past month, scored from 0 (“never”) to 4 (“very often”) ^55^.

### Aim 2: Gut Microbiota and Metabolite Outcomes

#### Fecal collection and processing

Participants provided two fecal samples at each of the four collection timepoints (baseline and end of each of the two study conditions) for a total of 8 samples per participant. Samples were collected during the five days preceding the first intervention and the final five days of each treatment and washout period. Samples were delivered within 15 minutes of defecation using provided kits (Commode Specimen Collection Systems, *Sage Products*) with ice packs and coolers. Upon arrival at the laboratory, samples were homogenized, aliquoted into cryovials for sequencing, flash-frozen in liquid nitrogen, and stored at -80°C. Additional aliquots were prepared for metabolite analysis: SCFAs, BCFAs, ammonia, and dry matter basis (DMB; pre-weighed aluminum tins). These aliquots were stored at -20°C until analysis. Fecal pH was measured (*Denver Instrument*) after samples had been aliquoted.

#### Fecal DNA isolation and 16S amplicon analyses

Fecal microbial DNA was extracted using the Power-Lyzer PowerSoil DNA Isolation Kit (*QIAGEN*). DNA was verified using 1% agarose gel electrophoresis, concentrations measured using a Qbit 3.0 Fluorometer (*ThermoFisher Scientific*), quantified from two samples collected per timepoint per participant, and pooled in equimolar amounts before sequencing at the University of Illinois W.M Keck Center for Biotechnology. 16S Amplicons were generated with V4 primers (*F*: 5’-GTGYCAGMGCCGCGGTAA, *R*: 5’-GGACTACNVGGGTWTCTAAT) ^56^ and sequenced on an Illumina NovaSeq 6000 SP (2x250 nt), producing ∼1 billion reads. Sequences with quality scores >25 were processed with *DADA2* ^57^ integrated in *QIIME2* (v2023.7) ^58^. Sequences were denoised and resolved into amplicon sequence variants (ASVs) using the *SILVA* 138-99 (515F/806R) naïve Bayes reference classifier ^57,59^. Samples were normalized to 350,000 reads; curves plateaued at this depth, indicating adequate coverage (**Supplemental Figure 2**). Rarefaction was performed with *vegan* v2.6.8 ^60^, filtering 1,521 ASVs (13%, primarily singletons or zero-count features) and retaining 10,373 ASVs. Alpha diversity and beta diversity were calculated with *phyloseq* (v1.48.0) ^61^, and Faith’s phylogenetic diversity (Faith’s PD) with *picante* (v1.8.2). Taxonomic summaries were created at the genus and phylum levels for downstream analyses.

#### Targeted Metabolites

Fecal dry matter was measured according to the Association of Official Analytical Chemists, 1984, methods ^62^. Ammonia concentrations were measured following the procedure of Chaney and Marbach ^63^. Aliquots for SCFA and BCFA were weighed, acidified with 2N hydrochloric acid, and stored at -20°C until analysis. Briefly, SCFA (acetate, propionate, butyrate), BCFA (isovalerate, valerate, and isobutyrate) concentrations were determined on the supernatant fraction of the acidified fecal samples using gas chromatography mass spectroscopy (GC-MS). Concentrations were calculated using standards^64^. Phenol and indole were quantified, according to Flickinger et al.^65^ Fecal metabolites are reported on the normalized DMB scale.

#### Digestive health

GI symptoms, bowel function, and stool characteristics were assessed using 7-day self-reported records, questionnaires, and laboratory measures for each timepoint ^66^. Participants rated GI symptom severity (burping, cramping, bloating, gas, nausea, reflux, rumbling) on a 4-point scale from 1 (“absent”) to 4 (“severe”) ^67^. Bowel movements and consistency were scored using a 7-point Bristol stool scale (1- “separate hard lumps”; 2- “sausage-shaped but lumpy”; 3- “sausage-like but with cracks on the surface”; 4- “sausage-like, smooth, and soft”; 5- “soft blobs with clear cut edges”; 6- “fluffy pieces, mushy”; 7- “entirely liquid”). And ease of passage rated on a 5-point scale from 1 (“very easy”) to 5 (“very difficult”). In the laboratory, fresh fecal samples were measured with a pH meter (Denver Instrument) ^66^, and a trained research team member recorded stool consistency using the Bristol scale.

### Aim 3: Exploratory Integrative and Functional Analyses

#### Mediation and moderation

Selected microbial and metabolite features for integrative analyses were prioritized from Aim 1 and 2 outcomes that met the statistical significance threshold and were also included when they showed consistent directional trends and biological plausibility based on prior literature ^27,34–39^. Associations between these candidates and cognitive outcomes were examined using Spearman correlations. We then used nonparametric bootstrapped causal mediation models to estimate indirect and direct effects within a causal inference framework ^68–70^. Moderation analyses were conducted using the Johson-Neyman framework with Aiken & West’s simple-slopes procedures ^71,72^. Together, these complementary approaches will provide further clarity on whether microbes or metabolites acted as intermediaries, transmitting the treatment (mediation), or as variables that altered the strength or direction of the treatment’s effect (moderation).

#### In vitro isolated culturing

*Parabacteroides distasonis* (*P. distasonis*), a species within the genus *Parabacteroides* (strain 31_2, deposited as *Porphyromonas* sp. HM-169), was provided from the Biodefense and Emerging Infections Research Resources Repository (BEI Resources, NIAID, NIH) and maintained anaerobically in brain heart infusion (BHI) and chopped-meat broth. Growth assays followed an adaptation of Garcia-Bayona et al. ^73^ using M9 supplemented with 50 mg/L L-cysteine, 5 mg/L hemin, 2.5 µg/L vitamin K1, 2 mg/L FeSO4·7H2O, and 5 µg/L vitamin B12. Treatments included glucose (0.25% wt/vol), CON (0.25% wt/vol), or SCF (0.4% wt/vol). BHI served as a positive control; for negative controls, M9 with no carbon (no growth control), and blanks (no inoculum) for each treatment (media contamination control). Stock solutions were sterile-prepared, filter-sterilized (0.22 µm), and stored at 4 °C.

Three separate *P. distasonis* overnight cultures were inoculated into M9 solutions prepared in a 10 x 10 honeycomb plate (1:10 dilution; starting OD_600_ = 0.1). For each treatment, measurements were performed in technical triplicate across the 3 independent biological replicates. Growth was monitored with the Bioscreen-C system (600nm, every 30 min, for 25.5h, 37°C, shaking before/after each read) (**Supplemental Figure 3**). All samples reached the stationary phase by 14h; therefore, analyses were restricted to the first 14h. OD values were blank-corrected and growth rates (μ) calculated at 2 h intervals.

#### 16S-inferred functional potential

Predicted functional potential for the whole cohort was inferred from 16S data using *PICRUSt2* (v2.5.3) ^74^ with default parameters (*picrust2_pipeline.py*) ^74,75^. Predicted functions were summarized as KEGG Orthologs (KOs) and pathways. *KEGGREST* (v112.1) was used to extract carbohydrate metabolism pathways (map00010, 00030, 00500, 00620, 00650) and carbohydrate-active KOs (KO01176, 01223, 01210, 01190, 01187, 012348). Stratified KO outputs were used to quantify genus-level contributions to predicted functional differences across conditions.

#### Responder shotgun metagenomics

Participants from the whole cohort were classified based on their *Parabacteroides* response to SCF using 16S genus-level relative abundance change (Δ = POST-PRE during the SCF period). For an exploratory responder analysis, we selected a subset of six participants comprising the two extremes in *Parabacteroides* responses to the SCF intervention period: three participants that experienced the largest increases in *Parabacteroides* after the SCF intervention period (responders, n=3) and the three participants that experienced the smallest changes in *Parabacteroides* after the SCF intervention period (null or negative; non-responders, n=3). Shotgun metagenomic sequencing of the extracted fecal DNA from the 3 responders and 3 non-responders was conducted to assess species-level taxonomic profiles and exploratory beta-diversity and functional patterns by responder status (**Supplemental Figure 4**).

Shotgun metagenomic sequencing was performed at the W. M. Keck Center for Biotechnology (University of Illinois Urbana-Champaign). Libraries were pooled, quantified by qPCR, and sequenced on half of a 10B lane for 151 cycles from both ends of the fragments using a NovaSeq X Plus with V1.0 sequencing kits. FastQ files were demultiplexed (*bcl2fastq* v2.20), adapters (CTGTCTCTTATACACATCT) were trimmed from the 3’-end of the reads, and base quality scores were scored using Sanger encoding (*ASCII offset* of 33). Data pre-processing was performed on the University cluster (Carl R. Woese Institute for Genomic Biology, University of Illinois at Urbana-Champaign) using *KneadData* (v0.12.0) for quality control and host DNA removal (*KneadData* 20230405 release). *Trimmomatic* (v0.39) removed low-quality bases, *Tandem Repeat Finder* (TRF) removed noise from repetitive sequences, and *FastQC* (v0.11.9) generated quality reports.

Following Huttenhower BioBakery pipelines ^76–78^, taxonomic profiling was performed using *MetaPhlAn* (v4.0.6) ^77^, with *Bowtie2* (v2.5.0) aligning reads against the *ChocoPhlAnSGB*_202212 reference. Functional profiles were generated using *HUMAnN* (v3.7) ^76^ with the *ChocoPhlAn* nucleotide database (v201901b) and *UniRef90* protein database (Nov 2024). Profiles were merged (*merge_metaphlan_tables.py*) to produce relative abundance and read counts ^77^. Descriptive analyses compared the log fold change between responders and non-responder’s delta values, retaining gene families with >2-fold change-targeted filtering for carbohydrate metabolism-related terms yielded 217 candidate families. Carbohydrate-active enzymes (CAZymes) were annotated using the CAZy database ^79,80^ and *run_dbcan* (v4.0.0)^81^, incorporating *HMMER* (*dbCAN_HMMdb)*, *DIAMOND* (CAZy protein sequences), and *dbCAN-sub* for substrate prediction validated against PUL (Polysaccharide Utilization Loci) datasets. Reads were pre-processed (*Trim Galore* v0.6.0, *Kraken2* v2.1.1), assembled into contigs ≥1000 bp (*MEGAHIT* v1.2.9), and annotated with *Prokka* (v1.4) before CAZyme assignment. Clean reads were mapped back to coding sequences (*BWA* v0.7.17-r1188, *Samtools* v1.7, *Bedtools* v2.27.1), and abundances were normalized to transcripts per million (TPM) using *dbcan_utils* scripts ^81^. Workflow was conducted on a Linux computer (128 GB RAM) utilizing *Anaconda* (v23.7.3) environment.

### Statistical analyses

#### General approach

Target enrollment was determined using *a priori* power calculations (*G-Power* v3.1.9.2) ^82^, estimating that 36 participants would provide 80% power (β = 0.80) to detect a small effect size (*r* = 0.20) at α = 0.05 for Aims 1 and 2. Due to the exploratory nature of Aim 3, results should be interpreted as hypothesis-generating and to inform effect size estimates for future studies. To allow for attrition, 42 participants were recruited. Analyses were conducted in *RStudio* (v2025.05.0+496; *R* v4.4.3) ^83^. The significance threshold was set at α = 0.05 with false discovery rate (FDR) adjustments applied where relevant.

#### Data preprocessing and assumptions

Data distributions were checked using Shapiro-Wilk’s test (*stats* v4.4.3; *W* > 0.9), Q-Q plots (*ggplot2* v3.5.2) ^84^, skewness/kurtosis metrics (*DescTools* v0.99.6*, e1071* v2.6), and residuals inspected visually (*performance* v0.13.0). Extreme outliers (>3 SD; *rstatix* v0.7.20) were confirmed using a Bonferroni *outlierTest* (*car* v3.1-3), then winsorized ^85,86^. To meet model assumptions, BCFAs POMS, PANAS, and Godin were square root transformed; SCFAs were log-transformed. Data are reported as mean ± SEM unless otherwise noted.

#### Modeling strategy

Outcomes were analyzed using linear (*lme4* v1.1.37) or generalized (*glmmTMB* v1.1.11) mixed-effect models with treatment, time, and their interaction (treatment:time) as fixed effects, and participant ID as a random intercept [*Outcome ∼*

*Treatment*Time + (1|ID)*]. Order effects were tested and did not improve model fit, demonstrating adequate washout time, therefore excluded. Missing data were not imputed; the mixed-effect model provides valid estimates under the missing-at-random assumption ^60^. We reported *p*-values, estimates (β), standard errors (SE), and 95% confidence intervals (CI; *stats*) for the treatment effect – the level of response attributable to the treatment – estimated by the treatment:time interaction term, in the corresponding **Tables**. Fixed effects were evaluated using analysis of variance (ANOVA; *car* v3.1-3). When the treatment:time term met the significance threshold, we summarized simple-effects contrasts of estimated marginal means (*emmeans* v1.11.0), including between-treatment comparisons at endpoint with Benjamini-Hochberg (FDR) corrections. Baseline between-treatment contrasts were null across analyses. For outcomes with non-normal residuals or otherwise not meeting model assumptions, we used paired Wilcoxon signed-rank tests. *Aim 1*. All measures were analyzed with mixed-effects models (*lme4* v1.1.37). Flanker task measures (ACC, RT, and interference scores) were tested using treatment-by-time models with congruent and incongruent trial types evaluated separately. Two flanker ACC and two P3 amplitude outcomes were winsorized; *CANTAB* delta values were compared using paired Wilcoxon Signed-rank tests.

*Aim 2*. 16S analyses utilized rarefied counts for normalization. Beta diversity was assessed by PERMANOVA (*vegan* v2.6.10; 5,000 permutations) using participant ID as the permutation stratum, with homogeneity of dispersion (*vegan*, 5,000 permutations) tested in parallel. Differential abundance was assessed with negative binomial mixed-effect models (*glmmTMB* v1.1.11) comparing a null [*Taxa ∼ (1|ID)*] and full [*Taxa ∼ Treatment*Time + (1|ID)*] model by a likelihood ratio test across all included taxa ^87–89^. Alpha diversity and fecal metabolites were analyzed with mixed-effects models (*lme4* v1.1.37). Gastrointestinal symptoms were compared with a paired Wilcoxon Signed-rank test.

*Aim 3*. *In vitro* growth rates across treatments were modeled using a linear model (*lm*) with treatment, timepoint (every 2h), and the interaction as fixed effects, followed by pairwise contrasts ^90^. Predicted KO and pathway differences were compared with *MaAsLin2* (v1.18.0) ^91^ and *ggpicrust2* (v2.0.0) ^92^. Correlations between cognitive and microbial outcomes were assessed using Spearman’s rank correlation test (*stats* v4.4.3). Mediation analysis used baseline-adjusted models to (1) predict the mediator from the Treatment [Δ*Mediator ∼ Treatment + Mediator_Baseline_*] and (2) predict the outcome from both the Treatment and the Mediator [Δ*RT ∼ Treatment \** Δ*Mediator + RT_PRE_*]. Causal inference was estimated using nonparametric bootstrapping (*mediation* v4.5.0; 8,000 simulations) with sensitivity analyses. Reported are the average causal mediation effect (ACME), average direct effect (ADE), total effect, and proportion mediated. Moderation models included treatment, a centered-scaled moderator (center=“TRUE”, scale=“TRUE”), and its interaction [Δ*RT ∼ Treatment \** Δ*Moderator_Z-Scaled_*]. Participant ID was included as a random intercept, which caused model singularity; therefore, it was excluded. Comparisons were probed using simple-slopes (*interactions* v1.2.0) and provided confidence intervals (*emmeans* v1.11.0) at low (−1 SD), mean (0 SD), and high (+1 SD) changes of the moderator.

## Results

### Participant characteristics

The CONSORT diagram (**Figure 1**) summarizes recruitment, enrollment, and retention. Forty-two participants were randomized, of whom 41 completed the trial (**Table 1**). All participants completed at least one cognitive testing visit, including one who withdrew after the first intervention period (*n* = 1 CON). Per-protocol analyses included compliant participants who consumed >80% of intervention sachets during at least one intervention period (Aim 1; *n =* 38 CON, 39 SCF). Ten participants had unusable P3 amplitude data (*n*=3 CON; 1 at pre, 2 at post testing) and seven for the SCF (*n* = 2 PRE; *n* = 5 POST). Two participants could not provide fecal samples but completed the primary cognitive aims (Aim 2; *n =* 38 CON, 37 SCF).

**Figure 1.**
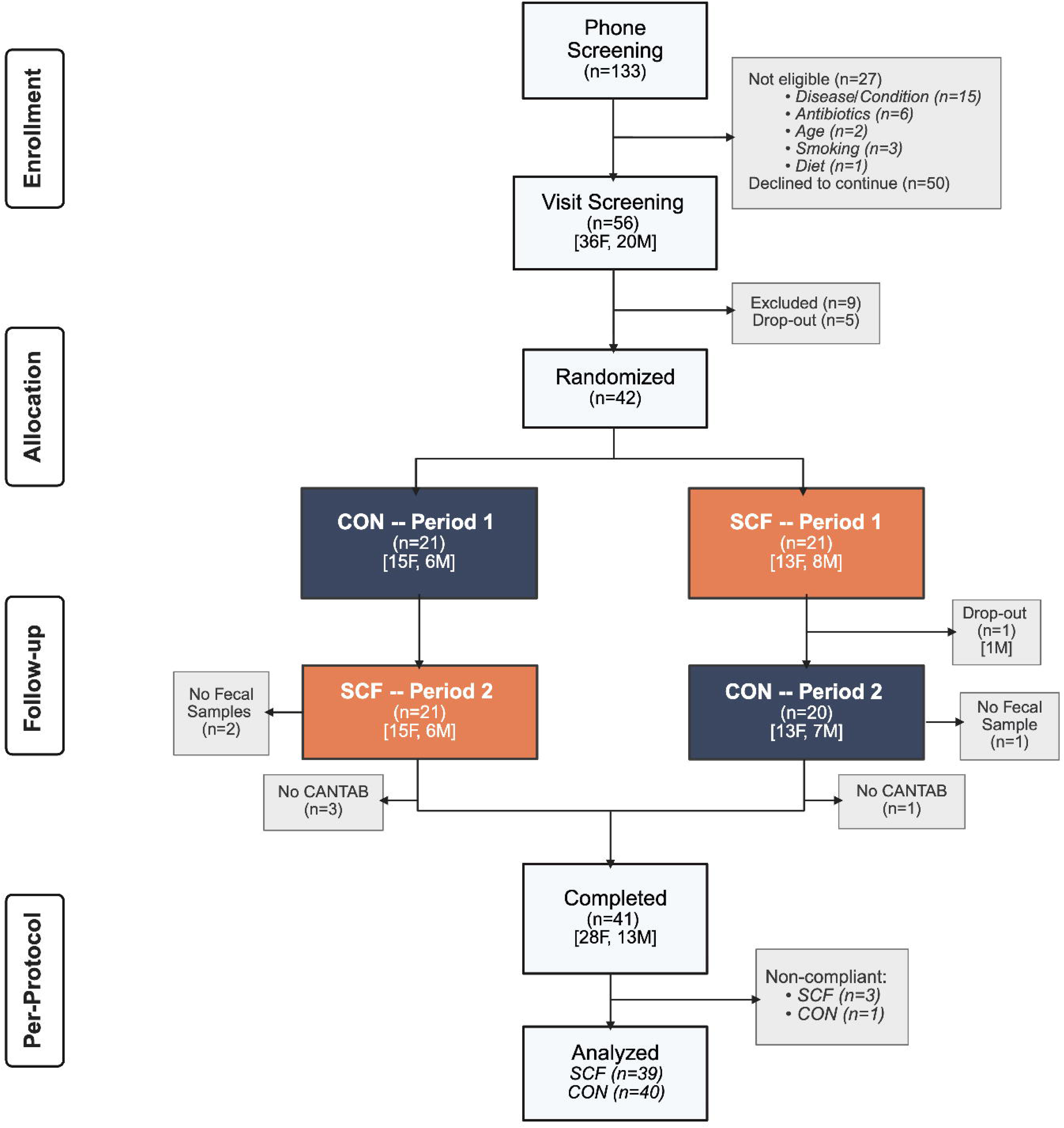
CONSORT flow diagram of recruitment, randomization, and retention. Participants were randomized (n=42) to start with either the maltodextrin control (CON) or soluble corn fiber (SCF) in Period 1, then crossed over to the alternate condition in Period 2 following washout. Boxes show the “*n*” indicating the number of participants at each stage and reasons for exclusion or missing data of the SCOPE study. (*Figure created using BioRender)*

#### Aim 1: Cognitive outcomes

### Executive function

Flanker task ACC remained high across the SCOPE study (≥ 92% scores), with ANOVA showing no treatment effect (treatment:time) during congruent (*P =* 0.42) or incongruent trial types (*P =* 0.53) (**Supplemental Figure 5**). RT exhibited a significant treatment effect during both trials (congruent *P =* 0.01, incongruent *P =* 0.01). At endpoint, RT was faster following SCF in both congruent (β = -9.8 ms, SE = 4.4, 95% CI: [-18.4, -1.2], FDR *P =* 0.01) and incongruent trials (β = -14.2 ms, SE = 4.4, 95% CI: [-22.8, -5.6], FDR *P =* 0.003) (**Figure 2**). P3 distributions were visualized (**Supplemental Figure 6**), no treatment effects were observed for P3 amplitudes (congruent *P =* 0.46; incongruent *P =* 0.33) or peak latency (congruent *P =* 0.63; incongruent *P =* 0.14). And no treatment effects were observed for interference scores of ACC (*P =* 0.10) or RT (*P =* 0.39).

**Figure 2.**
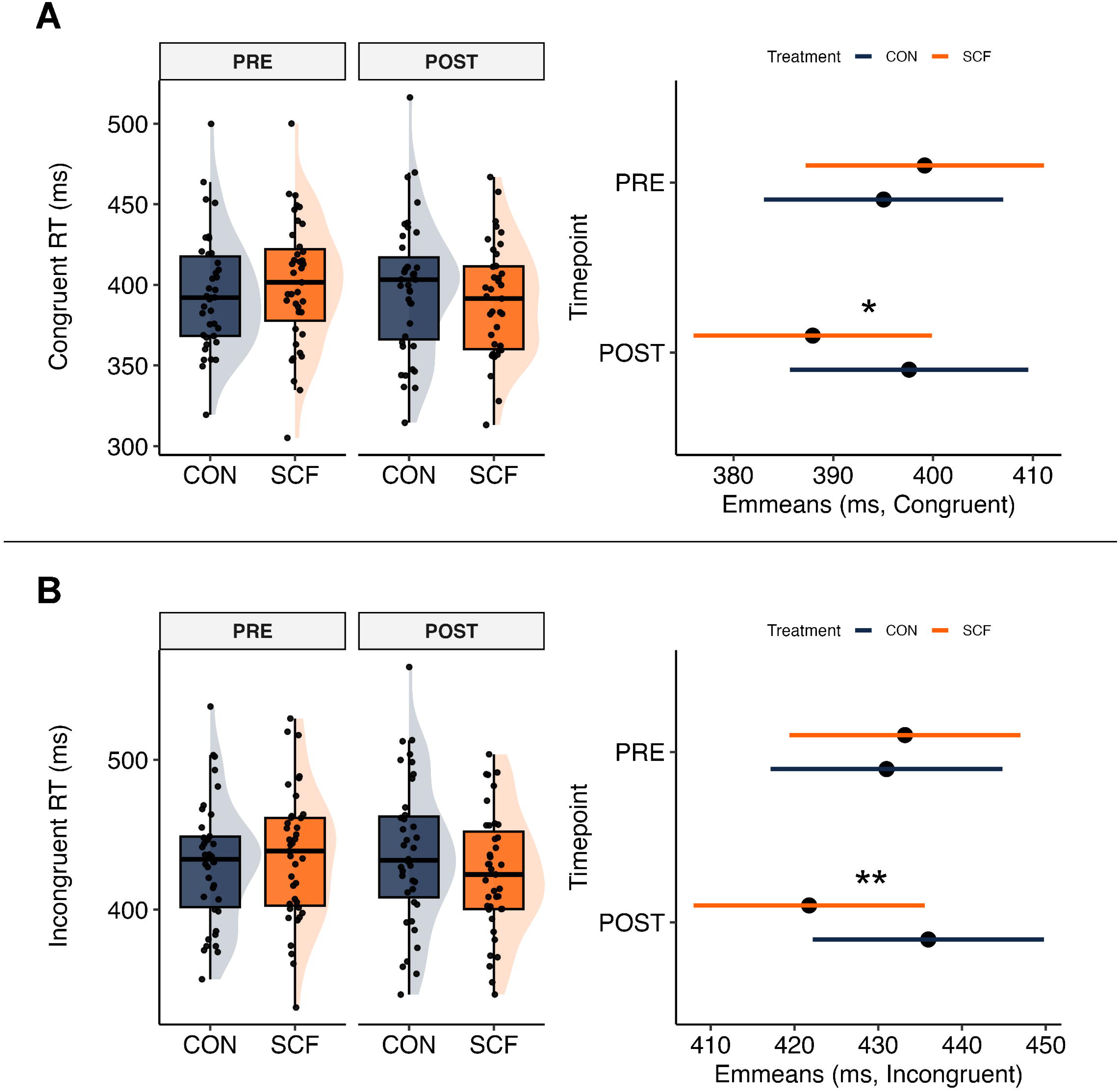
Soluble corn fiber (SCF) consumption improved flanker reaction times. Observed reaction times (RT, millisecond) are shown at both time points for the maltodextrin control (CON) and SCF intervention periods for (**A**) congruent trials and (**B**) incongruent trials. Right panels display model-estimated marginal means (coefficients and the 95% confidence intervals for both congruent and incongruent trials (FDR corrected pairwise contrasts; symbols shown in right panels). Statistical significance derived from pairwise comparisons. FDR-adjusted *p*-values annotated as follows: ** < 0.01; * < 0.05.

### Mood and Stress

No treatment effects were observed for mood subscales and total mood disturbance scores (*POMS*; all *P >* 0.53), relational memory performance (*iPosition*; all *P >* 0.45), affect scores (*PANAS*; all *P >* 0.45), and stress (*PSS*-10; *P* = 0.70) (**Supplemental Table 1**). Similarly, no treatment effects were observed across *CANTAB* measures (all *P >* 0.13) **(Supplemental Table 2-5**).

#### Aim 2: Gut Microbiota and Metabolite Outcomes

### 16S Fecal microbiota

Alpha diversity showed a treatment effect for observed richness (FDR *P =* 0.03), a marginal effect for Faith’s PD (FDR *P =* 0.09) and none for Shannon (FDR *P =* 0.26). Observed richness was reduced following SCF at endpoint (β = -29.9 counts, SE = 8.55, 95% CI [-47.1, - 12.6], FDR *P =* 0.01) (**Table 5**; **Supplemental Figure 7)**. Beta diversity showed a small but statistically significant treatment effect measured with Bray-Curtis (F = 0.81, R^2^ = 0.005, FDR *P <* 0.001), unweighted UniFrac (F = 0.47, R^2^ = 0.003, FDR *P =* 0.002), and weighted UniFrac (F = 1.07, R^2^ = 0.007, FDR *P <* 0.001) (**Supplemental Figure 8)**. Endpoint comparisons with each metric showed differences between SCF and CON for Bray-Curtis (F = 1.39, R^2^ = 0.019, FDR *P <* 0.001), unweighted UniFrac (F = 0.64, R^2^ = 0.010, FDR *P =* 0.005), and weighted UniFrac (F = 1.69, R^2^ = 0.023, FDR *P <* 0.001).

**Table 4.** Fecal volatile fatty acids across the intervention periods.

| | Maltodextrin | | Soluble Corn Fiber | | Treatment Effect (ΔSCF - ΔCON) | | | |
| --- | --- | --- | --- | --- | --- | --- | --- | --- |
|  | PRE | POST | PRE | POST |  |  |  |  |
| **Fermentation end-products**<br>(μmol/g DMB) | Mean ± SEM (*n*) | Mean ± SEM (*n*) | Mean ± SEM (*n*) | Mean ± SEM (*n*) | $\beta$ | SE | 95% CI | *P* |
| **<u>SCFAs</u>** |  |  |  |  |  |  |  |  |
| Butyrate | 43.3 ± 4.56 (38) | 43.9 ± 5.26 (37) | 38.8 ± 4.58 (37) | 40.1 ± 3.18 (37) | 0.10 | 0.13 | [-0.16, 0.37] | 0.44 |
| Acetate | 176. ± 13.9 (38) | 182. ± 13.5 (37) | 165. ± 13.2 (37) | 202. ± 14.9 (37) | 0.16 | 0.11 | [-0.04, 0.36] | 0.14 |
| Propionate | 48.1 ± 3.98 (38) | 50.8 ± 4.40 (37) | 47.5 ± 4.32 (37) | 55.6 ± 4.72 (37) | 0.14 | 0.10 | [-0.09, 0.36] | 0.23 |
| Total SCFAs | 267. ± 21.5 (38) | 273. ± 21.97 (37) | 255. ± 20.6 (37) | 298. ± 22.3 (37) | 0.15 | 0.11 | [-0.05, 0.36] | 0.15 |
| **<u>BCFAs</u>** |  |  |  |  |  |  |  |  |
| Valerate | 6.44 ± 0.59 (38) | 5.99 ± 0.47 (37) | 6.00 ± 0.47 (37) | 5.68 ± 0.48 (37) | -0.06 | 0.16 | [-0.36, 0.24] | 0.71 |
| Isobutyrate | 6.62 ± 0.54 (38) | 5.97 ± 0.39 (36) | 6.29 ± 0.52 (37) | 5.22 ± 0.33 (37) | -0.08 | 0.11 | [-0.30, 0.14] | 0.48 |
| Isovalerate | 9.49 ± 0.82 (38) | 8.30 ± 0.57 (36) | 8.66 ± 0.69 (37) | 7.35 ± 0.54 (37) | -0.04 | 0.14 | [-0.31, 0.22] | 0.75 |
| Total BCFAs | 22.6 ± 1.83 (38) | 20.4 ± 1.31 (37) | 21.0 ± 1.56 (37) | 18.3 ± 1.14 (37) | -0.03 | 0.20 | [-0.43, 0.36] | 0.87 |

Values are descriptive means ± standard error of the mean (SEM) and sample size (n) across each timepoint. Total SCFAs and BCFAs are pooled measures, within respective category, concentrations are on a dry matter basis (DMB). Linear mixed-effect models included participant ID as a random effect, and the fixed effects of treatment, time, and their interaction (Treatment Effect). Reported are the treatment-by-time interaction estimates ( $\beta$ , Standard Error (SE), 95% confidence interval (CI), *p*-values).

**Table 5.** Self-reported gastrointestinal symptom severity scores across the intervention.

|  | Maltodextrin |  | Soluble Corn Fiber |  | *P* |
| --- | --- | --- | --- | --- | --- |
|  | PRE | POST | PRE | POST |  |
| **GI Symptoms** | Median [IQR]<br>(*n*) | Median [IQR]<br>(*n*) | Median [IQR]<br>(*n*) | Median [IQR]<br>(*n*) |  |
| Gas | 1.57 [0.84] (39) | **1.43 [1.00] (39)** | 1.57 [0.97] (38) | **1.93 [1.00] (38)** | **0.01\*** |
| Cramping | 1.00 [0.00] (39) | 1.00 [0.00] (39) | 1.00 [0.00] (38) | 1.00 [0.14] (38) | 0.91 |
| Bloating | 1.00 [0.14] (39) | 1.00 [0.14] (39) | 1.00 [0.14] (38) | 1.00 [0.32] (38) | 0.05<sup>†</sup> |
| Nausea | 1.00 [0.00] (39) | 1.00 [0.00] (39) | 1.00 [0.00] (38) | 1.00 [0.00] (38) | 0.17 |
| Reflux | 1.00 [0.00] (39) | 1.00 [0.00] (39) | 1.00 [0.03] (38) | 1.00 [0.00] (38) | 0.84 |
| Rumbling | 1.00 [0.16] (39) | 1.00 [0.16] (39) | 1.00 [0.17] (38) | 1.00 [0.41] (38) | 0.68 |
| Burping | 1.00 [0.16] (39) | 1.00 [0.00] (39) | 1.00 [0.28] (38) | 1.00 [0.17] (38) | 0.70 |

Values are median [interquartile range (IQR)] and sample size (*n*) at each timepoint. Symptom were rated on a 4-point scale (1 = Absent, 2 = Mild, 3 = Moderate, 4 = Severe). Reported values include *p*-values from the paired Wilcoxon Signed-Rank testing the treatment effect on change over time using delta values. Significance threshold *p*: ‘\*’ <0.05, ‘†’ <0.1.

At the phylum level, none met the significance threshold (**Supplemental Table 6**). A total of 260 genera were detected, 128 passed filtering criteria (≥40% prevalence, >10 reads), and 14 met the significance threshold (all FDR *P <* 0.05); 11 were classified as low-abundance (<0.5%) or as a group of bacteria without taxonomical assignment (incertae_sedis) (**Supplemental Table 7)**. *Parabacteroides* enrichment was consistently observed across 90% of participants during the SCF arm (**Figure 3**), demonstrating a treatment effect (χ^2^ = 42.0, FDR *P <* 0.001), with a ∼4.1-fold increase following SCF compared to CON (β = 1.44 log, SE = 0.16, 95% CI [1.01, 1.88], FDR *P <* 0.001). *Fusicatenibacter* also showed a treatment effect (χ^2^ = 17.94, FDR *P <* 0.001) and ∼2.7-fold increase following SCF (β = 1.10 log, SE: 0.60, 95% CI: [0.59, 1.61], FDR *P* < 0.001). Similarly, *Dialister* had a treatment effect (χ^2^ = 7.25, FDR *P =* 0.02) and increased by ∼2.4-fold following SCF (β = 1.06 log, SE: 0.39, 95% CI: [0.29, 1.83], FDR *P =* 0.03) (**Supplemental** Figure 9).

**Figure 3.**
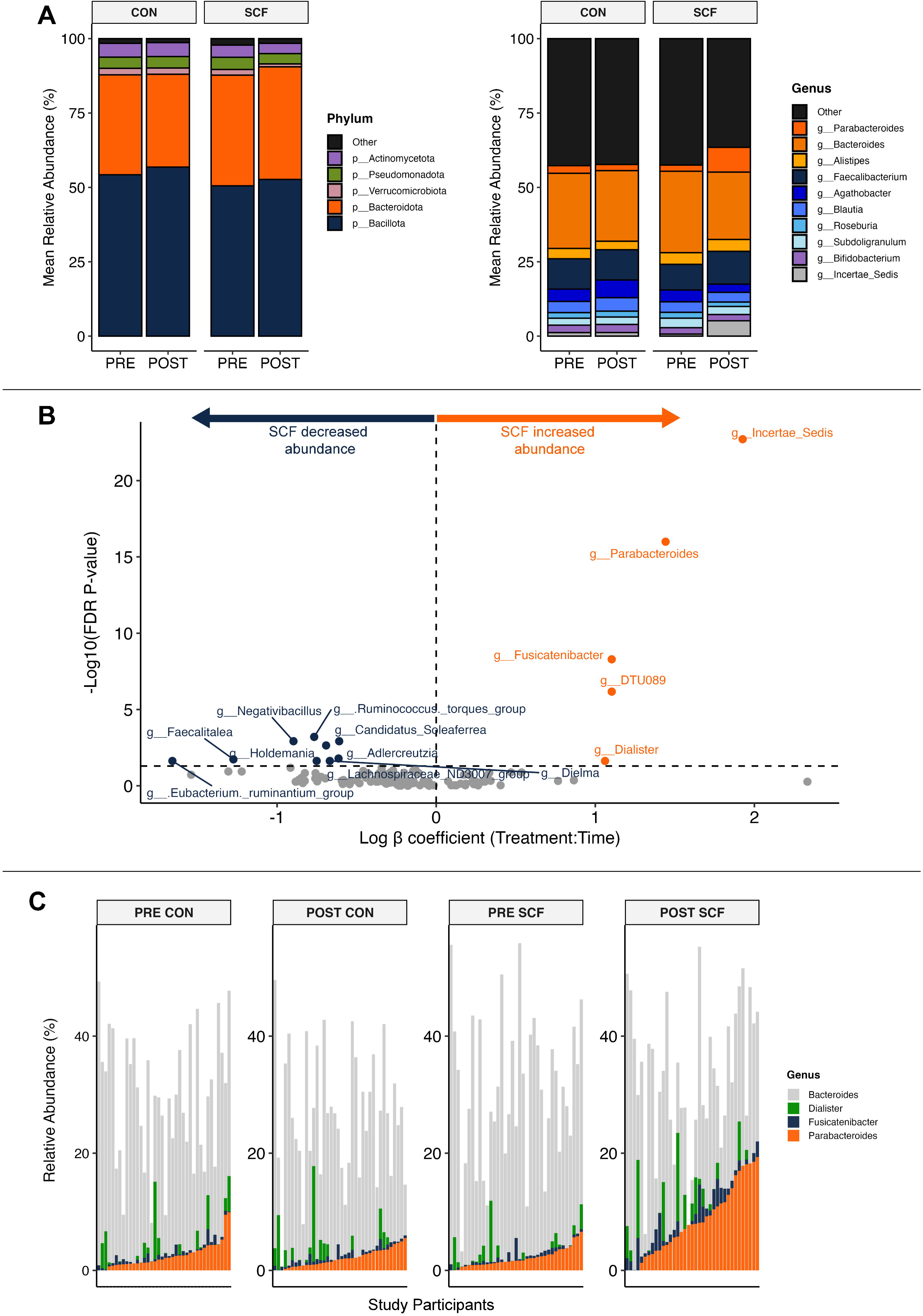
16S microbiota composition across condition and participants. (n=38 CON, n=37 SCF). (**A**) Mean relative abundance (% of sequence) of the top 5 for phyla (*left*) and top 10 for genera (*right*) at PRE and POST for each intervention. Genus colors correspond to their parent phylum. (**B**) Differential abundance of genera from a generalized linear mixed model showing the treatment effect estimates with effect size increasing from the center dashed vertical line. The y-axis corresponds to the significance of the genera, higher meaning more significant, with a horizontal dashed line indicating the FDR *P =* 0.05 threshold. Individual participant-sample relative abundances are shown as a (**C**) waterfall of the distribution of SCF-responsive features. Incertae sedis is group of bacteria without taxonomical assignment.

### Targeted metabolites

Fecal SCFAs (**Supplemental Figure 10**) and BCFAs did not statistically differ between treatments (all *P >* 0.14; **Table 4**). Pooled total indoles (indole, 2-methylindole, 3-methylindole, 7-methylindole), indole, pooled total phenols (4-methylphenol, 4-ethylphenol, phenol), 4-methylindole, and ammonia likewise did not show no treatment effect (all *P >* 0.32; **Supplemental Table 8**).

### Gastrointestinal symptoms

Symptom severity scores were generally absent to mild across conditions. No differences were observed between SCF and CON for changes in cramping, nausea, reflux, rumbling, or burping (all *P >* 0.17; **Table 5**). Flatulence (gas) severity scores increased following SCF (*P =* 0.01), although median severity remained <2 (mild) by endpoint (1.93 score). Bloating was marginally increased following SCF (*P =* 0.05), though median scores remained 1.00 (absent). No treatment effect was observed for self-reported feelings of consistency, ease of passage, bowel frequency, or in the measured fecal pH (all *P >* 0.22; **Supplemental Table 9**).

#### Aim 3: Exploratory Integrative and Functional Analyses

Aim 3 analyses were restricted to primary cognitive outcomes that differed by treatment (flanker RTs) and on the microbiota/metabolite features prioritized from Aim 2 based on either evidence of SCF-responsiveness (*Parabacteroides*, *Fusicatenibacter*, and *Dialister*) or a consistent directional trend with biological plausibility (acetate, propionate).

### Correlation analyses

Spearman correlations at the SCF endpoint showed marginal inverse associations between congruent RT and acetate (ρ = -0.33, FDR *P =* 0.09) and propionate (ρ = -0.36, FDR *P =* 0.09). *Parabacteroides* was marginally associated with acetate (ρ = 0.34, FDR *P =* 0.09). No taxa were associated with congruent RT (all FDR *P >* 0.14). For incongruent trials, no significant correlations were detected with any candidate (all ρ < 0.23 and FDR *P >* 0.30) **(Supplementary Figure 11)**.

### Mediation analyses

The proportion mediated was estimated at ≤10.0% across candidates for congruent trials and <4.1% for incongruent trials (**Table 6**). However, across candidates, indirect effects (ACME) were not detected for either congruency (all *p* ≥ 0.19), indicating no evidence of mediation.

**Table 6.** Mediation of SCF-related changes in Flanker reaction times through fecal SCFAs and 16S genera.

| Nonparametric Bootstrap Confidence Intervals with Percentile |  |  |  |  |  |
| --- | --- | --- | --- | --- | --- |
| Mediator | Flanker Task | Effect | $\beta$ | [95% CI] | *P* |
| *Acetate* | Congruent RT | ACME | -0.57 | [-2.51, 0.87] | 0.48 |
|  |  | ADE | -13.42 | [-23.7, -3.23] | 0.01* |
|  |  | Total effect | -13.99 | [-24.4, -14.0] | 0.009** |
|  |  | Proportion Mediated | 4.09% | [-0.08, 0.21] | 0.48 |
|  | Incongruent RT | ACME | -0.44 | [-2.87, 1.61] | 0.72 |
|  |  | ADE | -17.03 | [-29.4, -4.38] | 0.008** |
|  |  | Total effect | -17.47 | [-30.1, -4.63] | 0.007** |
|  |  | Proportion Mediated | 2.51% | [-0.11, 0.19] | 0.72 |
| *Propionate* | Congruent RT | ACME | -0.64 | [-2.39, 0.81] | 0.43 |
|  |  | ADE | -13.39 | [-23.9, -3.30] | 0.01* |
|  |  | Total effect | -14.03 | [-24.4, -3.78] | 0.008** |
|  |  | Proportion Mediated | 4.57% | [-0.28, 0.08] | 0.43 |
|  | Incongruent RT | ACME | -0.38 | [-4.32, 0.78] | 0.22 |
|  |  | ADE | -17.11 | [-23.0, -2.36] | 0.03* |
|  |  | Total effect | -17.49 | [-24.3, -3.77] | 0.01* |
|  |  | Proportion Mediated | 2.22% | [-0.11, 0.54] | 0.23 |
| *Parabacteroides* | Congruent RT | ACME | -0.15 | [-9.31, 4.27] | 0.93 |
|  |  | ADE | -13.85 | [-25.9, 0.39] | 0.06 |
|  |  | Total effect | -14.00 | [-24.3, -3.83] | 0.008** |
|  |  | Proportion Mediated | 1.06% | [-0.36, 1.02] | 0.93 |
|  | Incongruent RT | ACME | -0.48 | [-12.64, 4.53] | 0.86 |
|  |  | ADE | -17.00 | [-30.5, 0.20] | 0.03* |
|  |  | Total effect | -17.48 | [-30.0, -4.80] | 0.01* |
|  |  | Proportion Mediated | 2.75% | [-0.34, 0.96] | 0.86 |
| *Fusicatenibacter* | Congruent RT | ACME | -1.40 | [-4.98, 0.65] | 0.19 |
|  |  | ADE | -12.63 | [-23.0, -2.28] | 0.02* |
|  |  | Total effect | -14.03 | [-24.3, -3.84] | 0.007** |
|  |  | Proportion Mediated | 10.0% | [-0.06, 0.50] | 0.19 |
|  | Incongruent RT | ACME | -0.72 | [-4.98, 2.16] | 0.59 |
|  |  | ADE | -16.75 | [-29.1, -3.85] | 0.01* |
|  |  | Total effect | -17.47 | [-30.0, -4.79] | 0.007** |
|  |  | Proportion Mediated | 4.1% | [-0.18, 0.35] | 0.59 |
| *Dialister* | Congruent RT | ACME | 0.44 | [-0.89, 3.47] | 0.52 |
|  |  | ADE | -14.44 | [-25.4, -4.22] | 0.006** |
|  |  | Total effect | -14.00 | [-24.2, -3.75] | 0.007** |
|  |  | Proportion Mediated | -3.15% | [-0.33, 0.08] | 0.53 |
|  | Incongruent RT | ACME | 0.67 | [-1.26, 4.95] | 0.52 |
|  |  | ADE | -18.15 | [-31.8, -5.12] | 0.007** |
|  |  | Total effect | -17.49 | [-30.0, -4.70] | 0.007** |
|  |  | Proportion Mediated | -3.8% | [-0.35, 0.11] | 0.53 |

Mediation analysis ( $n = 37$ ) was conducted using nonparametric bootstrapped linear regression models (R; *mediation*) to estimate the average causal mediation effect (ACME), average direct effect (ADE), and total effects of treatment on changes in reaction times (RT). Reported are the model estimates ( $\beta$ ), Standard Error (SE), 95% confidence interval (CI). Proportion mediated indicates the percentage of the total effect explained by the ACME pathway. Significance threshold $p$ : ‘\*\*\*’ $<0.001$ , ‘\*\*’ $<0.01$ , ‘\*’ $<0.05$ , ‘†’ $<0.1$ .

### Moderation analyses

Moderation models provided no evidence that candidate changes modified the treatment effect on RT (all *P >* 0.49), except for *Parabacteroides*, which showed a moderation effect for congruent (*P =* 0.05) and incongruent RTs (*P =* 0.04). Adding *Parabacteroides* increased explained variance from ΔR^2^ = 0.07 (base model) to ΔR^2^ = 0.12 (adjusted), whereas other candidates contributed <1% (**Table 7**).

**Table 7.** Moderation of the SCF effect on change in flanker reaction time by change in 16S genera and fecal metabolites.

| Moderator | Flanker Task | Treatment x Moderator (Model Summary) |  |  |  |  |  |  |
| --- | --- | --- | --- | --- | --- | --- | --- | --- |
| | | $\Delta R^2$ | Adj. $\Delta R^2$ | $\beta$ | SE | t | [95% CI] | P |
| Acetate | Congruent RT | 0.11 | 0.07 | 3.13 | 5.62 | 0.56 | [-8.08, 14.3] | 0.58 |
|  | Incongruent RT | 0.10 | 0.06 | 2.21 | 6.80 | 0.33 | [-11.4, 15.8] | 0.75 |
| Propionate | Congruent RT | 0.11 | 0.07 | 0.43 | 5.60 | 0.08 | [-10.76, 11.6] | 0.94 |
|  | Incongruent RT | 0.10 | 0.06 | 0.40 | 6.80 | 0.06 | [-13.17, 14.0] | 0.95 |
| *Parabacteroides* | Congruent RT | **0.16** | 0.12 | **-19.8** | 10.12 | -1.95 | [-40.0, 0.43] | **0.05\*** |
|  | Incongruent RT | **0.15** | 0.12 | **-25.1** | 12.24 | -2.05 | [-49.5, 0.63] | **0.04\*** |
| *Fusicatenibacter* | Congruent RT | 0.10 | 0.06 | -1.31 | 13.48 | -0.10 | [-28.2, 25.6] | 0.92 |
|  | Incongruent RT | 0.11 | 0.07 | 11.49 | 16.20 | 0.71 | [-20.9, 43.8] | 0.49 |
| *Dialister* | Congruent RT | 0.11 | 0.07 | -4.49 | 7.39 | -0.61 | [-19.2, 10.3] | 0.55 |
|  | Incongruent RT | 0.10 | 0.06 | 0.23 | 8.98 | 0.03 | [-17.7, 18.2] | 0.98 |

Moderation analysis ( $n = 37$ ) was tested using linear regression models testing if changes in moderators explain variability in flanker reaction times (RT). Each model includes treatment, $\Delta$ moderator, and its interaction. Reported are the treatment-by-moderator estimates ( $\beta$ , Standard Error (SE), 95% confidence interval (CI), $p$ -values). The $\Delta R^2$ and adjusted $\Delta R^2$ columns show the variance explained by the full model. Statistical threshold for $p$ : ‘\*\*\*’ <0.001, ‘\*\*’ <0.01, ‘\*’ <0.05, ‘†’ <0.1.

For congruent trials, SCF-associated RT improvement depended on *Parabacteroides* change (z-scored; 0 SD = average change): RT was faster when *Parabacteroides* changes were small (−1SD: β = -29.5 ms, SE = 8.76, *t* = -3.37, 95% CI: [-47.0, -12.03], *P <* 0.001), but not when *Parabacteroides* change was near the mean (0 SD; β = -9.7 ms, SE = 8.27, *t* = -1.18, 95% CI: [-26.2, 6.8], *P =* 0.24) or much larger (+1 SD: β = 10.0 ms, SE = 16.27, *t* = 0.62, 95% CI: [-22.5, 42.5], *P =* 0.54). For incongruent trials, the treatment effect on RT similarly depended on *Parabacteroides* change. RT was faster when changes were small (β = -32.4 ms, SE = 10.59, *t* = - 3.06, 95% CI: [-53.5, -11.2], *P <* 0.001), but not when change was near the mean (β = -7.30 ms, SE = 10.0, *t* = -0.73, 95% CI: [-27.3, 12.7], *P =* 0.47) or much larger (β = 17.8 ms, SE = 19.68, *t* = 0.90, 95% CI: [-21.5, 57.0], *P =* 0.37) (**Figure 4**).

**Figure 4.**
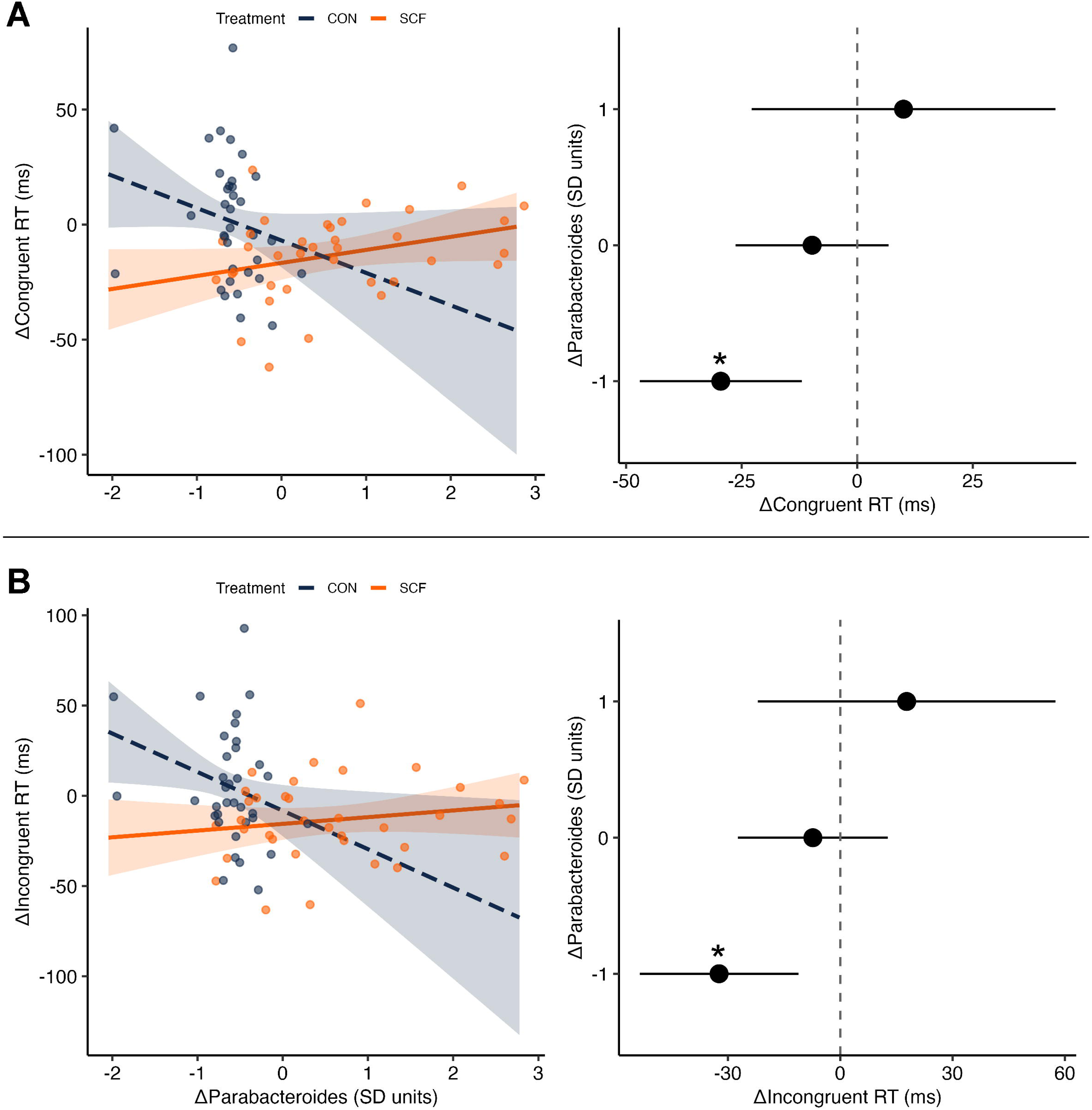
Interaction between 16S-derived Δ*Parabacteroides* and the SCF effect on flanker RT. Panels show moderation results for (**A**) for congruent and (**B**) incongruent trials. The outcome is ΔRT (POST-PRE, ms), where negative values indicate faster RT. The moderator is Δ*Parabacteroides* (POST-PRE), mean-centered and z-scaled (negative values reflect smaller-than-average changes). Left panels show observed data and model-predicted simple slopes for CON (blue) and SCF (orange); vertical reference lines indicate -1 SD (standard deviation), average (0 SD), and +1 SD. Right panels show estimated SCF-CON emmeans contrasts in ΔRT at -1,0, and +1SD of Δ*Parabacteroides*; the vertical line at 0 indicates no difference. Statistical significance threshold *p* * <0.05.

### In vitro culturing of P. distasonis

*P. distasonis* was confirmed to grow with SCF in isolation displaying a distinct growth kinetics with significant media effect (*P <* 0.001), time (*P <* 0.001), and interaction (*P <* 0.001). Growth on glucose and CON showed a short lag phase (<1h), and a rapid log phase reaching stationary phase within 5-6h (glucose: OD_600_ = 0.59, 5.5h; CON: OD_600_ = 0.54, 5h). Growth on SCF exhibited a two-phase diauxic growth pattern: a slower and smaller initial phase reaching OD_600_ = 0.31 at 6h, followed by a ∼2h lag, then a second log phase that peaked at OD_600_ = 0.61 by 14h. *Post-hoc* comparisons showed that SCF had significantly lower growth rates at 2h (all FDR *P <* 0.001) and 4h (vs. glucose, FDR *P =* 0.03; vs. CON, FDR *P =* 0.01) relative to controls. By 6h, no differences were detected across treatments. At 8-10h, SCF had higher growth rates compared to controls (all FDR *P <* 0.001), and remained higher at 12h (vs. glucose, FDR *P =* 0.002). By 14h, growth stabilized with no differences detected between treatments (**Figure 5)**.

**Figure 5.**
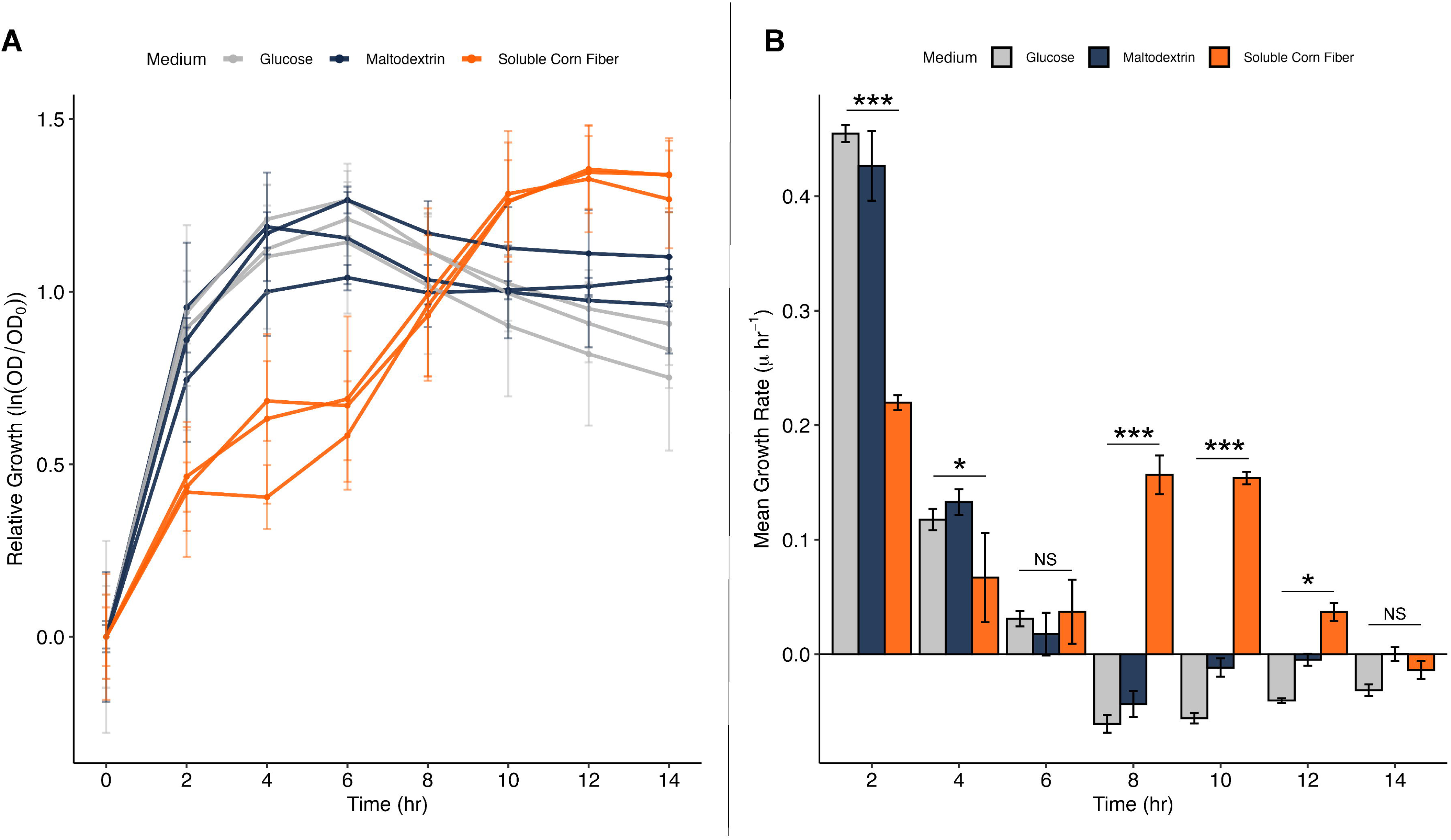
Growth rates of *Parabacteroides distasonis* cultured in M9 supplement with SCF compared to controls. The relative log growth curve of *P. distasonis* (**A**) in M9 minimal medium supplemented with different carbohydrate sources. OD_0_ represents the initial optical density (OD_600_) at time zero. Each curve represents a mean biological replicate (N = 3), and error bars indicate propagated standard deviations. The mean growth rate (**B**), calculated as the slope of ln(OD) over sequential 2-hour intervals, cut off at 10-hrs. Bars represent average growth rate across replicates. Significance threshold *p*: ‘***’ < 0.001, ‘**’ < 0.01, ‘*’ < 0.05, ‘†’ < 0.1, ‘NS’ > 0.1.

### 16S-Predicted functional potential analyses

Predicted microbial pathway profiles from the full cohort indicated higher *N*-glycan biosynthesis and α-linolenic acid metabolism pathways following SCF at endpoint compared to control (all FDR *P <* 0.01). Predicted enzyme profiles (ECs) identified 23 functions differing by treatment at endpoint (all FDR *P <* 0.05) (**Supplemental Figure 12**). MaAsLin2 detected 2,701 carbohydrate-metabolism KOs associated with SCF at endpoint (FDR *P <* 0.05). Among the top SCF-associated KOs, 18 showed contribution by *Parabacteroides* (including glycan degradation and complex carbohydrate utilization) and two by *Fusicatenibacter* (**Supplemental Figure 13**).

### Responder shotgun metagenomic analyses

Metagenomic profile comparisons between responder vs. non-responded indicated no differences in alpha diversity (all *P >* 0.09) or beta diversity. Responder status explained ∼17% of variance using Bray-Curtis PERMANOVA (R^2^ = 0.167, *P =* 0.05), with no main effect of time (R^2^ = 0.083, *P =* 0.44) and no difference in the interaction (R^2^ = 0.079, *P =* 0.49) (**Figure 6)**. Metagenomic genus-level profiles focused on the treatment-responsive genera identified in 16S analyses and confirmed that *Parabacteroides* was enriched in responders (Δ = 13.6%) and slightly reduced in non-responders (Δ = -0.4%), consistent with 16S results. At the species level, *P. distasonis* increased in responders (Δ = 10.9%) and remained non-detectable in non-responders, while *P. merdae* also increased in responders (Δ = 2.7%) and decreased in non-responders (Δ = - 0.4%). *F. saccharivorans* negligibly increased in responders (Δ = 0.1%), compared to a larger increase in non-responders (Δ = 2.2%). *D. hominis* (Δ = 1.4%) and *D. invisus* (0.3%) increased in non-responder, while only *D. invisus* was detected and decreased in responders (Δ = -0.3%).

**Figure 6.**
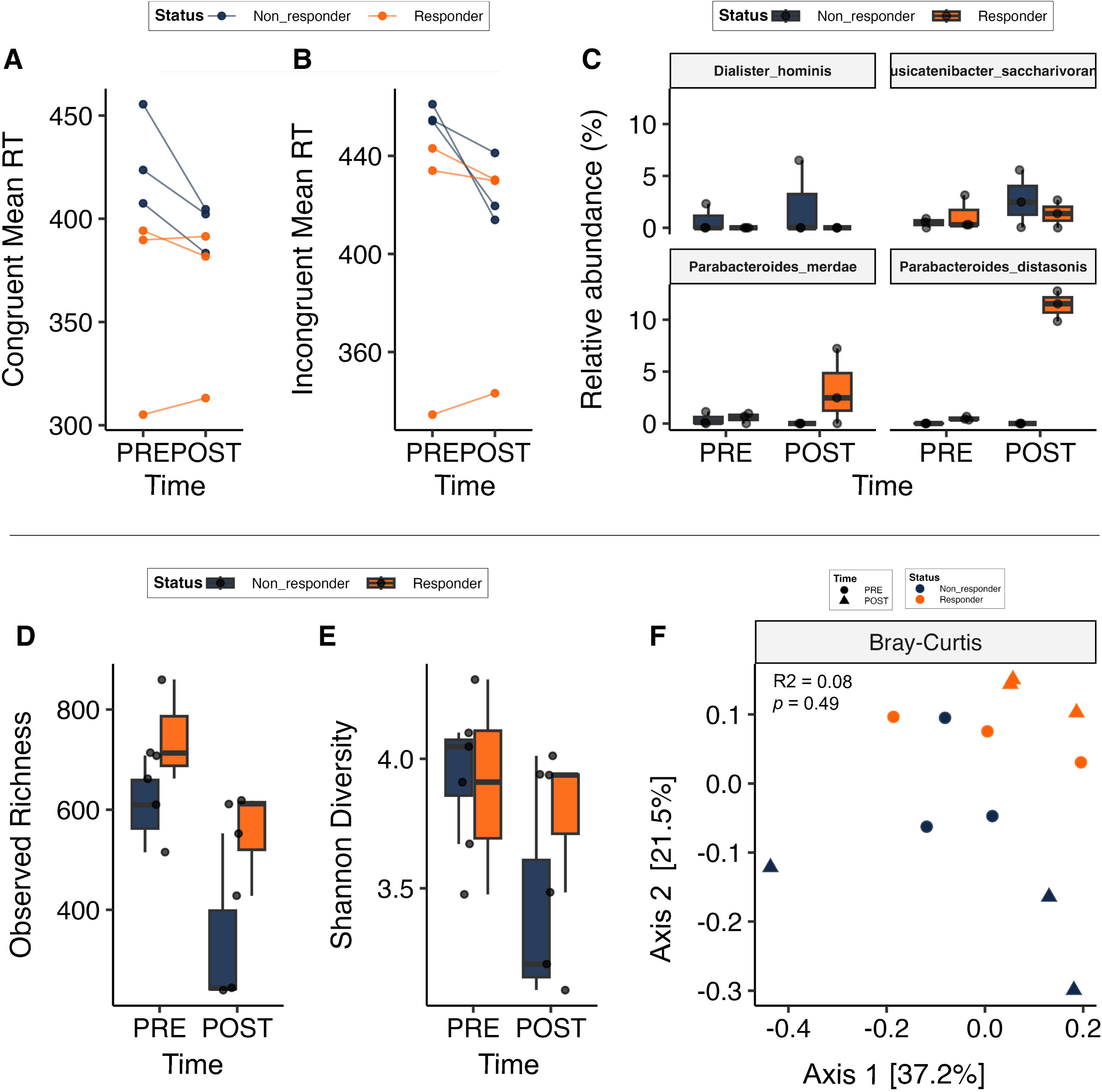
Exploratory responder analysis during the SCF period with MetaPhlAn4. N = 6. Responders (n=3) were the participants with the largest *Parabacteroides* increase after SCF consumption, and non-responders (n=3) were the three with the smallest change (null/negative). The cognitive outcomes from the Flanker task demonstrate the responders (**A**) congruent and (**B**) incongruent reaction times (RT). Shotgun sequencing and MetaPhlan4 yielded (**C**) species resolution of microbial hits from 16S genus analysis. The bottom row demonstrates the diversity metrics for Alpha (**D**) richness and (**E**) Shannon evenness, and the principal coordinate analysis of (**F**) Beta diversity is shown using Bray-Curtis with adonis2 PERMANOVA values.

Functional analyses with *HUMAnN* showed an increase in carbohydrate metabolism-related KOs among responders (UDP-glucose 6-dehydrogenase, α-amylases, hexokinase), while non-responders demonstrated a lack of presence in these KOs (**Supplemental Figure 14**). Genus-(**Supplemental Figure 15)** and species-level time comparisons (**Figure 10**) in responders indicated increased contributions from *P. distasonis* to multiple carbohydrate-active gene families, including glycoside hydrolases (GHs), α-amylases, and hexokinase. CAZy annotations provided further resolution of GH families and subfamilies between groups. Of 228 GH families detected, those with positive log fold changes in responders were isolated (**Supplemental Figure 16**). Several GH families were enriched (GH43_8, GH43_33, GH25, GH24, GH179, GH13_9, GH130_2, GH12) and positively correlated with *Parabacteroides* abundances (**Supplemental Figure 17**).

Responders demonstrated faster RT across the SCF intervention for both congruent (Δ = - 0.90ms) and incongruent (Δ = 2.7ms) trials. In contrast, non-responders exhibited slower baseline RT but demonstrated larger SCF-induced improvements (congruent Δ = -32.1ms, incongruent Δ = -31.8ms). Parallel changes were observed in SCFAs; responders had higher baseline SCFA levels and was increased in both groups for acetate (responders Δ = 66.4 vs. non-responders Δ = 71.6 µmol/g DMB) and propionate (responders Δ = 20.9 vs. non-responders Δ = 28.6 µmol/g µmol/g DMB). (**Table 8**).

**Table 8.** Responder vs. non-responder descriptive summary during the SCF intervention period.

|  | Non-Responder |  | Responder |  |
| --- | --- | --- | --- | --- |
|  | PRE | POST | PRE | POST |
| Pilot Analyses | Mean ± SD (n) | Mean ± SD (n) | Mean ± SD (n) | Mean ± SD (n) |
| **Genera** (% of Sequences) |  |  |  |  |
| *Parabacteroides* | 0.43 ± 0.64 (3) | ND (3) | 1.04 ± 0.33 (3) | 14.61 ± 2.82 (3) |
| *Dialister* | 1.50 ± 0.79 (3) | 3.18 ± 3.25 (3) | 0.25 ± 0.43 (3) | ND (3) |
| *Fusicatenibacter* | 0.48 ± 0.45 (3) | 2.69 ± 2.77 (3) | 1.24 ± 1.65 (3) | 1.35 ± 1.34 (3) |
| **Species** (% of Sequences) |  |  |  |  |
| *P. distasonis* | 0.01 ± 0.01 (3) | ND (3) | 0.49 ± 0.17 (3) | 11.38 ± 1.47 (3) |
| *P. merdae* | 0.41 ± 0.63 (3) | ND (3) | 0.55 ± 0.50 (3) | 3.23 ± 3.67 (3) |
| *P. faecis* | 0.01 ± 0.01 (3) | ND (3) | ND (3) | ND (3) |
| *D. invisus* | 0.73 ± 0.73 (3) | 1.00 ± 1.74 (3) | 0.25 ± 0.43 (3) | ND (3) |
| *D. hominis* | 0.77 ± 1.34 (3) | 2.17 ± 3.75 (3) | ND (3) | ND (3) |
| *F. saccharivorans* | 0.48 ± 0.45 (3) | 2.69 ± 2.77 (3) | 1.24 ± 1.65 (3) | 1.35 ± 1.34 (3) |
| **Flanker Reaction Time** (ms) |  |  |  |  |
| Congruent | 428.9 ± 24.4 (3) | 396.7 ± 11.6 (3) | 363.0 ± 50.2 (3) | 362.1 ± 42.7 (3) |
| Incongruent | 456.6 ± 3.9 (3) | 424.9 ± 14.4 (3) | 403.9 ± 60.2 (3) | 401.1 ± 50.2 (3) |
| **SCFAs** (μmol/g DMB) |  |  |  |  |
| Acetate | 153.8 ± 60.9 (3) | 225.4 ± 88.9 (3) | 180.2 ± 95.1 (3) | 246.6 ± 59.1 (3) |
| Propionate | 28.47 ± 12.2 (3) | 57.02 ± 23.7 (3) | 51.89 ± 23.6 (3) | 72.81 ± 27.2 (3) |
| **Demographics** |  |  |  |  |
| Age (year) | 53.7 ± 7.2 (3) |  | 54.3 ± 2.5 (3) |  |

Responders (n=3) and non-responders (n=3) following the SCF intervention. Responders were the three participants with the largest increase in *Parabacteroides* (16S) following SCF; non-responders were the three participants with the smallest change following SCF (total N = 6 participants). Data are presented as mean ± standard deviation (SD) and sample size (n). Genus and species relative abundances (%) were derived from shotgun metagenomic profiling using MetaPhlAn4. Species initials correspond as genus followed by species name: *P* = *Parabacteroides*, *D* = *Dialister*, and *F* = *Fusicatenibacter*. ND = not detected (no reads assigned at the reported taxonomic level.)

## Discussion

In this randomized, double-blind, placebo-controlled, crossover trial, consuming SCF daily for 4-weeks improved attentional inhibition, reflected by faster RT without compromising accuracy on both congruent and incongruent flanker task trial types compared to control. Effects were domain-specific to attentional inhibition, with no changes observed in relational memory, rule acquisition and reversal, set shifting, sustained attention, response inhibition, or spatial working memory observed (*Aim 1*). SCF consumption also shifted fecal microbial diversity, selectively enriched *Parabacteroides*, numerically increased fecal acetate concentrations, and produced absent-to-mild GI symptoms (*Aim 2*). SCF consumption had no effect on P3 amplitude or latency, mood, or stress. Exploratory analyses did not identify any mediators but moderation analyses suggested that SCF-related RT improvements varied by the magnitude of *Parabacteroides* change. Consistent with 16S results identifying *Parabacteroides* as SCF-responsive at the genus level, shotgun metagenomics identified *P. distasonis* as the primary SCF-responsive species, while *in vitro* culturing independently confirmed its capacity to metabolize SCF independent of cross-feeding (*Aim 3*).

Attentional inhibition is a core domain of cognitive control that can naturally and insidiously decline with aging ^31^, underscoring the need for dietary strategies to preserve healthy cognitive aging. Few clinical trials have assessed the impact of dietary fiber consumption on cognition while integrating microbiome outcomes. The present work demonstrated that SCF consumption selectively supported attentional inhibition, reflected by faster RT in congruent and incongruent flanker trials, with no changes in other cognitive outcomes. This domain-specific pattern reinforces the concept that gut-brain communication can influence higher-order cognitive processes. Another randomized trial among 84 adults with overweight and obesity reported improved accuracy, but no change in RT, during a modified flanker task following 12 weeks of daily avocado consumption (12.8-16 grams fiber) relative to a control providing 3.2 – 4 grams fiber daily ^93^. Notably, avocado provides a mixed plant cell-wall fiber profile that is predominantly insoluble (cellulose/hemicellulose) with soluble pectin (along with other nutrients) ^94^, whereas SCF is a digestion-resistant maltodextrin dextrinized to produce a mixture of ɑ-linkages (ɑ-1,2, - 1,3, -1,4, and -1,6) ^18,19^, which may yield distinct fermentation kinetics and downstream signaling. Specific to the microbiota, the avocado intervention increased ɑ-diversity and enriched *Faecalibacterium, Lachnospira*, and *Alistipes* and resulted in 18% greater fecal acetate, concentrations due to fiber-rich avocado consumption ^95^. Another nutrition trial reported mood benefits with 16 g/day inulin for 12 weeks versus 16 g maltodextrin among adults with obesity, particularly among individuals exhibiting a specific microbial signature, however, no effects were observed on inhibitory control, working memory, or cognitive flexibility regardless of changes in the gut microbiota ^39^. Inulin is a β(2,1)-linked fructan (fructose polymer), whereas SCF is glucose based ^96,97^. Other lifestyle interventions, such as increased physical activity, have been shown to improve flanker RT after a single exercise session, with one randomized trial among 106 healthy-weight adults showing improvements in both congruent (458 ms vs 475 ms) and incongruent (528 ms vs 547 ms) paradigms immediately following 30 min of high-intensity aerobic exercise compared with seated rest ^98^. Collectively, these studies suggest that cognition effects are domain-and context-specific, shaped by factors such as fiber type, population characteristics, physical activity status, and intervention duration, which complicates direct cross-study comparisons but underscores nutrition as a potential modifiable factor for cognitive health. However, the fiber type (e.g., SCF, pectin), host and microbiome characteristics, and exposure duration needed to exact cognitive benefits is yet to be delineated, highlighting the need for additional robust clinical trials. Our work demonstrated that *Parabacteroides* was enriched following SCF consumption, which is consistent with previous SCF supplementation trials. Specifically, a randomized dose-responsive trial with free-living pubertal females (0, 10, and 20g fiber/d, 4 weeks per dose), a randomized double-blind trial with healthy adults (20 g/d; 10 days per period), and a randomized double-blind trial with only healthy adult men (21 g/d; 21 days per period) all reported enrichment of *Parabacteroides* ^21,99,100^. In the present study, SCF consumption reduced observed richness at endpoint, which has been previously reported with fermentable fibers when selective utilization leads to an increase of a narrower set of taxa and a more dominated community structure, without necessarily indicating an adverse effect ^101^ as shown by its consistent role as a fermentable and generally well tolerated fiber ^97^.

The convergence of 16S with the full cohort and shotgun metagenomes on responder subset supports the robustness of the *Parabacteroides* enrichment across sequencing approaches. In responders, *P. distasonis* abundance was inversely related to *Bacteroides vulgatus*, suggesting possible ecological competition under SCF exposure. Predicted functional profiles and shotgun metagenomics indicated increased carbohydrate metabolism pathways and glycan degradation capacity, with CAZy highlighting an increased in *Parabacteroides*-associated GHs (GH35, GH43, and GH73) ^102,103^. *P. distasonis*-specific GHs (GH144 and GH13) involved in the hydrolysis α-glycosidic bonds were also detected ^102,103^. CAZymes serving similar functions are typically organized within PULs, gene clusters that coordinate sensing, import, and degradation of dietary glycans ^104^. The enrichment of these CAZymes likely reflects an activation of SCF-targeting PULs, enabling *P. distasonis* to maintain a competitive niche in the large intestine. Our *in vitro* experiment supported this metabolic capacity, showing that *P. distasonis*, with no prior SCF exposure, fermented SCF in a diauxic pattern, consistent with the mixture of fiber and non-fiber starches in the formula. Together, these findings position *Parabacteroides* as a primary degrader of SCF, although mechanistic insights remain limited.

These findings are consistent with the ISAPP definition of a prebiotic: “a substrate that is selectively utilized by host microorganisms, conferring a health benefit” ^105^. SCF induced small shift in the GI microbiota, selectively increasing *Parabacteroides*, and provided a cognitive benefit in healthy older adults. *Parabacteroides* responded to SCF in 90% of the healthy cohort, consistent with the ‘Health-Associated Core Keystone’ (HACK) index which identified *P. distasonis, P. merdae*, and *F. saccharivorans* as core and stability-associated taxa ^106^. Prior work links *P. distasonis* with anti-inflammatory effects, gut barrier protection ^107–109^, and bioactive metabolite production (e.g., GABA)^110^, relevant to gut-brain communication and health outcomes ^102,111,112^. Although *Parabacteroides* did not directly mediate cognitive benefits in the SCOPE study, its consistent response to SCF suggests its potential role as a keystone taxon in healthy older adults.

In this study, fecal SCFA concentration differences did not reach statistical significance; however, their directional changes align with prior dietary fiber interventions and remain plausible candidates for gut-brain signaling, given their ability to cross and modulate the blood-brain barrier’s structural integrity ^34,35,113–116^. Notably, fecal SCFA concentrations in the present study were lower than those reported in a younger, male-only SCF trial using a higher dose (21g/d) ^19^, which may reflect differences in participant characteristics such transit time, absorption, and/or dilution with greater stool mass. Prior human studies show that colonic SCFA delivery influences anticipatory brain reward responses and attenuates cortisol reactivity to psychosocial stress ^35^. Despite biological plausibility, in the present study, mediation and moderation models did not support fecal SCFAs as either mediators or moderators of the SCF effect.

Moderation analyses suggested that the SCF effect on RT varied with the magnitude of change in *Parabacteroides*. RT improvements were greatest at smaller *Parabacteroides* changes (−1SD), whereas larger *Parabacteroides* changes were not associated with faster RTs. Furthermore, responders consistently performed quicker at baseline and endpoints, while non-responders exhibited larger improvements from SCF for RT speeds. These exploratory findings suggest that *Parabacteroides* may exert a context-dependent influence, but its enrichment may not be linearly beneficial for cognitive performance.

Secondary taxa provided additional exploratory context for the SCF-RT association. Although mediation models did not support Dialister or *Fusicatenibacter* as intermediaries of the SCF effect on RT, their descriptive patterns were informative. Bacillota members such as *Fusicatenibacter* are capable of cross-feeding on substrates released by members of the Bacteroidota phylum ^117^ (e.g., *Parabacteroides*), producing beneficial metabolites including acetate, lactate, formate, and succinate ^118^. Potentially, *Fusicatenibacter* may have had the role of the main secondary consumer working with *Parabacteroides* in response to SCF. Responders had higher baseline levels of *F. saccharivorans* which showed negligible enrichment with SCF, whereas non-responders exhibited a relatively larger enrichment in *F. saccharivorans*. In contrast,

*D. invisus* and *D. hominis* were enriched only in non-responders and remained below detectable levels in responders. A prior study linked *Fusicatenibacter saccharivorans* with executive and visuospatial functions with community-dwelling older adults ^119^, whereas *Dialister invisus* was reported to be more abundant in older adults with mild cognitive impairment ^119^. Together, these patterns suggest that baseline community composition and differential responsiveness may contribute to cognitive variability, though mechanistic inferences remain preliminary.

The SCOPE study has several limitations. First, the cohort was predominantly white, female, and highly educated, which may restrict generalizability. Mediation and moderation analysis were exploratory and did not fully account for the crossover design in its statistical framework and utilized change and baseline adjustments. Shotgun metagenomic analyses were limited to a small subset of responders and non-responders, reducing power to detect subtle effects. The absence of blood samples prevented assessment of circulating SCFAs or other metabolites, while the targeted fecal metabolomics panel may have overlooked additional relevant compounds (e.g., tryptophan catabolites). Furthermore, fecal SCFAs can be influenced by colonic transit time and may only reflect a fraction *in vivo* production. Systemic inflammatory markers and neuroactive metabolites were not assessed and should be incorporated in future mechanistic trials. Although participants agreed to maintain their habitual diets, inter-individual dietary variation may have introduced unmeasured noise into microbiome and metabolite outcomes. Despite these limitations, the SCOPE study provides novel evidence linking SCF to selective cognitive benefits, supported by exploratory insights into microbial and SCFA response.

In conclusion, daily consumption of SCF for four weeks improved RT in middle aged and older adults and induced a microbiota shift dominated by *Parabacteroides*. Exploratory moderation suggested a threshold effect, with RT benefits most evident at smaller changes in *Parabacteroides*. Notably, cognitive improvements were also observed in non-responders with negligible *Parabacteroides*, indicating that cognitive benefits are unlikely to hinge on a single taxon but may instead depend on the ecological context and balance between health associated keystone microbes (e.g., *Parabacteroides* and *Fusicatenibacter*) for contributing complementary routes to induce GI-brain interactions ^120^. Host factors may also contribute to interindividual variability in cognitive response to SCF. These findings position SCF as a dietary fiber with potential cognitive relevance, while underscoring the need for multi-omics, untargeted metabolomics, dosing trials, and systemic biomarker assessments to refine mechanistic models and advance precision nutrition strategies for cognitive health.

## Data Availability

All data produced in the present study are available upon reasonable request to the authors

## Acknowledgments

We thank Dr. Michael J. Miller for his expertise, equipment, and facility to conduct the *in vitro* culturing experiment. We also thank Dr. Pratik Banerjee and his graduate students (Papri Suraya Rahman and Edwin Valenzuela De Leon) for providing the *P. distasonis* strain and assisting with the equipment set-up. We are also grateful to the HDH graduate students: Alexis Baldeon, Marahi Perez-Tamayo, Nadine Veasly, and Maria Sanabria-Veaz; HDH staff: Eva Cornman, Myra Esmail, Karen Gutierrez, and Tehila Abdiel; and NAK graduate students Shelby Keyes and John Kim, as well as the undergraduate research assistants from HDH and NAK laboratories, for their assistance during the appointments, sample processing, and data management.

## Author Contributions

HDH, NAK, DAA and TAH: designed the research; MDB and TM: recruited and enrolled participants and coordinated study procedures; DAA: conducted the *in vitro* experiment; DAA, TAH, SM, TM and RS: conducted the experiments and testing appointments; MA, DR and DAA: managed biological samples collection and storages; DAA and TAH: managed study data; DAA and NS: prepared data for bioinformatic pipelines; DAA and TAH: analyzed the data: DAA, TAH, NAK and HDH: wrote the manuscript; NAK and HDH: had primary responsibility for final content. All authors read and approved the final manuscript.

## Author Disclosures

The authors report no conflict of interest.

## Funding

This work was supported by Tate & Lyle Ingredients Americas LLC (to NAK) and the USDA NIFA, Hatch Project (1026591, to HDH).

## Data Availability

Data described in the manuscript, code book, and analytic code will be made available upon request.

## Abbreviations

(CANTAB): *Parabacteroides distasonis* (*P. distasonis*), Cambridge Neuropsychological Test Automated Battery
(CAZyme): carbohydrate-active enzymes
(ERP): Event-related potential
(False Discovery Rate): FDR
(CON, control): Maltodextrin
(PSS): Perceived Stress Score
(PANAS),: Positive and Negative Affect Schedule
(POMS): Profile of Moods Survey
(RT): Reaction Time
SEM: SE (Standard Error)
(SCFA): (Standard Error of the Mean), Short-chain fatty acids
(BCFA): Branched-chain fatty acids
(SCF): Soluble corn fiber/ PROMITOR® Soluble Fibre™
(ACC): Accuracy

## Notes

### Competing Interest Statement

The authors have declared no competing interest.

### Clinical Trial

NCT05066425

### Funding Statement

This work was supported by Tate & Lyle Ingredients Americas LLC and the USDA NIFA, Hatch Project (1026591, to HDH).

### Author Declarations

The University of Illinois at Urbana-Champaign Institutional Review Board (IRB) has reviewed and approved the research study as described. Soluble Corn Fiber for Promoting Executive Function (SCOPE) Study, IRB Number: 21839.

### Summary of Updates

Corrected model summaries that were reporting the same thing but with different references (SCF or CON); they are now consistent.

## References

1. Morris MC, Wang Y, Barnes LL, Bennett DA, Dawson-Hughes B, Booth SL. Nutrients and bioactives in green leafy vegetables and cognitive decline: Prospective study. Neurology. 2018;90(3):E214–E222. doi:10.1212/WNL.0000000000004815

2. Perim Baldo M, Aboussaleh Y, Turrini A, et al. Adherence to a Mediterranean-Style Diet and effects on Cognition in Adults: A Qualitative evaluation and Systematic Review of Longitudinal and Prospective Trials. 2016;3:1. doi:10.3389/fnut.2016.00022

3. Khan NA, Raine LB, Donovan SM, Hillman CH. IV. The cognitive implications of obesity and nutrition in childhood. Monogr Soc Res Child Dev. 2014;79(4):51–71. doi:10.1111/mono.12130

4. Cho SS, Qi L, Fahey GC, Klurfeld DM. Consumption of cereal fiber, mixtures of whole grains and bran, and whole grains and risk reduction in type 2 diabetes, obesity, and cardiovascular disease. American Journal of Clinical Nutrition. 2013;98(2):594–619. doi:10.3945/ajcn.113.067629

5. Trock B, Lanza E, Greenwald P, Lanza E, Greenwald P. REVIEW Dietary Fiber, Vegetables, and Colon Cancer: Critical Review and Meta-Analyses of the Epidemiologic Evidence. https://academic.oup.com/jnci/article/82/8/650/882222

6. Gov D. Dietary Guidelines for Americans Make Every Bite Count With the Dietary Guidelines. https://www.

7. Marchesi JR, Ravel J. The vocabulary of microbiome research: a proposal. Microbiome. 2015;3(1). doi:10.1186/s40168-015-0094-5

8. Fleming SA, Monaikul S, Patsavas AJ, Waworuntu R V., Berg BM, Dilger RN. Dietary polydextrose and galactooligosaccharide increase exploratory behavior, improve recognition memory, and alter neurochemistry in the young pig. Nutr Neurosci. 2019;22(7):499–512. doi:10.1080/1028415X.2017.1415280

9. Gronier B, Savignac HM, Miceli M Di, et al. Increased cortical neuronal responses to NMDA and improved attentional set-shifting performance in rats following prebiotic (B-GOS s ) ingestion. doi:10.1016/j.euroneuro.2017.11.001

10. Khan NA, Raine LB, Drollette ES, Scudder MR, Kramer AF, Hillman CH. Dietary Fiber Is Positively Associated with Cognitive Control among Prepubertal Children. J Nutr. 2015;145(1):143–149. doi:10.3945/JN.114.198457

11. Hassevoort KM, Lin AS, Khan NA, Hillman CH, Cohen NJ. Added sugar and dietary fiber consumption are associated with creativity in preadolescent children. Nutr Neurosci. 2020;23(10):791–802. doi:10.1080/1028415X.2018.1558003

12. Naveed S, Venäläinen T, Eloranta AM, et al. Associations of dietary carbohydrate and fatty acid intakes with cognition among children. Public Health Nutr. 2020;23(9):1657–1663. doi:10.1017/S1368980019003860

13. Cryan JF, O KJ, M Cowan CS, et al. The Microbiota-Gut-Brain Axis. Physiol Rev. 2019;99:1877–2013. doi:10.1152/physrev.00018.2018.-The

14. Stewart ML, Nikhanj SD, Timm DA, Thomas W, Slavin JL. Evaluation of the effect of four fibers on laxation, gastrointestinal tolerance and serum markers in healthy humans. Ann Nutr Metab. 2010;56(2):91–98. doi:10.1159/000275962

15. Shuan W, Tan K, Fen P, et al. The Role of Soluble Corn Fiber on Glycemic and Insulin Response. doi:10.3390/nu12040961

16. Jakeman SA, Henry CN, Martin BR, et al. Soluble corn fiber increases bone calcium retention in postmenopausal women in a dose-dependent manner: A randomized crossover trial. American Journal of Clinical Nutrition. 2016;104(3):837–843. doi:10.3945/ajcn.116.132761

17. Klosterbuer AS, Hullar MAJ, Li F, et al. Gastrointestinal effects of resistant starch, soluble maize fibre and pullulan in healthy adults. doi:10.1017/S0007114513000019

18. LinAmy AHM, Nichols BL, Quezada-Calvillo R, et al. Unexpected high digestion rate of cooked starch by the Ct-maltase-glucoamylase small intestine mucosal α-glucosidase subunit. PLoS One. 2012;7(5). doi:10.1371/journal.pone.0035473

19. Vester Boler BM, Rossoni Serao MC, Bauer LL, et al. Digestive physiological outcomes related to polydextrose and soluble maize fibre consumption by healthy adult men. British Journal of Nutrition. 2011;106(12):1864–1871. doi:10.1017/S0007114511002388

20. Maathuis A, Venema K, Hoffman A, Evans A, Sanders L. The effect of the undigested fraction of maize products on the activity and composition of the microbiota determined in a dynamic in vitro model of the human proximal large intestine. J Am Coll Nutr. 2009;28(6):657–666. doi:10.1080/07315724.2009.10719798

21. Whisner CM, Martin BR, Nakatsu CH, et al. Soluble corn fiber increases calcium absorption associated with shifts in the gut microbiome: A randomized dose-response trial in free-living pubertal females. Journal of Nutrition. 2016;146(7):1298–1306. doi:10.3945/jn.115.227256

22. Arroyo MC, Laurie I, Rotsaert C, Marzorati M, Risso D, Karnik K. Age-Dependent Prebiotic Effects of Soluble Corn Fiber in M-SHIME® Gut Microbial Ecosystems. Plant Foods for Human Nutrition. 2023;78(1):213–220. doi:10.1007/s11130-023-01043-z

23. Holscher HD, Gregory Caporaso J, Hooda S, Brulc JM, Fahey GC, Swanson KS. Fiber supplementation influences phylogenetic structure and functional capacity of the human intestinal microbiome: Follow-up of a randomized controlled trial. American Journal of Clinical Nutrition. 2015;101(1):55–64. doi:10.3945/ajcn.114.092064

24. Brown AJ, Goldsworthy SM, Barnes AA, et al. The orphan G protein-coupled receptors GPR41 and GPR43 are activated by propionate and other short chain carboxylic acids. Journal of Biological Chemistry. 2003;278(13):11312–11319. doi:10.1074/jbc.M211609200

25. Cani PD, Lecourt E, Dewulf EM, et al. Gut microbiota fermentation of prebiotics increases satietogenic and incretin gut peptide production with consequences for appetite sensation and glucose response after a meal. American Journal of Clinical Nutrition. 2009;90(5):1236–1243. doi:10.3945/ajcn.2009.28095

26. Corrêa-Oliveira R, Fachi JL, Vieira A, Sato FT, Vinolo MAR. Regulation of immune cell function by short-chain fatty acids. Clin Transl Immunology.John Wiley and Sons Inc. 2016;5(4). doi:10.1038/cti.2016.17

27. Dalile B, Van Oudenhove L, Vervliet B, Verbeke K. The role of short-chain fatty acids in microbiota–gut–brain communication. Nat Rev Gastroenterol Hepatol.Nature Publishing Group. 2019;16(8):461–478. doi:10.1038/s41575-019-0157-3

28. Zhou SY, Guo ZN, Yang Y, Qu Y, Jin H. Gut-brain axis: Mechanisms and potential therapeutic strategies for ischemic stroke through immune functions. Front Neurosci.Frontiers Media S.A. 2023;17. doi:10.3389/fnins.2023.1081347

29. Silva YP, Bernardi A, Frozza RL. The Role of Short-Chain Fatty Acids From Gut Microbiota in Gut-Brain Communication. Front Endocrinol (Lausanne).Frontiers Media S.A. 2020;11. doi:10.3389/fendo.2020.00025

30. Mansuy-Aubert V, Ravussin Y. Short chain fatty acids: the messengers from down below. Front Neurosci. 2023;17. doi:10.3389/fnins.2023.1197759

31. Diamond A. Executive functions. Annu Rev Psychol.Annual Reviews Inc. 2013;64:135–168. doi:10.1146/annurev-psych-113011-143750

32. Duncan-Johnson CC. P300 latency: A new metric of information processing. Psychophysiology. 1981;18(3):207–215. doi:10.1111/j.1469-8986.1981.tb03020.x

33. Polich J. Updating P300: An integrative theory of P3a and P3b. Clinical Neurophysiology. 2007;118(10):2128–2148. doi:10.1016/j.clinph.2007.04.019

34. La Torre D, Verbeke K, Dalile B. Dietary fibre and the gut–brain axis: microbiota-dependent and independent mechanisms of action. Gut Microbiome. 2021;2. doi:10.1017/gmb.2021.3

35. Dalile B, Fuchs A, La Torre D, Vervliet B, Van Oudenhove L, Verbeke K. Colonic butyrate administration modulates fear memory but not the acute stress response in men: A randomized, triple-blind, placebo-controlled trial. Prog Neuropsychopharmacol Biol Psychiatry. 2024;131. doi:10.1016/j.pnpbp.2024.110939

36. Cheng J, Hu H, Ju Y, et al. Gut microbiota-derived short-chain fatty acids and depression: deep insight into biological mechanisms and potential applications. Gen Psychiatr.BMJ Publishing Group. 2024;37(1). doi:10.1136/gpsych-2023-101374

37. Luqman A, He M, Hassan A, et al. Mood and microbes: a comprehensive review of intestinal microbiota’s impact on depression. Front Psychiatry.Frontiers Media SA. 2024;15. doi:10.3389/fpsyt.2024.1295766

38. Haskell CF, Scholey AB, Jackson PA, et al. Cognitive and mood effects in healthy children during 12 weeks’ supplementation with multi-vitamin/minerals. British Journal of Nutrition. 2008;100(5):1086–1096. doi:10.1017/S0007114508959213

39. Leyrolle Q, Cserjesi R, D.G.H. Mulders M, et al. Prebiotic effect on mood in obese patients is determined by the initial gut microbiota composition: A randomized, controlled trial. Brain Behav Immun. 2021;94:289–298. doi:10.1016/j.bbi.2021.01.014

40. Godin-Leisure-Time-Exercise-Questionnaire_070815.

41. Edwards CG, Walk AM, Cannavale CN, et al. Dietary choline is related to neural efficiency during a selective attention task among middle-aged adults with overweight and obesity. 10.1080/1028415X20191623456. 2019;24(4):269–278. doi:10.1080/1028415X.2019.1623456

42. Chatrian GE, Lettich E, Nelson PL. American Journal of EEG Technology Ten Percent Electrode System for Topographic Studies of Spontaneous and Evoked EEG Activities Ten Percent Electrode System for Topographic Studies of Spontaneous and Evoked EEG Activities. Published online 2015. doi:10.1080/00029238.1985.11080163

43. Delorme A, Makeig S. EEGLAB: An open source toolbox for analysis of single-trial EEG dynamics including independent component analysis. J Neurosci Methods. 2004;134(1):9–21. doi:10.1016/J.JNEUMETH.2003.10.009

44. Lopez-Calderon J, Luck SJ. ERPLAB: An open-source toolbox for the analysis of event-related potentials. Front Hum Neurosci. 2014;8(1 APR):213. doi:10.3389/FNHUM.2014.00213/BIBTEX

45. Kamijo K, Pontifex MB, Khan NA, et al. The association of childhood obesity to neuroelectric indices of inhibition. Psychophysiology. 2012;49(10):1361–1371. doi:10.1111/J.1469-8986.2012.01459.X

46. Walk AM, Edwards CG, Baumgartner NW, et al. The role of retinal carotenoids and age on neuroelectric indices of attentional control among early to middle-aged adults. Front Aging Neurosci. 2017;9(JUN):183. doi:10.3389/FNAGI.2017.00183/BIBTEX

47. Luck SJ, Gaspelin N. How to get statistically significant effects in any ERP experiment (and why you shouldn’t). Psychophysiology. 2017;54(1):146–157. doi:10.1111/PSYP.12639

48. Pontifex MB. Stylized Topographic Map Plugin for EEGLAB/ERPLAB.

49. Horecka KM, Dulas MR, Schwarb H, Lucas HD, Duff M, Cohen NJ. Reconstructing relational information. Hippocampus. 2018;28(2):164–177. doi:10.1002/hipo.22819

50. Langley C, Sahakian B, Robbins T. Cambridge neuropsychological test automated battery (cantab). SAGE Publications Ltd. 2023;0:435–468.

51. Wild K V., Musser ED. The cambridge neuropsychological test automated battery in the assessment of executive functioning. In: Handbook of Executive Functioning. Springer New York; 2014:171–190. doi:10.1007/978-1-4614-8106-5_11

52. Stephen J. Gibson. The Measurement of Mood States in Older Adults. The Journals of Gerontology: Series B. 1997;52B(4):167–174.

53. Boyle GJ, Helmes E, Matthews G, Izard CE. Measures of Affect Dimensions. In: Measures of Personality and Social Psychological Constructs. Elsevier Inc.; 2015:190–224. doi:10.1016/B978-0-12-386915-9.00008-5

54. Humboldt S von, Monteiro A, Leal I. Validation of the PANAS: A Measure of Positive and Negative Affect for Use with Cross-National Older Adults. Rev Eur Stud. 2017;9(2):10. doi:10.5539/res.v9n2p10

55. Lee EH. Review of the psychometric evidence of the perceived stress scale. Asian Nurs Res (Korean Soc Nurs Sci). 2012;6(4):121–127. doi:10.1016/j.anr.2012.08.004

56. Walters W, Hyde ER, Berg-Lyons D, et al. Improved Bacterial 16S rRNA Gene (V4 and V4-5) and Fungal Internal Transcribed Spacer Marker Gene Primers for Microbial Community Surveys. mSystems. 2016;1(1). doi:10.1128/msystems.00009-15

57. Callahan BJ, McMurdie PJ, Rosen MJ, Han AW, Johnson AJA, Holmes SP. DADA2: High-resolution sample inference from Illumina amplicon data. Nat Methods. 2016;13(7):581–583. doi:10.1038/nmeth.3869

58. Bolyen E, Rideout JR, Dillon MR, et al. Reproducible, interactive, scalable and extensible microbiome data science using QIIME 2. Nat Biotechnol.Nature Publishing Group. 2019;37(8):852–857. doi:10.1038/s41587-019-0209-9

59. Yilmaz P, Parfrey LW, Yarza P, et al. The SILVA and “all-species Living Tree Project (LTP)” taxonomic frameworks. Nucleic Acids Res. 2014;42(D1). doi:10.1093/nar/gkt1209

60. Morton JT, Marotz C, Washburne A, et al. Establishing microbial composition measurement standards with reference frames. Nat Commun. 2019;10(1). doi:10.1038/s41467-019-10656-5

61. McMurdie PJ, Holmes S. Phyloseq: An R Package for Reproducible Interactive Analysis and Graphics of Microbiome Census Data. PLoS One. 2013;8(4). doi:10.1371/journal.pone.0061217

62. Kenneth Helrich. Official Methods for Analysis of the Association of Official Analytical Chemists.; 1990.

63. Chaney AL, MEP. Modified Reagents for Determination of Urea and Ammonia. Clin Chem. 1962;8(2):130–132.

64. Holscher HD, Doligale JL, Bauer LL, et al. Gastrointestinal tolerance and utilization of agave inulin by healthy adults. Food Funct. 2014;5(6):1142–1149. doi:10.1039/c3fo60666j

65. Flickinger EA, Schreijen EMWC, Patil AR, et al. Nutrient digestibilities, microbial populations, and protein catabolites as affected by fructan supplementation of dog diets Get access Arrow. J Anim Sci. 2003;81(8):2008–2018.

66. Holscher HD, Chumpitazi BP, Dahl WJ, et al. Perspective: Assessing Tolerance to Nondigestible Carbohydrate Consumption. Advances in Nutrition. 2022;13(6):2084–2097. doi:10.1093/advances/nmac091

67. Maki K, Rains T, Kelley K, Cook C, Schild A, Gietl E. Fibermalt is well tolerated in healthy men and women at intakes up to 60 g/d: a randomized, double-blind, crossover trial. Int J Food Sci Nutr. 2013;63(3):274–281.

68. Tingley D, Yamamoto T, Hirose K, Keele L, Imai K. Journal of Statistical Software Mediation: R Package for Causal Mediation Analysis.; 2014. http://www.jstatsoft.org/

69. Chi WE, Huang S, Jeon M, Park ES, Melguizo T, Kezar A. A Practical Guide to Causal Mediation Analysis: Illustration With a Comprehensive College Transition Program and Nonprogram Peer and Faculty Interactions. Front Educ (Lausanne). 2022;7. doi:10.3389/feduc.2022.886722

70. Sohn MB, Li H. Compositional mediation analysis for microbiome studies. Annals of Applied Statistics. 2019;13(1):661–681. doi:10.1214/18-AOAS1210

71. Aiken LS, West SG. Interaction Effects. In: Encyclopedia of Statistics in Behavioral Science. Wiley; 2005. doi:10.1002/0470013192.bsa306

72. Preacher KJ, Curran PJ, Bauer DJ. Computational Tools for Probing Interactions in Multiple Linear Regression, Multilevel Modeling, and Latent Curve Analysis. Vol 31.; 2006. http://www.quantpsy.org/.437

73. García-Bayona L, Comstock LE. Streamlined genetic manipulation of diverse bacteroides and parabacteroides isolates from the human gut microbiota. mBio. 2019;10(4). doi:10.1128/mBio.01762-19

74. Douglas GM, Maffei VJ, Zaneveld JR, et al. PICRUSt2 for prediction of metagenome functions. Nat Biotechnol.Nature Research. 2020;38(6):685–688. doi:10.1038/s41587-020-0548-6

75. Langille MGI, Zaneveld J, Caporaso JG, et al. Predictive functional profiling of microbial communities using 16S rRNA marker gene sequences. Nat Biotechnol. 2013;31(9):814–821. doi:10.1038/nbt.2676

76. Beghini F, McIver LJ, Blanco-Míguez A, et al. Integrating taxonomic, functional, and strain-level profiling of diverse microbial communities with biobakery 3. Elife. 2021;10. doi:10.7554/eLife.65088

77. Blanco-Míguez A, Beghini F, Cumbo F, et al. Extending and improving metagenomic taxonomic profiling with uncharacterized species using MetaPhlAn 4. Nat Biotechnol. 2023;41(11):1633–1644. doi:10.1038/s41587-023-01688-w

78. Segata N, Izard J, Waldron L, et al. Metagenomic biomarker discovery and explanation. Genome Biol. 2011;12(6). doi:10.1186/gb-2011-12-6-r60

79. Drula E, Garron ML, Dogan S, Lombard V, Henrissat B, Terrapon N. The carbohydrate-active enzyme database: Functions and literature. Nucleic Acids Res. 2022;50(D1):D571–D577. doi:10.1093/nar/gkab1045

80. Cantarel BI, Coutinho PM, Rancurel C, Bernard T, Lombard V, Henrissat B. The Carbohydrate-Active EnZymes database (CAZy): An expert resource for glycogenomics. Nucleic Acids Res. 2009;37(SUPPL. 1). doi:10.1093/nar/gkn663

81. Zheng J, Ge Q, Yan Y, Zhang X, Huang L, Yin Y. DbCAN3: Automated carbohydrate-Active enzyme and substrate annotation. Nucleic Acids Res. 2023;51(W1):W115–W121. doi:10.1093/nar/gkad328

82. Erdfelder E, FAul F, Buchner A, Lang AG. Statistical power analyses using G*Power 3.1: Tests for correlation and regression analyses. Behav Res Methods. 2009;41(4):1149–1160. doi:10.3758/BRM.41.4.1149

83. R Core Team. R: A language and environment for statistical computing. *Foundation for Statistical Computing*. Preprint posted online 2013.

84. H. Wickham. ggplot2: Elegant Graphics for Data Analysis. *Springer-VerlagNew York*. Published online 2016.

85. Kwak SK, Kim JH. Statistical data preparation: Management of missing values and outliers. Korean J Anesthesiol.Korean Society of Anesthesiologists. 2017;70(4):407–411. doi:10.4097/kjae.2017.70.4.407

86. Kafadar K. John Tukey and Robustness. Statistical Science. 2003;3(18(3)):319–331. doi:1076102419

87. Zhang X, Mallick H, Tang Z, et al. Negative binomial mixed models for analyzing microbiome count data. BMC Bioinformatics. 2017;18(1). doi:10.1186/s12859-016-1441-7

88. Sweeny AR, Lemon H, Ibrahim A, et al. A mixed-model approach for estimating drivers of microbiota community composition and differential taxonomic abundance. mSystems. Published online July 25, 2023. doi:10.1128/msystems.00040-23

89. Jamieson PE, Smart EB, Bouranis JA, et al. Gut enterotype-dependent modulation of gut microbiota and their metabolism in response to xanthohumol supplementation in healthy adults. Gut Microbes. 2024;16(1). doi:10.1080/19490976.2024.2315633

90. Zwietering MH, Jongenburger I, Rombouts FM, Van ’ K, Riet T. Modeling of the Bacterial Growth Curve PUm.; 1990. https://journals.asm.org/journal/aem

91. Mallick H, Rahnavard A, McIver LJ, et al. Multivariable association discovery in population-scale meta-omics studies. PLoS Comput Biol. 2021;17(11). doi:10.1371/journal.pcbi.1009442

92. Yang C, Mai J, Cao X, Burberry A, Cominelli F, Zhang L. ggpicrust2: an R package for PICRUSt2 pre-dicted functional profile analysis and visualiza-tion. *Bioinformatics*, YYYY.:0-0. doi:10.1093/bioinformatics/xxxxx

93. Edwards CG, Walk AM, Thompson S V., et al. Effects of 12-week avocado consumption on cognitive function among adults with overweight and obesity. International Journal of Psychophysiology. 2020;148:13–24. doi:10.1016/j.ijpsycho.2019.12.006

94. Ford NA, Spagnuolo P, Kraft J, Bauer E. Nutritional Composition of Hass Avocado Pulp. Foods.Multidisciplinary Digital Publishing Institute (MDPI). 2023;12(13). doi:10.3390/foods12132516

95. Thompson S V., Bailey MA, Taylor AM, et al. Avocado Consumption Alters Gastrointestinal Bacteria Abundance and Microbial Metabolite Concentrations among Adults with Overweight or Obesity: A Randomized Controlled Trial. Journal of Nutrition. 2021;151(4):753–762. doi:10.1093/jn/nxaa219

96. Fda. Review of the Scientific Evidence on the Physiological Effects of Certain Non-Digestible Carbohydrates.; 2018.

97. Mysonhimer AR, Holscher HD. Gastrointestinal Effects and Tolerance of Nondigestible Carbohydrate Consumption. Advances in Nutrition. 2022;13(6):2237–2276. doi:10.1093/advances/nmac094

98. Mehren A, Luque CD, Brandes M, et al. Intensity-dependent effects of acute exercise on executive function. Neural Plast. 2019;2019. doi:10.1155/2019/8608317

99. Timm DA, Thomas W, Boileau TW, Williamson-Hughes PS, Slavin JL. Polydextrose and soluble corn fiber increase five-day fecal wet weight in healthy men and women. Journal of Nutrition. 2013;143(4):473–478. doi:10.3945/jn.112.170118

100. Hooda S, Vester Boler BM, Rossoni Serao MC, et al. 454 pyrosequencing reveals a shift in fecal microbiota of healthy adult men consuming polydextrose or soluble corn fiber. Journal of Nutrition. 2012;142(7):1259–1265. doi:10.3945/jn.112.158766

101. Cantu-Jungles TM, Hamaker BR. Tuning Expectations to Reality: Don’t Expect Increased Gut Microbiota Diversity with Dietary Fiber. Journal of Nutrition. 2023;153(11):3156–3163. doi:10.1016/j.tjnut.2023.09.001

102. Shimizu H, Nakajima M, Miyanaga A, et al. Characterization and Structural Analysis of a Novel exo-Type Enzyme Acting on β-1,2-Glucooligosaccharides from Parabacteroides distasonis. Biochemistry. 2018;57(26):3849–3860. doi:10.1021/acs.biochem.8b00385

103. Ducarmon QR, Karcher N, Tytgat HLP, et al. Large-scale computational analyses of gut microbial CAZyme repertoires enabled by Cayman. Preprint posted online January 8, 2024. doi:10.1101/2024.01.08.574624

104. Lapébie P, Lombard V, Drula E, Terrapon N, Henrissat B. Bacteroidetes use thousands of enzyme combinations to break down glycans. Nat Commun. 2019;10(1). doi:10.1038/s41467-019-10068-5

105. Gibson GR, Hutkins R, Sanders ME, et al. Expert consensus document: The International Scientific Association for Probiotics and Prebiotics (ISAPP) consensus statement on the definition and scope of prebiotics. Nat Rev Gastroenterol Hepatol.Nature Publishing Group. 2017;14(8):491–502. doi:10.1038/nrgastro.2017.75

106. Goel A, Shete O, Goswami S, et al. Toward a health-associated core keystone index for the human gut microbiome. Cell Rep. 2025;44(3):115378. doi:10.1016/j.celrep.2025.115378

107. Cui Y, Zhang L, Wang X, et al. Roles of intestinal Parabacteroides in human health and diseases. FEMS Microbiol Lett.Oxford University Press. 2022;369(1). doi:10.1093/femsle/fnac072

108. Cuffaro B, Boutillier D, Desramaut J, et al. Characterization of Two Parabacteroides distasonis Candidate Strains as New Live Biotherapeutics against Obesity. Cells. 2023;12(9). doi:10.3390/cells12091260

109. Ezeji JC, Sarikonda DK, Hopperton A, et al. Parabacteroides distasonis: intriguing aerotolerant gut anaerobe with emerging antimicrobial resistance and pathogenic and probiotic roles in human health. *Gut Microbes*.*Bellwether Publishing*, Ltd. 2021;13(1). doi:10.1080/19490976.2021.1922241

110. Liang Z, Di N, Li L, Yang D. Gut microbiota alterations reveal potential gut–brain axis changes in polycystic ovary syndrome. J Endocrinol Invest. 2021;44(8):1727–1737. doi:10.1007/s40618-020-01481-5

111. Wei W, Wong CC, Jia Z, et al. Parabacteroides distasonis uses dietary inulin to suppress NASH via its metabolite pentadecanoic acid. Nat Microbiol. 2023;8(8):1534–1548. doi:10.1038/s41564-023-01418-7

112. Liu D, Zhang S, Li S, et al. Indoleacrylic acid produced by Parabacteroides distasonis alleviates type 2 diabetes via activation of AhR to repair intestinal barrier. BMC Biol. 2023;21(1). doi:10.1186/s12915-023-01578-2

113. Keaney J, Campbell M. The dynamic blood-brain barrier. FEBS Journal. 2015;282(21):4067–4079. doi:10.1111/febs.13412

114. Van Dokkum W, Wezendonk B, Srikumar TS, Van Den Heuvel EGHM. Effect of nondigestible oligosaccharides on large-bowel functions, blood lipid concentrations and glucose absorption in young healthy male subjects. Eur J Clin Nutr. 1999;53(1):1–7. doi:10.1038/sj.ejcn.1600668

115. Pedersen A, Sandström B, Van Amelsvoort JMM. The effect of ingestion of inulin on blood lipids and gastrointestinal symptoms in healthy females. British Journal of Nutrition. 1997;78(2):215–222. doi:10.1079/bjn19970141

116. Dalile B, La Torre D, Kalc P, et al. Extruded Wheat Bran Consumption Increases Serum Short-Chain Fatty Acids but Does Not Modulate Psychobiological Functions in Healthy Men: A Randomized, Placebo-Controlled Trial. Front Nutr. 2022;9. doi:10.3389/fnut.2022.896154

117. Wang S, Mu L, Yu C, et al. Microbial collaborations and conflicts: unraveling interactions in the gut ecosystem. Gut Microbes.Taylor and Francis Ltd. 2024;16(1). doi:10.1080/19490976.2023.2296603

118. Takada T, Kurakawa T, Tsuji H, Nomoto K. Fusicatenibacter saccharivorans gen. nov., sp. nov., isolated from human faeces. Int J Syst Evol Microbiol. 2013;63(PART10):3691–3696. doi:10.1099/ijs.0.045823-0

119. Li J, Zhu S, Wang Y, et al. Metagenomic association analysis of cognitive impairment in community-dwelling older adults. Neurobiol Dis. 2023;180. doi:10.1016/j.nbd.2023.106081

120. Loh JS, Mak WQ, Tan LKS, et al. Microbiota–gut–brain axis and its therapeutic applications in neurodegenerative diseases. Signal Transduct Target Ther.Springer Nature. 2024;9(1). doi:10.1038/s41392-024-01743-1

