## Supplementary Material for "Effects of Soluble Corn Fiber Consumption on Executive Functions and Gut Microbiota in Middle to Older Age Adults: A Randomized Controlled Crossover Trial"

*SUPPLEMENTAL TABLES & FIGURES*

**Supplemental Table 1.** Secondary outcomes for mood, stress, memory, affect, and physical activity across treatments

| | Maltodextrin | | Soluble Corn Fiber | | Treatment Effect (ΔSCF - ΔCON) | | | |
|---|---|---|---|---|---|---|---|---|
| | **PRE** | **POST** | **PRE** | **POST** | | | | |
| **Secondary Measures** | Mean ± SEM *(n)* | Mean ± SEM *(n)* | Mean ± SEM *(n)* | Mean ± SEM *(n)* | β | SE | 95% CI | *p* |
| **POMS** | | | | | | | | |
| Tension | 6.37 ± 0.61 *(38)* | 6.82 ± 0.70 *(38)* | 5.39 ± 0.62 *(38)* | 5.78 ± 0.54 *(37)* | 0.02 | 0.17 | [-0.32, 0.36] | 0.92 |
| Confusion | 5.26 ± 0.49 *(38)* | 5.58 ± 0.48 *(38)* | 4.92 ± 0.51 *(38)* | 5.57 ± 0.61 (37) | 0.01 | 0.14 | [-0.26, 0.29] | 0.92 |
| Depression | 4.05 ± 0.83 *(38)* | 5.03 ± 0.88 *(38)* | 3.74 ± 1.01 *(38)* | 5.14 ± 1.13 *(37)* | 0.03 | 0.28 | [-0.52, 0.59] | 0.91 |
| Anger | 4.11 ± 0.84 *(38)* | 4.05 ± 0.86 *(38)* | 2.84 ± 0.63 *(38)* | 4.16 ± 1.03 *(37)* | 0.14 | 0.27 | [-0.39, 0.67] | 0.61 |
| Fatigue | 4.74 ± 0.63 *(38)* | 4.82 ± 0.58 *(38)* | 4.50 ± 0.54 *(38)* | 4.54 ± 0.65 *(37)* | -0.11 | 0.24 | [-0.58, 0.35] | 0.63 |
| Vigour | 14.00 ± 0.88 *(38)* | 14.37 ± 0.94 *(38)* | 14.71 ± 1.11 *(38)* | 14.57 ± 1.07 *(37)* | -0.06 | 0.17 | [-0.39, 0.27] | 0.74 |
| TMD | 10.53 ± 2.89 *(38)* | 11.92 ± 3.33 *(38)* | 6.68 ± 3.29 *(38)* | 10.62 ± 3.79 *(37)* | 0.38 | 0.60 | [-0.78, 1.53] | 0.53 |
| **iPosition** | | | | | | | | |
| Accurate Single Item Placement | 2.47 ± 0.13 *(36)* | 2.48 ± 0.13 *(37)* | 2.75 ± 0.11 *(37)* | 2.74 ± 0.11 *(37)* | -0.03 | 0.12 | [-0.25, 0.20] | 0.82 |
| Original Misplacement | 245.9 ± 11.62 *(36)* | 237.3 ± 12.68 *(37)* | 213.6 ± 10.52 *(37)* | 210.1 ± 10.10 *(37)* | 0.25 | 0.32 | [-0.39, 0.89] | 0.45 |
| **PANAS** | | | | | | | | |
| Positive | 33.00 ± 1.01 *(37)* | 33.18 ± 1.11 *(39)* | 33.01 ± 1.26 *(40)* | 33.41 ± 1.25 *(39)* | -0.33 | 1.30 | [-2.85, 2.20] | -0.80 |
| Negative | 15.41 ± 0.87 *(37)* | 16.28 ± 0.91 *(39)* | 14.40 ± 0.93 *(40)* | 14.61 ± 0.95 *(38)* | -0.09 | 0.12 | [-0.32, 0.13] | 0.42 |
| **PSS-10** | 19.95 ± 0.53 *(39)* | 19.68 ± 0.44 *(38)* | 18.45 ± 0.67 *(40)* | 18.4 ± 0.67 *(39)* | 0.26 | 0.67 | [-1.04, 1.56] | 0.70 |
| **Godin** | 36.46 ± 3.50 *(39)* | 36.69 ± 4.63 *(39)* | 33.83 ± 3.10 *(40)* | 37.20 ± 4.31 *(40)* | 0.13 | 0.32 | [-0.50, 0.76] | 0.70 |

Values are descriptive means ± standard error of the mean (SEM) and sample size (*n*) across each timepoint. Total SCFAs and BCFAs are pooled measures, within respective category. Linear mixed-effect models included participant ID as a random effect, and the fixed effects of treatment, time, and their interaction. Reported are the treatment-by-time interaction estimates (β, Standard Error (SE), 95% confidence interval (CI), *p*-values). Positive and Negative Affect Scores (PANAS; likert scale), perceived stress scores (PSS-10; [0-40] total scores), profile of mood surveys (POMS), relational memory (iPosition; accurate single item place [0-6] and original misplacement (pixel units)).

*SUPPLEMENTAL TABLES & FIGURES*

**Supplemental Table 2.** Rapid Visual Information Processing (RVP) task performance across treatments

| | Maltodextrin | | Soluble Corn Fiber | | |
|---|---|---|---|---|---|
| | PRE | POST | PRE | POST | |
| **Rapid Visual Information Processing Task** | Median [IQR] | Median [IQR] | Median [IQR] | Median [IQR] | *p* |
| RVPA | 1.0 [0.1] | 1.0 [0.0] | 1.0 [0.0] | 1.0 [0.0] | 0.24 |
| RVPLSD | 122.6 [83.7] | 85.0 [55.9] | 104.9 [100.1] | 99.0 [81.7] | 0.24 |
| RVPMDL | 435.5 [53.9] | 430.0 [63.0] | 423.7 [55.4] | 439.3 [63.9] | 0.71 |
| RVPML | 467.3 [72.0] | 458.6 [83.5] | 458.3 [51.7] | 475.2 [72.7] | 0.76 |
| RVPPFA | 0.0 [0.0] | 0.0 [0.0] | 0.0 [0.0] | 0.0 [0.0] | 0.30 |
| RVPPH | 0.9 [0.2] | 0.9 [0.2] | 0.9 [0.2] | 0.9 [0.1] | 0.21 |
| RVPTFA | 1.0 [2.3] | 1.0 [1.3] | 1.0 [2.0] | 1.0 [2.0] | 0.28 |
| RVPTH | 47.0 [12.8] | 47.0 [8.5] | 49.0 [9.3] | 48.0 [7.0] | 0.21 |
| RVPTM | 7.0 [12.8] | 7.0 [8.5] | 5.0 [9.3] | 6.0 [7.0] | 0.21 |

Values are descriptive median ± interquartile range (IQR) and sample size ($n$ = 35) across each timepoint. Reported values include $p$-values from the paired Wilcoxon Signed-Rank tests.

**<u>Supplemental Table 3.</u>** Stop Signal Task (SST) Scores performance across treatments

| | **Maltodextrin** | | **Soluble Corn Fiber** | | |
|---|---|---|---|---|---|
| | **PRE** | **POST** | **PRE** | **POST** | |
| **Stop Signal Task** | Median [IQR] | Median [IQR] | Median [IQR] | Median [IQR] | *p* |
| SSTDEG | 1.0 [2.0] | 1.0 [2.3] | 1.0 [2.0] | 0.5 [3.3] | 0.69 |
| SSTDES | 41.0 [5.0] | 40.5 [5.0] | 40.5 [3.3] | 41.0 [4.0] | 0.67 |
| SSTMFRTI | 1.0 [0.0] | 1.0 [0.0] | 1.0 [0.0] | 1.0 [0.0] | 0.82 |
| SSTFSG | 1.0 [0.0] | 1.0 [0.0] | 1.0 [0.0] | 1.0 [0.0] | NaN |
| SSTMRTG | 498.5 [72.8] | 517.8 [96.5] | 506.8 [77.8] | 500.8 [98.4] | 0.62 |
| SSTMRTGF | 526.8 [83.5] | 528.0 [79.3] | 525.8 [105.5] | 519.3 [64.5] | 0.79 |
| SSTMRTGG | 499.0 [67.3] | 505.0 [102.5] | 495.5 [68.5] | 490.3 [96.5] | 0.73 |
| SSTMRTGS | 513.0 [87.3] | 512.8 [74.8] | 500.3 [84.5] | 506.0 [98.8] | 0.70 |
| SSTMT | 1.0 [2.0] | 1.0 [2.0] | 0.0 [1.0] | 0.5 [2.3] | 0.76 |
| SSTSSRT | 226.9 [27.8] | 223.5 [38.9] | 223.3 [41.2] | 227.4 [38.5] | 0.49 |

Values are descriptive median ± interquartile range (IQR) and sample size ($n$ = 35) across each timepoint. Reported values include $p$-values from the paired Wilcoxon Signed-Rank tests.

**Supplemental Table 4.** Spatial Working Memory (SWM) Task performance across treatments

| | **Maltodextrin** | | **Soluble Corn Fiber** | | |
|---|---|---|---|---|---|
| | **PRE** | **POST** | **PRE** | **POST** | |
| **Spatial Working Memory Task** | Median [IQR] | Median [IQR] | Median [IQR] | Median [IQR] | *p* |
| SWMBE | 6.5 [12.5] | 6.5 [11.3] | 3.5 [12.0] | 2.5 [11.0] | 0.29 |
| SWMBE4 | 0.0 [0.0] | 0.0 [0.0] | 0.0 [0.0] | 0.0 [0.0] | 0.84 |
| SWMBE6 | 0.0 [3.3] | 0.0 [2.3] | 0.0 [2.0] | 0.0 [1.3] | 0.88 |
| SWMBE8 | 3.5 [9.3] | 6.0 [10.3] | 2.5 [10.3] | 1.5 [10.3] | 0.20 |
| SWMDE | 0.0 [0.0] | 0.0 [0.0] | 0.0 [0.0] | 0.0 [0.0] | 0.78 |
| SWMDE6 | 0.0 [0.0] | 0.0 [0.0] | 0.0 [0.0] | 0.0 [0.0] | 0.82 |
| SWMDE8 | 0.0 [0.0] | 0.0 [0.0] | 0.0 [0.0] | 0.0 [0.0] | 0.52 |
| SWMS | 7.5 [5.0] | 7.0 [3.0] | 6.5 [6.0] | 6.5 [5.3] | 0.73 |
| SWMS6 | 3.0 [2.0] | 3.0 [2.0] | 3.0 [3.0] | 2.5 [2.0] | 0.85 |
| SWMTE | 6.5 [13.3] | 7.5 [12.0] | 3.5 [12.3] | 3.0 [11.0] | 0.25 |
| SWMTE4 | 0.0 [0.0] | 0.0 [0.0] | 0.0 [0.0] | 0.0 [0.0] | 0.84 |
| SWMTE6 | 0.0 [3.3] | 0.0 [3.0] | 0.0 [2.0] | 0.0 [1.3] | 0.87 |
| SWMTE8 | 3.5 [9.3] | 6.0 [9.5] | 2.5 [11.0] | 2.5 [10.3] | 0.14 |
| SWMWE | 0.0 [0.0] | 0.0 [1.0] | 0.0 [0.3] | 0.0 [0.0] | 0.34 |
| SWMWE6 | 0.0 [0.0] | 0.0 [0.0] | 0.0 [0.0] | 0.0 [0.0] | 0.71 |
| SWMWE8 | 0.0 [0.0] | 0.0 [0.0] | 0.0 [0.0] | 0.0 [0.0] | 0.24 |

Values are descriptive median ± interquartile range (IQR) and sample size ($n$ = 35) across each timepoint. Reported values include $p$-values from the paired Wilcoxon Signed-Rank tests. Significance threshold $p$: '*' <0.05, '†' <0.1.

**Supplemental Table 5.** Intra-Extra Dimensional (IED) Set Shift Task performance across treatments

| | Maltodextrin | | Soluble Corn Fiber | | |
|---|---|---|---|---|---|
| | **PRE** | **POST** | **PRE** | **POST** | |
| **Intra-Extra Dimensional Set Shift Task** | Median [IQR] | Median [IQR] | Median [IQR] | Median [IQR] | *p* |
| IEDECS | 9.5 [8.0] | 9.0 [3.3] | 9.0 [4.0] | 9.0 [6.3] | 0.52 |
| IEDEEDS | 3.0 [15.0] | 3.0 [5.0] | 3.0 [4.5] | 2.0 [5.5] | 0.21 |
| IEDERPRE | 6.0 [4.0] | 6.0 [3.0] | 5.0 [2.0] | 5.0 [3.0] | 0.49 |
| IEDES1 | 1.0 [1.0] | 0.0 [1.0] | 0.0 [1.0] | 0.0 [1.0] | 0.62 |
| IEDES2 | 1.0 [0.0] | 1.0 [0.0] | 1.0 [0.0] | 1.0 [0.0] | 0.67 |
| IEDES3 | 0.0 [1.0] | 0.0 [1.0] | 0.0 [1.0] | 0.0 [1.0] | 0.28 |
| IEDES4 | 0.0 [0.0] | 0.0 [0.0] | 0.0 [0.0] | 0.0 [0.0] | 0.14 |
| IEDES5 | 1.0 [0.0] | 1.0 [0.5] | 1.0 [0.0] | 1.0 [0.0] | 0.80 |
| IEDES6 | 1.0 [1.0] | 0.0 [1.0] | 0.0 [1.0] | 0.5 [1.0] | 0.37 |
| IEDES7 | 1.0 [0.0] | 1.0 [0.0] | 1.0 [0.0] | 1.0 [0.0] | 0.63 |
| IEDES8 | 3.0 [15.0] | 3.0 [5.0] | 3.0 [4.5] | 2.0 [5.5] | 0.21 |
| IEDES9 | 1.0 [0.8] | 1.0 [1.0] | 1.0 [1.0] | 2.0 [2.0] | 0.72 |
| IEDTL | 90,459.0 [48,505.5] | 80,500.0 [27.661.3] | 79,616.0 [36,407.0] | 73,600.0 [25,161.0] | 0.83 |
| IEDTLS1 | 4,939.0 [2,913.3] | 4,528.5 [2520.8] | 4,136.5 [1,751.8] | 3,712.5 [1,103.8] | 0.66 |
| IEDTLS2 | 6,185.0 [2,402.8] | 6,240.5 [1,550.0] | 5,645.5 [1,888.3] | 6,085.0 [1,504.8] | 0.91 |
| IEDTLS3 | 9,310.5 [4,928.8] | 9,702.5 [4,267.5] | 7,673.5 [3,524.5] | 7,715 [3,753.0] | 0.41 |
| IEDTLS4 | 6,615.0 [2,792.8] | 6,792.0 [2,271.0] | 6,699.0 [2,261.5] | 6,626.5 [2,037.5] | 0.94 |
| IEDTLS5 | 7,803.0 [4,3260.5] | 7,840.0 [2,799.5] | 8,185.5 [3,183.3] | 7,870.5 [2,926.5] | 0.93 |
| IEDTLS6 | 9,434.0 [4,261.5] | 9,308.0 [3,781.0] | 8,558.0 [3,820.3] | 9,006.0 [3,918.0] | 0.52 |
| IEDTLS7 | 7,728.0 [2,699.5] | 7,059.0 [1,702.0] | 7,481.5 [3,373.0] | 7,098.0 [2,151.3] | 0.94 |
| IEDTLS8 | 18,522.0 [25,190.5] | 15,853.0 [11,204.0] | 15,770.5 [10,405.5] | 13,415.0 [14,949.5] | 0.40 |
| IEDTLS9 | 7,781.5 [4,697.0] | 7,988.5 [3,579.3] | 8,187.0 [5,429.0] | 9,255.0 [5,799.5] | 0.53 |
| IEDTT | 72.0 [23.3] | 68.5 [9.3] | 67.0 [12.0] | 69.0 [11.0] | 0.55 |
| IEDTTA | 76.0 [48.5] | 70.0 [23.3] | 68.0 [16.0] | 71.0 [22.5] | 0.24 |
| IEDTTS1 | 7.0 [2.0] | 6.0 [1.0] | 6.0 [1.0] | 6.0 [1.0] | 0.61 |
| IEDTTS2 | 7.0 [0.0] | 7.0 [0.0] | 7.0 [0.0] | 7.0 [0.0] | 0.89 |
| IEDTTS3 | 6.0 [2.0] | 6.0 [2.0] | 6.0 [1.3] | 6.0 [2.3] | 0.45 |
| IEDTTS4 | 6.0 [0.0] | 6.0 [0.0] | 6.0 [0.0] | 6.0 [0.0] | 0.07 |
| IEDTTS5 | 7.0 [0.0] | 7.0 [0.5] | 7.0 [0.0] | 7.0 [0.0] | 0.98 |
| IEDTTS6 | 7.0 [1.0] | 6.0 [1.0] | 6.0 [1.0] | 6.5 [1.0] | 0.31 |
| IEDTTS7 | 7.0 [0.0] | 7.0 [0.0] | 7.0 [0.0] | 7.0 [0.0] | 0.68 |
| IEDTTS8 | 11.0 [30.5] | 11.0 [7.5] | 11.0 [9.8] | 10.5 [9.8] | 0.27 |
| IEDTTS9 | 7.0 [1.8] | 7.0 [2.0] | 7.0 [3.0] | 8.0 [2.5] | 0.87 |
| IEDYCOST | 9.0 [0.3] | 9.0 [0.0] | 9.0 [0.0] | 9.0 [0.0] | 0.57 |
| IEDYERT | 10.5 [12.8] | 9.5 [4.5] | 9.0 [4.0] | 9.0 [6.5] | 0.35 |
| IEDYERTA | 14.0 [29.8] | 10.0 [13.3] | 9.5 [7.5] | 11.0 [10.5] | 0.16 |
| IEDYSTCO | 66.0 [14.5] | 66.0 [9.0] | 67.0 [12.3] | 67.5 [12.0] | 0.78 |

Values are descriptive median ± interquartile range (IQR) and sample size ($n$ = 35) across each timepoint. Reported values include $p$-values from the paired Wilcoxon Signed-Rank tests.

**Supplemental Table 6**. Differential abundance of 16S phylum across treatment and time

| | **Maltodextrin** | | **Soluble Corn Fiber** | | **Treatment Effect (ΔSCF - ΔCON)** | | |
|---|---|---|---|---|---|---|---|
| | **PRE** | **POST** | **PRE** | **POST** | | | |
| **Phylum (% of sequence)** | Mean ± SEM *(n)* | Mean ± SEM *(n)* | Mean ± SEM *(n)* | Mean ± SEM *(n)* | β | 95% CI | FDR *p* |
| Bacillota | 54.20 ± 1.40 *(38)* | 56.83 ± 1.77 *(38)* | 50.56 ± 1.65 *(37)* | 52.69 ± 1.78 *(37)* | 0.00 | [-0.10, 0.10] | 0.10† |
| Bacteroidota | 33.64 ± 1.41 *(38)* | 31.20 ± 1.81 *(38)* | 37.19 ± 1.91 *(37)* | 37.81 ± 1.86 *(37)* | 0.11 | [-0.05, 0.28] | 0.09† |
| Actinobacteriota | 4.66 ± 0.59 *(38)* | 4.72 ± 0.63 *(38)* | 4.11 ± 0.60 *(37)* | 3.46 ± 0.50 *(37)* | -0.17 | [-0.49, 0.14] | 0.29 |
| Pseudomonadota | 3.78 ± 0.73 *(38)* | 3.82 ± 0.87 *(38)* | 4.08 ± 0.92 *(37)* | 3.45 ± 0.69 *(37)* | -0.14 | [-0.58, 0.31] | 0.91 |
| Verrucomicrobiota | 2.13 ± 0.57 *(38)* | 2.09 ± 0.70 *(38)* | 1.88 ± 0.55 *(37)* | 1.01 ± 0.23 *(37)* | 0.08 | [-0.79, 0.96] | 0.91 |

Values are means ± standard error of the mean (SEM) and sample size (n) across each timepoint. For differential abundance of genera, a generalized linear mixed-effect negative binomial model included participant ID as a random effect, and the fixed effects of treatment, time, and their interaction. Reported are the treatment-by-time interaction log-scaled estimates (β, Standard Error (SE), 95% confidence interval (CI), FDR-adjusted *p*-values). Significance threshold *p*: '***' <0.001, '**' <0.01, '*' <0.05, '†' <0.1.

*SUPPLEMENTAL TABLES & FIGURES*

**Supplemental Table 7.** Differential abundance of 16S genera classified as *Incertae_Sedis* or small-abundance across treatment and time

| | **Maltodextrin** | | **Soluble Corn Fiber** | | **Treatment Effect (ΔSCF - ΔCON)** | | |
|---|---|---|---|---|---|---|---|
| | **PRE** | **POST** | **PRE** | **POST** | | | |
| **Genus (% of Sequence)** | Mean ± SEM *(n)* | Mean ± SEM *(n)* | Mean ± SEM *(n)* | Mean ± SEM *(n)* | β | 95% CI | FDR *p* |
| Incertae_Sedis | 1.21 ± 0.43 *(38)* | 1.23 ± 0.34 *(38)* | 0.71 ± 0.13 *(37)* | 5.18 ± 0.92 *(37)* | 1.93 | [1.39, 2.46] | < 0.001*** |
| *.Ruminococcus._torques_group* | 0.96 ± 0.23 *(38)* | 0.95 ± 0.26 *(38)* | 0.89 ± 0.19 *(37)* | 0.41 ± 0.09 *(37)* | -0.77 | [-1.25, -0.28] | < 0.001*** |
| *Holdemanella* | 0.32 ± 0.19 *(38)* | 0.33 ± 0.22 *(38)* | 0.21 ± 0.13 *(37)* | 0.40 ± 0.31 *(37)* | -0.69 | [-1.09, -0.30] | 0.002** |
| *Adlercreutzia* | 0.59 ± 0.13 *(38)* | 0.66 ± 0.15 *(38)* | 0.41 ± 0.06 *(37)* | 0.32 ± 0.07 *(37)* | -0.61 | [-1.05, -0.17] | 0.016* |
| *Faecalitalea* | 0.21 ± 0.13 *(38)* | 0.19 ± 0.10 *(38)* | 0.15 ± 0.08 *(37)* | 0.15 ± 0.09 *(37)* | -1.27 | [-2.13, -0.42] | 0.018* |
| *.Eubacterium._ruminantium_group* | 0.67 ± 0.25 *(38)* | 0.52 ± 0.18 *(38)* | 0.73 ± 0.25 *(37)* | 0.07 ± 0.04 *(37)* | -1.66 | [-2.62, -0.69] | 0.023* |
| DTU*089* | 0.04 ± 0.01 *(38)* | 0.03 ± 0.01 *(38)* | 0.03 ± 0.00 *(37)* | 0.11 ± 0.03 *(37)* | 1.10 | [0.41, 1.80] | < 0.001*** |
| *Negativibacillus* | 0.23 ± 0.10 *(38)* | 0.23 ± 0.09 *(38)* | 0.19 ± 0.07 *(37)* | 0.08 ± 0.03 *(37)* | -0.90 | [-1.53, -0.27] | 0.001** |
| *Lachnospiraceae_ND3007_group* | 0.11 ± 0.02 *(38)* | 0.12 ± 0.02 *(38)* | 0.11 ± 0.03 *(37)* | 0.06 ± 0.01 *(37)* | -0.75 | [-1.37, -0.14] | 0.02* |
| *Candidatus_Soleaferrea* | 0.02 ± 0.00 *(38)* | 0.02 ± 0.00 *(38)* | 0.02 ± 0.00 *(37)* | 0.01 ± 0.00 *(37)* | -0.61 | [-1.10, -0.12] | 0.001** |
| *Dielma* | 0.02 ± 0.01 *(38)* | 0.01 ± 0.01 *(38)* | 0.01 ± 0.00 *(37)* | 0.01 ± 0.00 *(37)* | -0.67 | [-1.73, 0.39] | 0.02* |

Values are means ± standard error of the mean (SEM) and sample size (n) across each timepoint. For differential abundance of genera, a generalized linear mixed-effect negative binomial model included participant ID as a random effect, and the fixed effects of treatment, time, and their interaction. Reported are the treatment-by-time interaction log-scaled estimates (β, Standard Error (SE), 95% confidence interval (CI), FDR-adjusted *p*-values). Significance threshold *p*: '***' <0.001, '**' <0.01, '*' <0.05, '†' <0.1.

**Supplemental Table 8.** Fecal fermentation end-products across treatment and time

| | **Maltodextrin** | | **Soluble Corn Fiber** | | | | | |
|---|---|---|---|---|---|---|---|---|
| | **PRE** | **POST** | **PRE** | **POST** | **Treatment Effect (ΔSCF - ΔCON)** | | | |
| **Fermentation end-products** | Mean ± SEM *(n)* | Mean ± SEM *(n)* | Mean ± SEM *(n)* | Mean ± SEM *(n)* | β | SE | 95% CI | *p* |
| Indole | 1.07 ± 0.09 *(32)* | 1.08 ± 0.09 *(31)* | 1.08 ± 0.10 *(28)* | 1.02 ± 0.11 *(21)* | -0.16 | 0.17 | [-0.49, 0.16] | 0.32 |
| Total Indole | 1.52 ± 0.17 *(32)* | 1.33 ± 0.11 *(32)* | 1.34 ± 0.10 *(30)* | 1.13 ± 0.11 *(26)* | -0.13 | 0.23 | [-0.59. 0.34] | 0.59 |
| 4-Methylphenol | 2.79 ± 0.28 *(35)* | 2.42 ± 0.20 *(35)* | 2.79 ± 0.23 *(35)* | 2.15 ± 0.16 *(31)* | -0.29 | 0.35 | [-0.97, 0.40] | 0.42 |
| Total Phenol | 2.72 ± 0.30 *(37)* | 2.48 ± 0.21 *(36)* | 2.79 ± 0.23 *(35)* | 2.21 ± 0.16 *(32)* | -0.34 | 0.38 | [-1.08, 0.41] | 0.38 |
| Ammonia | 125.6 ± 12.57 *(38)* | 108.8 ± 8.37 *(37)* | 112.2 ± 10.3 *(37)* | 99.85 ± 8.40 *(37)* | 0.82 | 0.29 | [-23.3, 29.5] | 0.82 |

Values are descriptive means ± standard error of the mean (SEM) and sample size (n) across each timepoint. Total indole includes pooled values from indole, 2-methylindole, 3-methylindole, and 7-methylindole. Total phenol includes pooled values from 4-methylphenol, 4-ethylphenol, and phenol. Linear mixed-effect models included participant ID as a random effect, and the fixed effects of treatment, time, and their interaction. Reported are the treatment-by-time interaction estimates (β, Standard Error (SE), 95% confidence interval (CI), *p*-values). Significance threshold *p*: '***' <0.001, '**' <0.01, '*' <0.05, '†' <0.1.

**Supplemental Table 9.** Stool Characteristics across treatment and time

| | **Maltodextrin** | | **Soluble Corn Fiber** | | | | | |
|---|---|---|---|---|---|---|---|---|
| | **PRE** | **POST** | **PRE** | **POST** | **Treatment Effect (ΔSCF - ΔCON)** | | | |
| **Stool Characteristics** | Mean ± SEM *(n)* | Mean ± SEM *(n)* | Mean ± SEM *(n)* | Mean ± SEM *(n)* | β | SE | 95% CI | *p* |
| Consistency | 3.31 ± 0.17 *(38)* | 3.79 ± 0.15 *(38)* | 3.63 ± 0.17 *(36)* | 3.83 ± 0.14 *(36)* | -0.28 | 0.25 | [-0.77, 0.21] | 0.27 |
| Ease of Passage | 2.15 ± 0.12 *(38)* | 1.91 ± 0.11 *(38)* | 2.24 ± 0.15 *(36)* | 1.86 ± 0.11 *(36)* | -0.16 | 0.19 | [-0.52, 0.21] | 0.41 |
| pH | 6.95 ± 0.07 *(38)* | 6.92 ± 0.09 *(37)* | 7.02 ± 0.09 *(37)* | 6.82 ± 0.08 *(37)* | -0.16 | 0.13 | [-0.40, 0.09] | 0.22 |
| Bowel Movements | 0.87 ± 0.03 *(38)* | 0.88 ± 0.03 *(38)* | 0.85 ± 0.03 *(36)* | 0.86 ± 0.03 *(36)* | -0.01 | 0.04 | [-0.08, 0.07] | 0.9 |

Values are descriptive means ± standard error of the mean (SEM) and sample size (*n*) across each timepoint. Linear mixed-effect models included participant ID as a random effect, and the fixed effects of treatment, time, and their interaction. Reported are the treatment-by-time interaction estimates (β, Standard Error (SE), 95% confidence interval (CI), *p*-values).

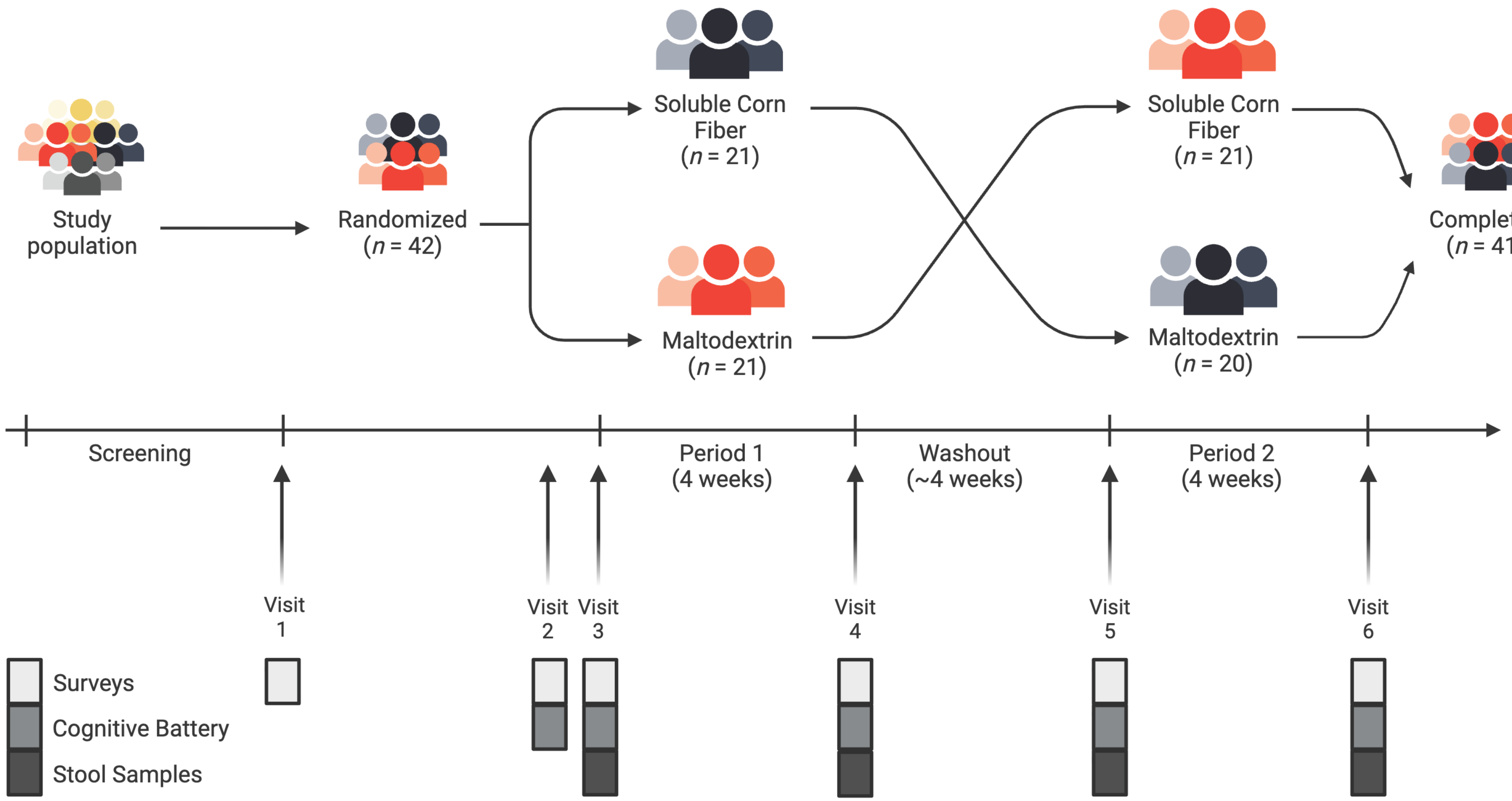


**Supplemental Figure 1. Study design timeline and sample collection procedures.** Participants completed six laboratory visits across two 4-week intervention periods separate by a washout. Visit 1: Eligibility (screening, consent/surveys); if eligible, they were randomized and scheduled (~2 weeks) for their Visit 2 with stool collection and diet record deployed for Visit 3 first intervention baseline. Visit 2: Practice/familiarization sessions with EEG cap placement, scheduled ~1 week before to testing visit. Visits 3 and 5: Pre-intervention testing days with the full EEG-based cognitive battery. Visits 4 and 6: Post-intervention follow-ups with the same testing battery. Stools samples were collected over the final 5-days of each intervention period and returned at the subsequent testing visit. (*Figure created using BioRender)*

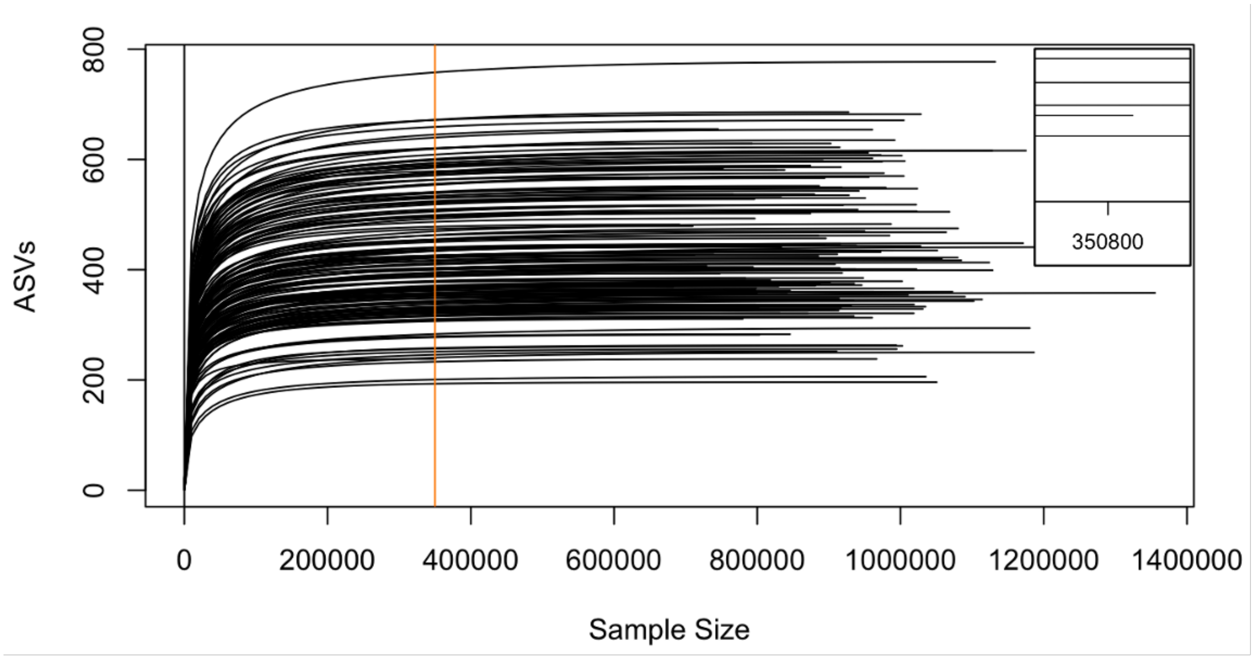


**Supplemental Figure 2. Rarefaction curve showing amplicon sequence variants across sampling depths**. The rarefaction curve illustrates the number of amplicon sequence variants (ASVs) observed as a function of sequencing depth across subject samples. The orange line represents the rarefaction sampling depth used (350,000 reads) to standardize sequencing depth across all samples. Rarefaction sampling depth selected based on the min depth shown in the upper right corner with the smallest sequencing depth value at 350,840.

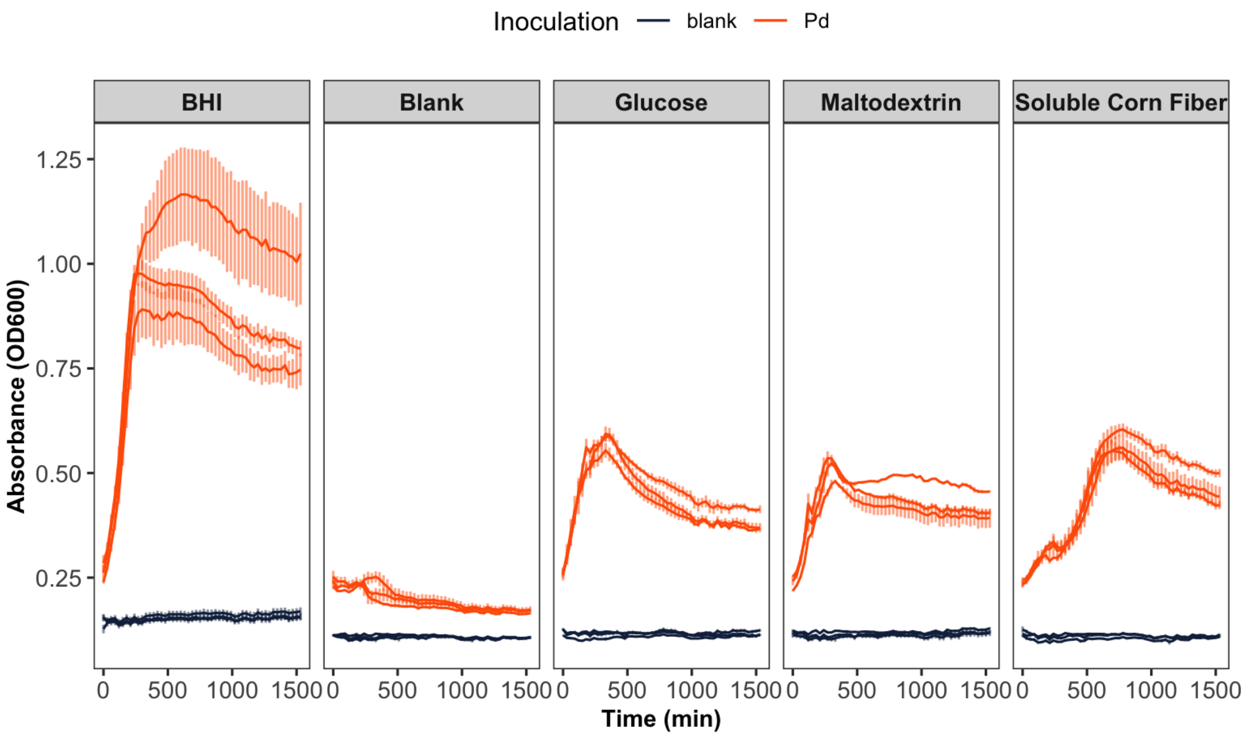


**Supplemental Figure 3. Parabacteroides distasonis growth curves of CHO-supplemented M9 of each biological replicate.** Growth curves of Parabacteroides distasonis (Pd) in M9 minimal media with BHI, blank, glucose, maltodextrin, or soluble corn fiber over 25.5 hrs (displayed in minutes on plot). Blue indicates media with Pd and red indicates media without Pd. Each line represents a biological replicate mean from 3 technical replicates.

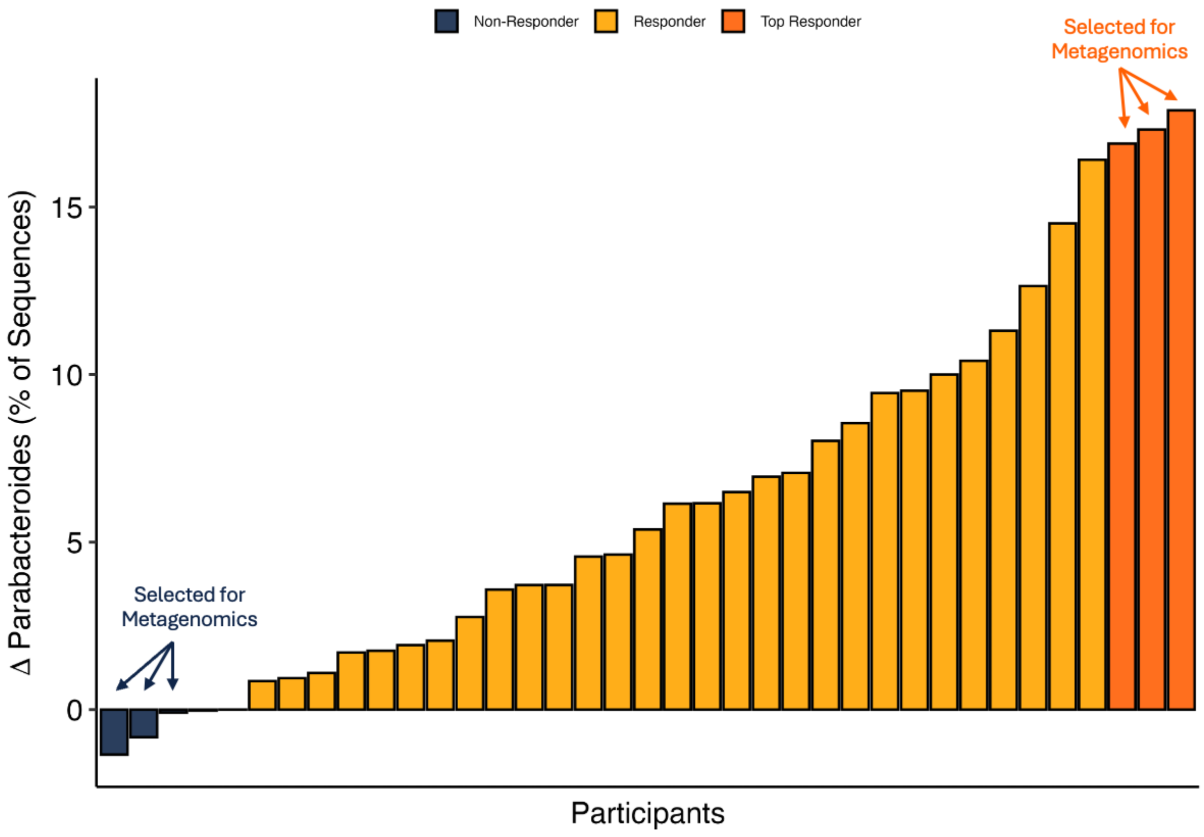


**Supplemental Figure 4. Participant-level changes in 16S rRNA–derived Parabacteroides relative abundance from PRE to POST.** Bars show each individual's change (POST-PRE) in *Parabacteroides* abundance (%) within the SCF arm; orange bars denote responders (increased abundance), navy bars denote non-responders (no change or decrease). The top three responders with the largest positive change are highlighted in dark orange and selected for shotgun metagenomic sequencing.

*SUPPLEMENTAL TABLES & FIGURES*

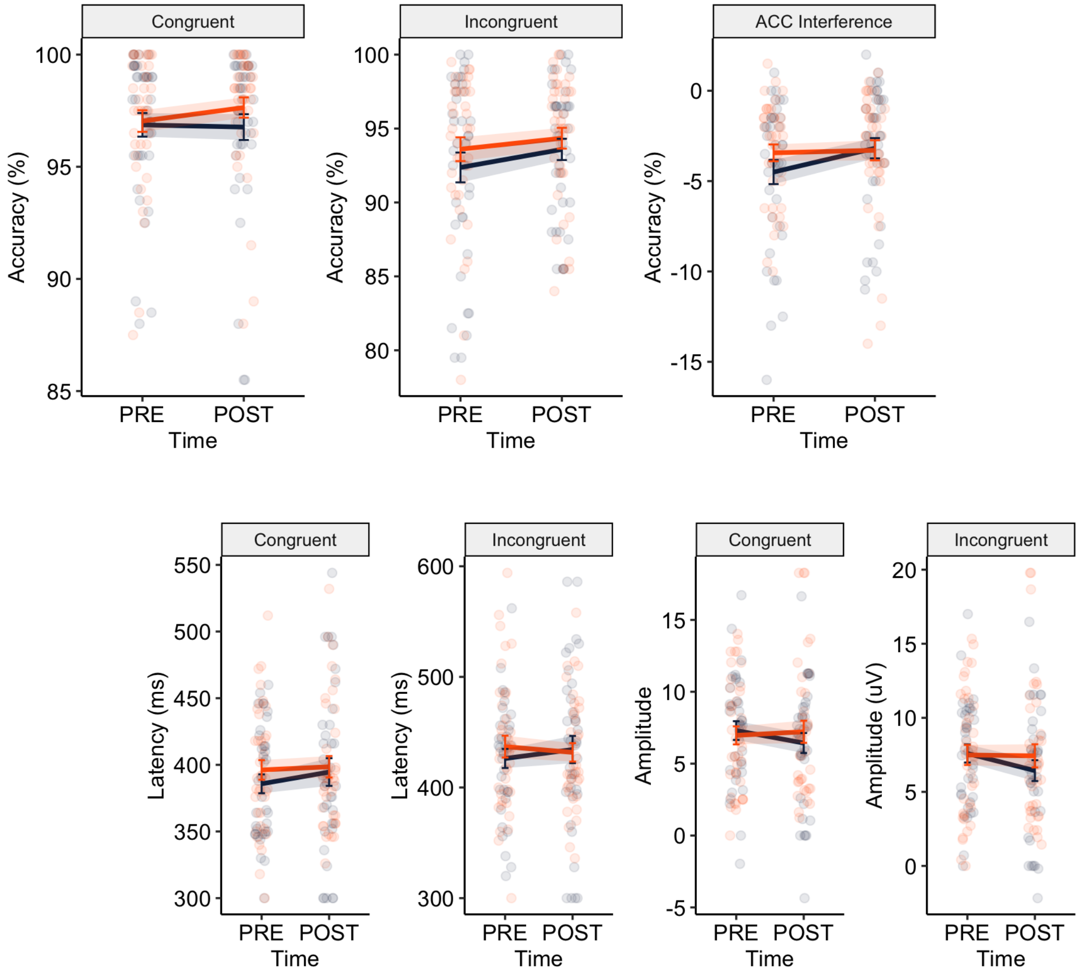


**Supplemental Figure 5. Panels demonstrating the effects of SCF consumption compared to CON on Behavioral and ERP Flanker Task measures**. Values represent the raw unadjusted values (top row) accuracy (%), and the P3 components (bottom row) for Amplitude (µV; left), and the latency (ms; right).

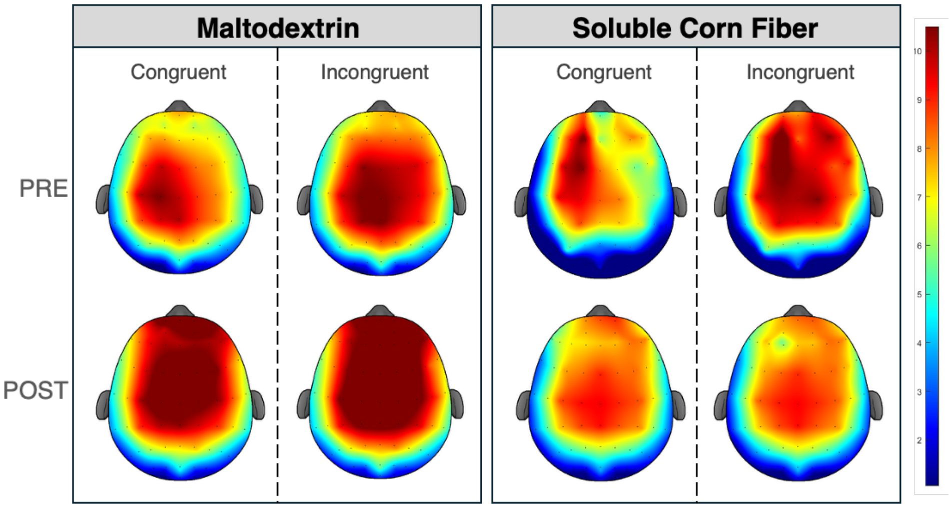


**Supplemental Figure 6. Topographic plot of the P3 300-600 ms.** Window representing the spatial P3 amplitude distributions during the ERP flanker task for each participant at PRE and POST treatment.

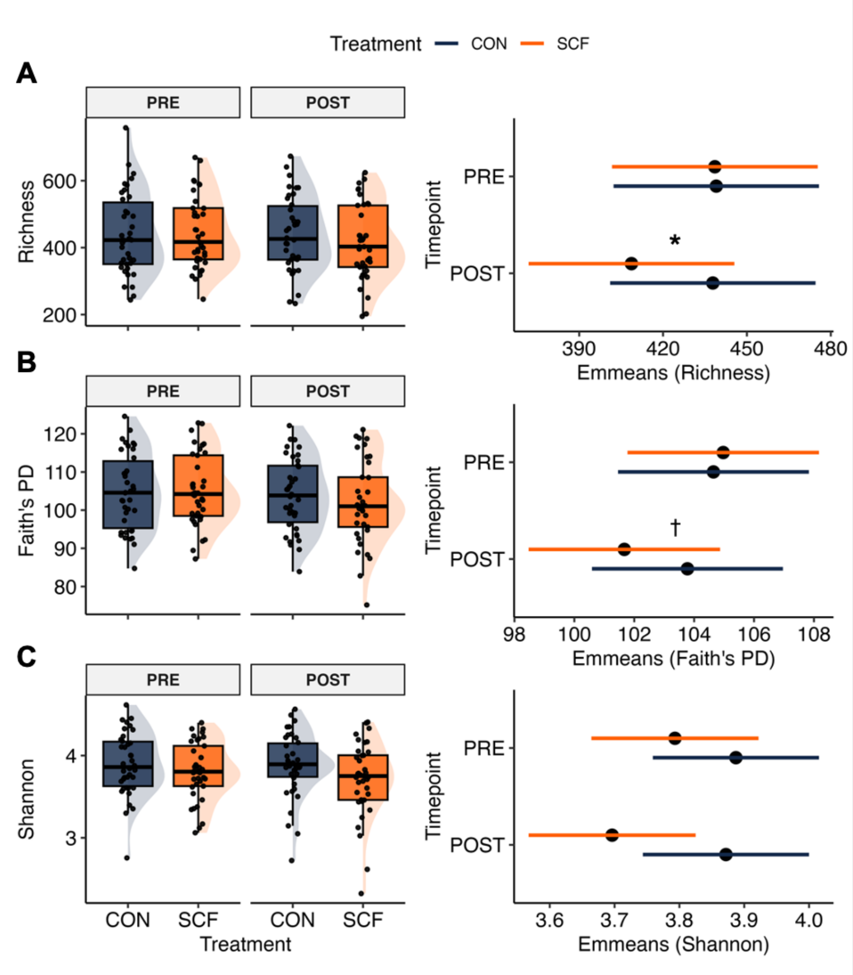


**Supplemental Figure 7. 16S Alpha Diversity across conditions using different alpha metrics.** *n* = 38 CON and *n* = 37 SCF. The alpha diversity plots demonstrate the (**A**) richness, (**B**) Shannon, and (**C**) Faith's phylogenetic diversity metrics across timepoints. Each point represents a sample, with boxplots coloring denoting the specific treatment. Statistical significance derived from the linear mixed effect model described in the methods. FDR adjusted *p*-values annotated as follows: * < 0.05; † < 0.1.

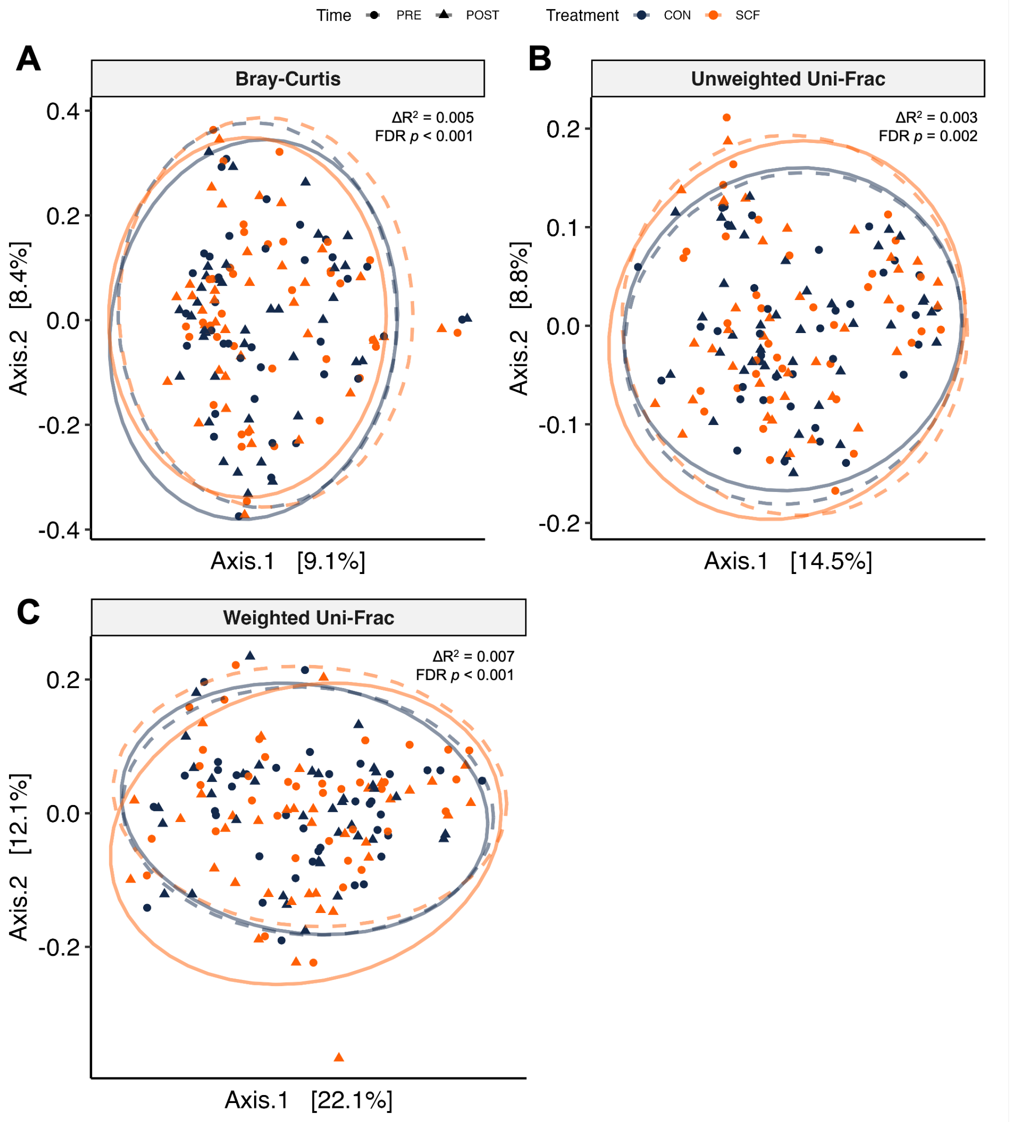


**Supplemental Figure 8. 16S Beta Diversity principal coordinates analyses**. ($n$ = 38 CON, $n$ = 37 SCF). Principal Coordinates Analysis (PCoA) of Bray-Curtis dissimilarity and (**A**), unweighted Uni-Frac (**B**), and weighted Uni-Frac (**C**). Points represent individual samples, colored by treatment (CON = blue, SCF = orange) and shaped by collection time (baseline = circles, endpoints = triangles). Ellipses indicate 95% confidence intervals, with dashed lines for baseline and solid lines for endpoints of interventions. Axes show the first two principal coordinates, with variance explained [%]. Reported are $R^2$ and FDR-adjusted $p$ values from the treatment-by-time interaction term from the PERMANOVA (5000 permutations; blocking by ID).

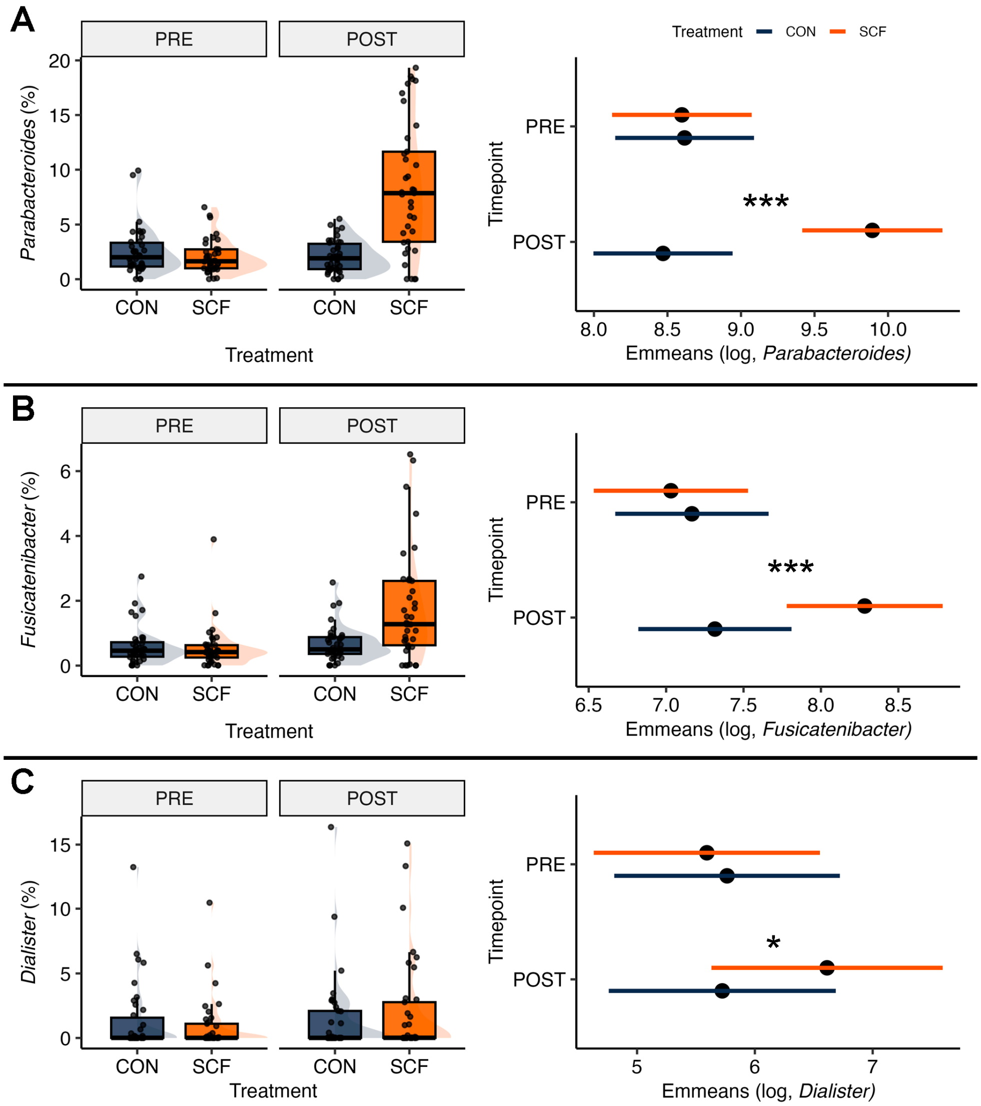


**Supplemental Figure 9. Distribution and *post-hoc* comparison of 16S selected genera.** Relative abundances (% of sequences) on the left and estimated marginal pairwise comparisons (Emmeans, right; log scale) of *Parabacteroides* (**A**), *Fusicatenibacter* (**B**), and *Dialister* (**C**) across conditions. *Post-hoc* analyses show FDR-adjusted *p* between treatment at endpoint. Statistical significance is annotated as follows: *** < 0.001; ** < 0.01; * < 0.05; † < 0.1.

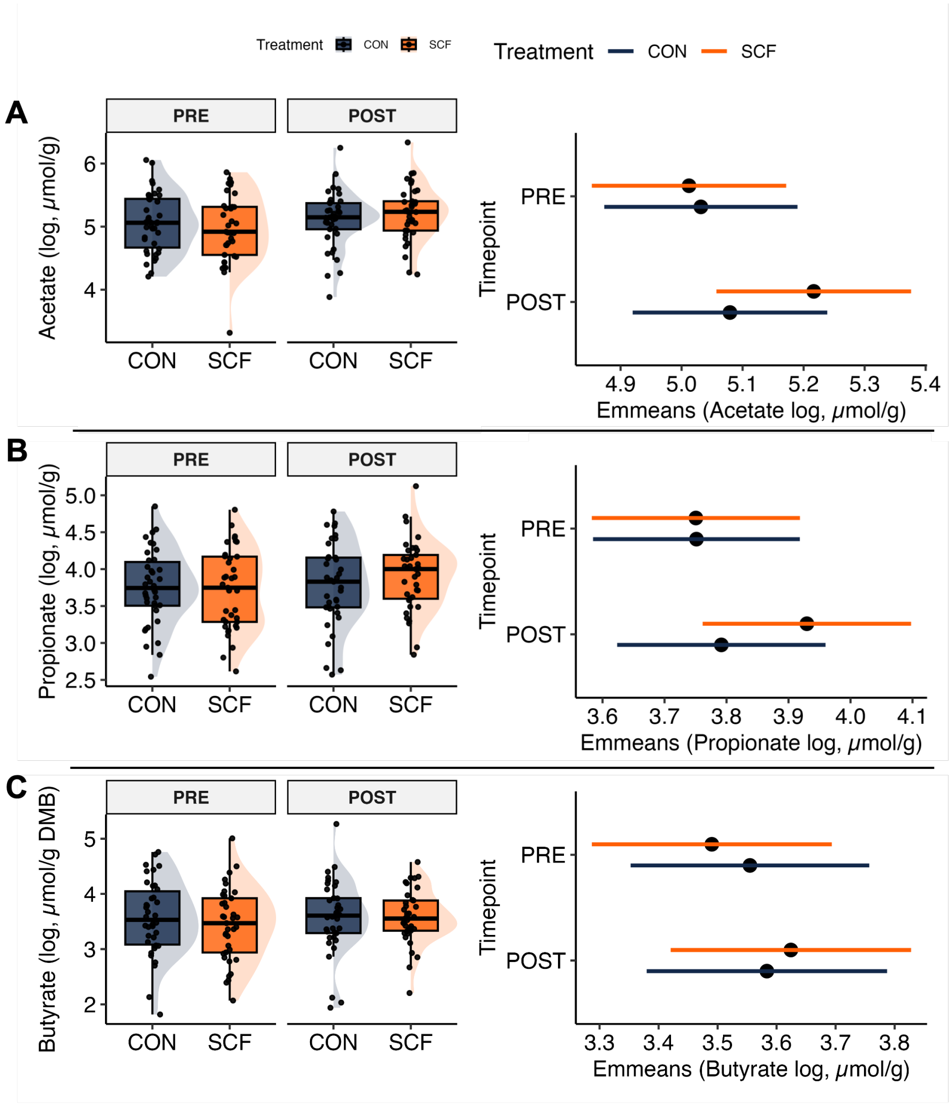


**Supplemental Figure 10. Distribution and exploratory *post-hoc* comparison of SCFAs.** Fecal short chain fatty acids (SCFA) on a dry matter basis (DMB) from the intervention on the left and estimated marginal pairwise comparisons (Emmeans, right; log scale) of **(A)** *acetate*, (**B**) *propionate*, (**C**) and *butyyrate* across conditions. *Post-hoc* analyses show FDR-adjusted *p* between treatment at endpoint. Statistical significance is annotated as follows: *** < 0.001; ** < 0.01; * < 0.05; † < 0.1.

### ***SUPPLEMENTAL TABLES & FIGURES***

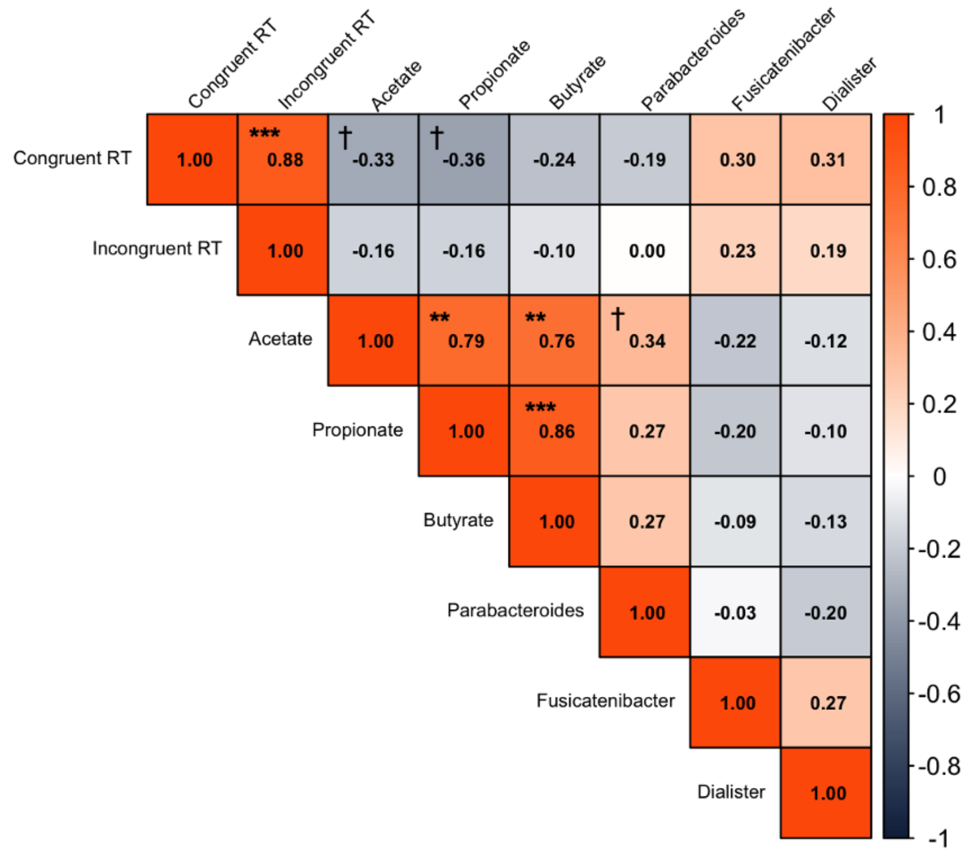


**Supplemental Figure 11. Spearman correlation between reaction times, fecal SCFAs, and 16S genera following soluble corn fiber intervention at endpoint.** Heatmap displays Spearman's coefficients (ρ) between the markers of interest and the flanker task reaction times (*n* = 37; SCF arm only). Positive correlations are orange, negative correlations blue, with intensity reflecting the magnitude. Spearman *p*-values were FDR adjusted. Statistical significances are annotated as follows: *p* *** < 0.001, † < 0.1.

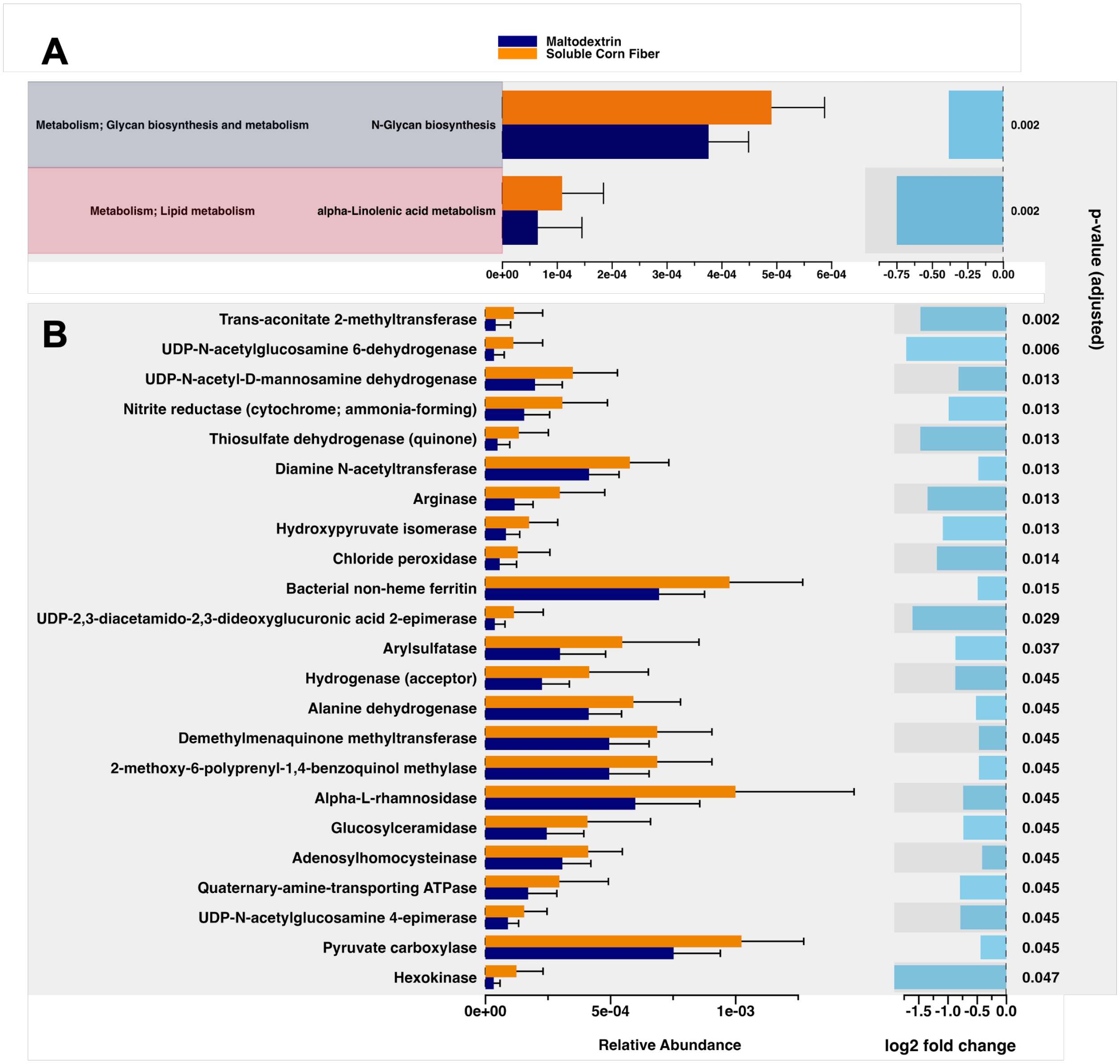


**Supplemental Figure 12. Linear differential abundance (LinDA) analysis of PICRUSt2-predicted functional profiles from endpoint samples.** ($n$ = 39). (**A**) KEGG Orthology (KO) pathways and (**B**) Enzyme Commission (EC) numbers showing mean relative abundance ± SEM, with $\log_2$ fold-changes and FDR adjusted $p$-values displayed alongside. Only features with FDR $p < 0.05$ are shown. Figures generated through ggpicrust R-studio package.

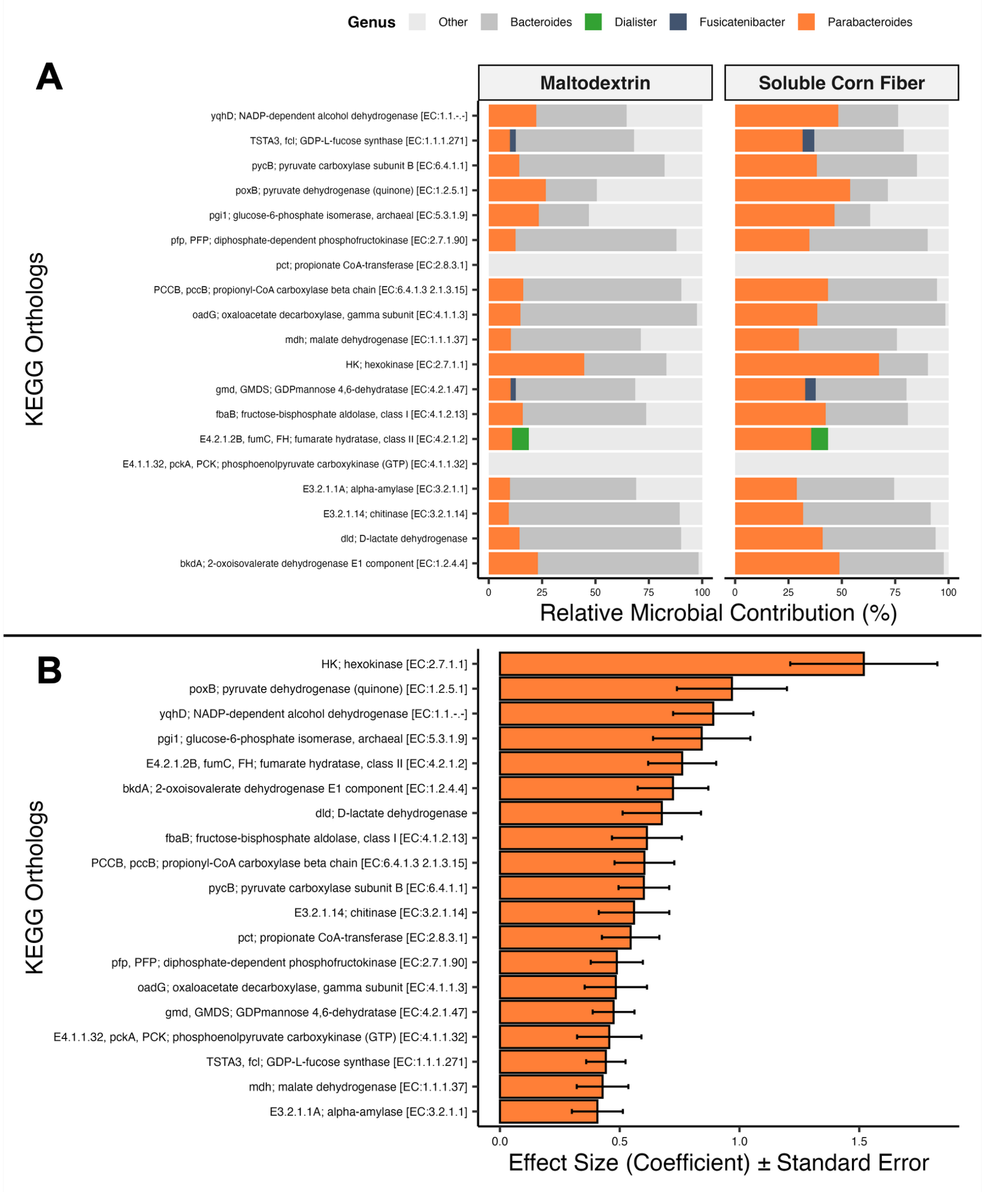


**Supplemental Figure 13. Differential abundance of predicted carbohydrate KOs at endpoint between treatments**. *n=39* (**A**) Relative microbial contribution of selected genera across all participants for 18 of the 20 KOs with the highest MaAsLin coefficients significantly associated with the soluble corn fiber intervention. Propionate CoA-transferase [EC:2.8.3.1] and phosphoenolpuruvate carboxykinase [EC:4.1.1.32] did not have detectable relative contribution from the selected genera of interest. This reflects the end-point facetted by treatment. The genera of interest were colored orange, green or blue, and Bacteroides colored in gray for each carbohydrate-metabolism KO. (**B**) KOs effect size from MaAsLin2 coefficients estimating the direction and magnitude of the association between soluble corn fiber (SCF) treatment and KO abundance relative to control.

.

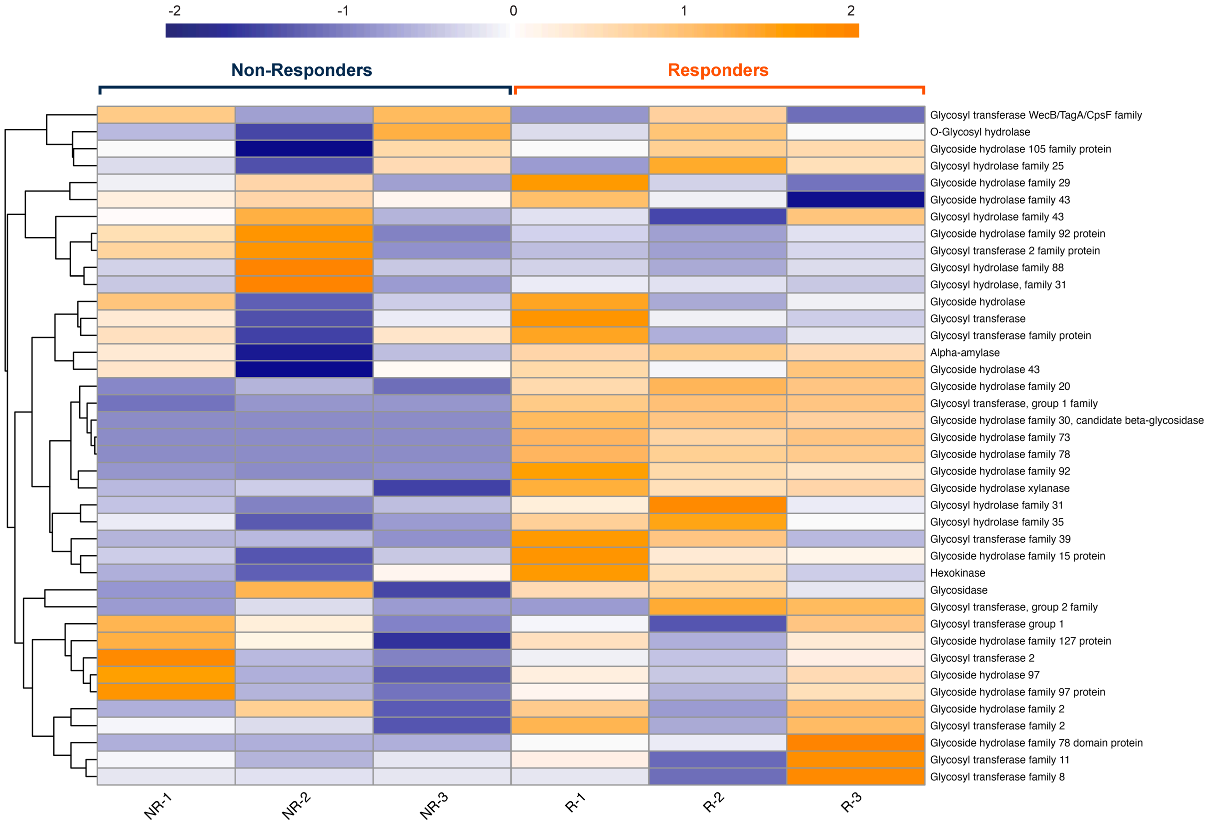


**Supplemental Figure 14. Abundance of carbohydrate metabolism-associated gene family comparisons between responders and non-responders at endpoint of the SCF intervention.** Heatmap displaying the abundance of carbohydrate metabolism-related gene families annotated by HUMAnN 3.0 across six participants (N = 6), stratified by responder (R1–R3) and non-responder (NR1–NR3) status. Genes families shown (rows) were selected from a total of 1,180 unique carbohydrate-associated gene families based on a $\log_2$ fold change > 2 in mean abundance between responders and non-responders [log2(R/NR)]. The top 40 gene families are presented. Abundance values were row-scaled using Z-score normalization to emphasize relative variation across samples. Orange indicates higher abundance, and blue indicates lower abundance for each gene.

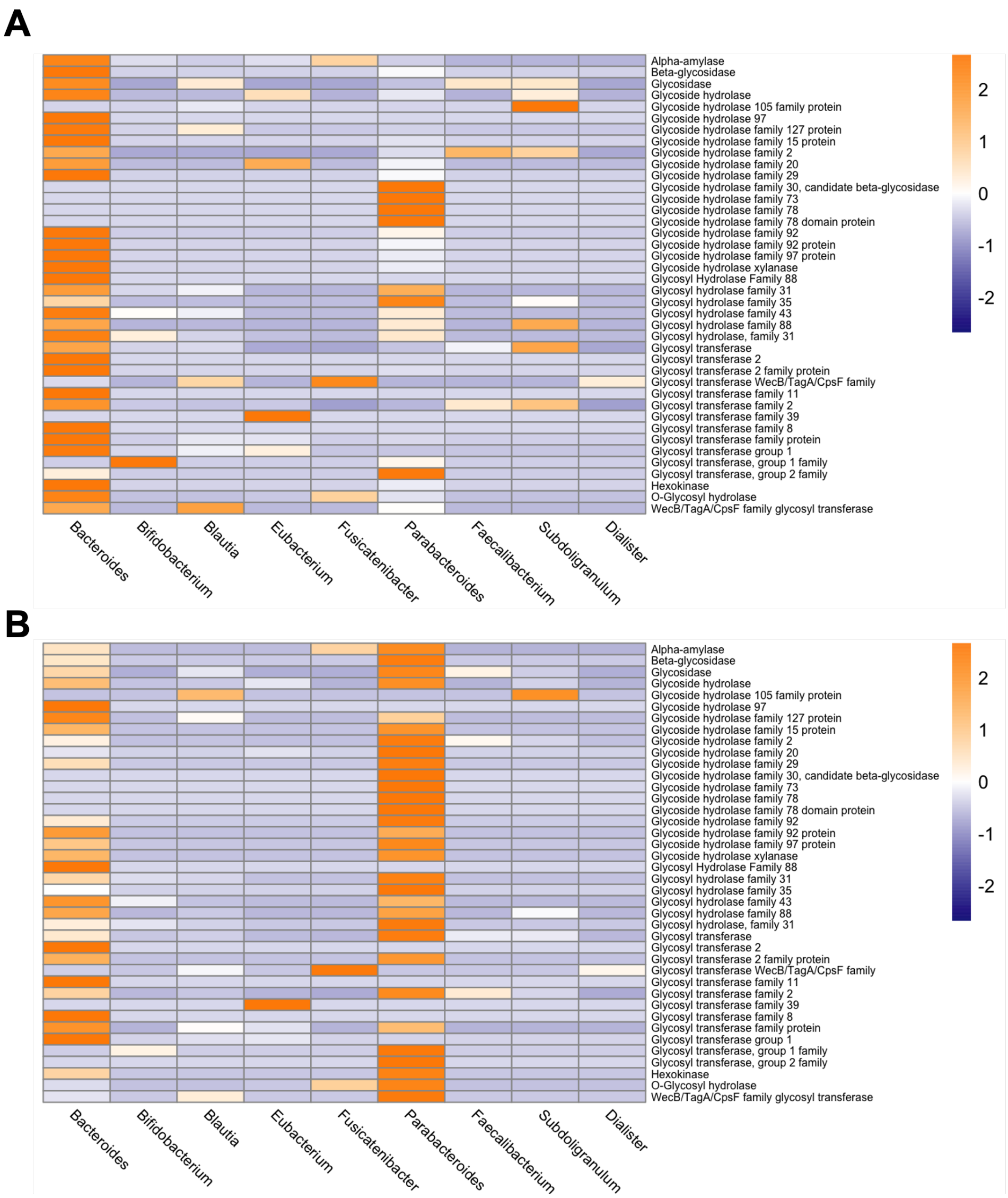


**Supplemental Figure 15. Differential Genus Contribution to Carbohydrate Metabolism Gene HUMAnN Profiles before and after the SCF intervention.** N = 6. The heatmap visualizes the contributions of key microbial genera (columns) to the abundance of carbohydrate metabolism-associated genes (rows). Each gene's abundance was aggregated by genus, and row scaling was applied to normalize the abundance of each gene across the selected genera. This scaling highlights the relative contributions of genera to each gene, with orange indicating higher abundance and blue indicating lower abundance for that gene across genera.

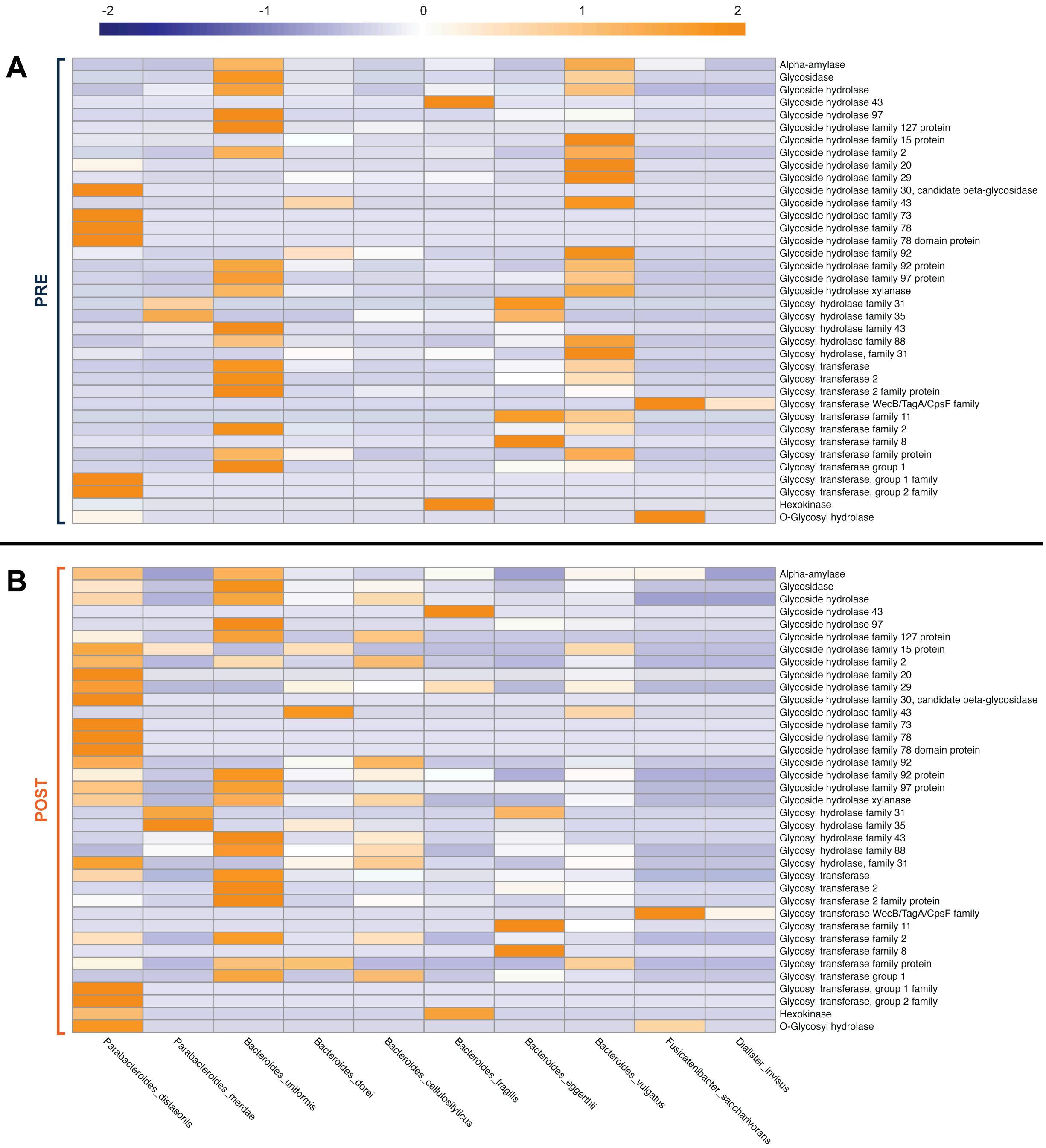


**Supplemental Figure 16. Responders' species-level profile contributions over time to carbohydrate metabolism gene family abundances.** Heatmaps showing the contribution of individual species (columns) to the abundance of 40 carbohydrate metabolism-associated gene families (rows) of responders. The top panel (**A**) shows the baseline timepoint while the bottom panel (**B**) shows the endpoint of the SCF intervention. Gene abundance was row-normalized using Z-scores to highlight species-level contributions, with orange indicating higher and blue indicating lower abundance for each gene.

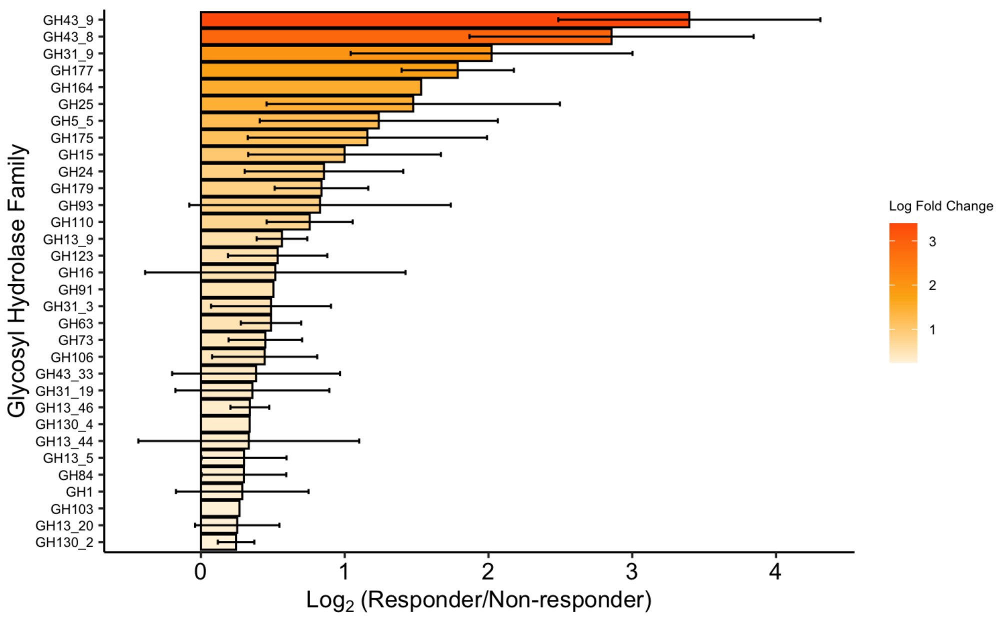


**Supplemental Figure 17. Log$_2$ Fold Change in CAZy Glycosyl Hydrolase (GH) Family Abundance Between Responders and Non-Responders after SCF intervention.** Log$_2$ fold change values comparing the relative abundance of glycosyl hydrolase (GH) families and subfamilies (denoted by "_#") in responders versus non-responders (N = 6), based on CAZy annotations. Only GH families with a positive association in responders (log$_2$FC > 0.2) are displayed. Standard errors (SE) were calculated using error propagation from group-level standard errors. Bars are color-scaled by log$_2$FC magnitude (white to red) and are presented in descending order of fold change.

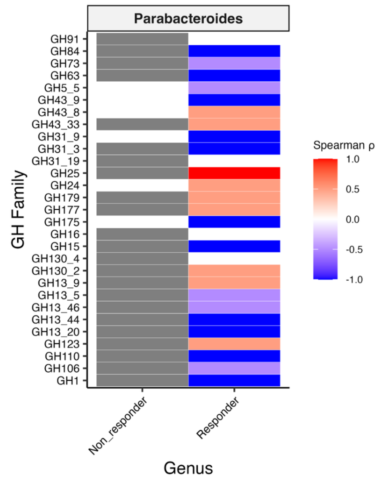


**Supplemental Figure 15. Spearman correlation analysis of Parabacteroides-associated and CAZy-GH family abundance in Responders vs. Non-Responders at endpoint of SCF intervention.** N = 6 (Responders = 3, Non-responders=3). Data is derived from the CAZy database, highlighting glycoside hydrolase (GH) families that exhibited a positive association with Responders (log$_2$FC > 0.2). Bars are colored according to Spearman's ρ, where red represents positive correlations and blue represents negative correlations with Parabacteroides abundance. Gray bars indicate cases where Parabacteroides data was unavailable, preventing correlation analysis for the corresponding GH family.
